# Treatment outcomes of drug-resistant *Mycobacterium tuberculosis* infection in Cameroon: a systematic review and meta-analysis

**DOI:** 10.64898/2026.08.04.26359729

**Authors:** Fabrice Zobel Lekeumo Cheuyem, Arole Darwin Touko, Chabeja Achangwa, Rick Tchamani, Raïssa Katy Noa Otsali, Clovis Josué Kachiwouo Mapouo, Mazou Ngou Temgoua

## Abstract

**Background:** Drug-resistant tuberculosis (DR-TB) remains a major challenge to tuberculosis control in sub-Saharan Africa. Cameroon faces substantial challenges in managing DR-TB; however, national evidence on treatment outcomes remains unsynthesized. This systematic review and meta-analysis aimed to estimate pooled treatment outcomes, adverse drug events (ADEs), and predictors of unfavorable outcomes among patients with DR-TB in Cameroon.

**Methods:** This systematic review and meta-analysis followed the PRISMA 2020 guidelines. PubMed, Scopus, Embase, Web of Science, the Cochrane Library, African Journals Online. Google Scholar and reference lists were also searched. Studies reporting World Health Organization-defined treatment outcomes among patients with DR-TB were included. Random-effects meta-analyses using generalized linear mixed models with logit transformation were performed. Heterogeneity was assessed using the *I*² statistic, and publication bias and sensitivity analyses were conducted.

**Results:** Fifteen studies conducted between 1998 and 2022 were included. The pooled mortality rate was 6.8% (95% CI: 4.7–9.7; 14 reports; n = 2,351 participants), loss to follow-up was 4.1% (95% CI: 2.8–6.1; 12 studies; n = 2,244 participants), and treatment failure was 5.0% (95% CI: 1.1–19.8; 12 studies; n = 2,050 participants). The pooled treatment success rate was 74.2% (95% CI: 60.4–84.4; 13 reports; n = 2,146 participants). Treatment success improved over time and was higher with modified regimens (87.2%; 95% CI: 83.8–89.9; 3 studies; n = 460 participants) than with standard regimens (68.8%; 95% CI: 52.2–81.6; 10 studies; n = 1,686 participants). Among patients with multidrug-resistant-TB, the pooled prevalence of adverse drug events was 70.8% (95% CI: 40.2–89.7; 3 studies; n = 251 participants), with ototoxicity (41.9%; 95% CI: 23.7–62.6; 3 studies; n = 251 participants) and gastrointestinal disorders (40.9%; 95% CI: 25.5–58.2; 2 studies; n = 172 participants) being the most common events. HIV co-infection was significantly associated with unfavorable treatment outcomes (pooled OR = 2.76; 95% CI: 1.95–3.93; 6 studies), and male gender was also associated with increased odds of unfavorable outcomes (OR = 1.73; 95% CI: 1.25–2.40; 5 studies).

**Conclusions:** Approximately three-quarters of patients with DR-TB in Cameroon achieved successful treatment, although mortality, treatment failure, and adverse drug events remain important concerns. Strengthening pharmacovigilance, integrated TB/HIV care, and the implementation of effective all-oral regimens are essential to improve treatment outcomes.

**Systematic review registration number:** CRD420261404490.

## 1. Background

Tuberculosis (TB) continues to be among the leading infectious diseases causing morbidity and mortality around the globe. According to the World Health Organization (WHO) Global TB Report 2023, an estimated 10.6 million new TB cases and 1.35 million deaths were recorded globally in 2022 [1]. Drug-resistant tuberculosis (DR-TB) encompasses a spectrum of resistance patterns, ranging from mono-resistance to multidrug-resistant (MDR) and extensively drug-resistant forms, and poses a growing challenge to global TB control efforts [2].

Sub-Saharan Africa bears a heavy burden of tuberculosis globally. Africa makes up about 23-25% of the global TB incidence rate and more than one-third of deaths due to TB [3, 4]. The number of cases of MDR/RR-TB (multidrug-resistant/rifampicin-resistant tuberculosis) in the WHO African Region in 2024 is estimated to be 57,000, which is about 15% of the global MDR/RR-TB burden [1]. High prevalence rates of HIV infection make outcomes for patients with drug-resistant TB even worse, since co-infection significantly increases the risk of treatment failure and death from the illness [5].

According to WHO, Cameroon falls into the category of a high TB burden country in Africa. All TB patients are diagnosed and treated at the 248 diagnostic and treatment centers (DTCs) across the country under the coordination of the National TB Program. In 2016, there were notifications of 25,975 TB cases, out of which 175 were RR/MDR-TB cases [6]. WHO estimates that there would be about 1,200 (95% confidence interval [CI] 1,000-2,200) RR/MDR-TB cases annually in Cameroon, indicating a gap of more than 80% in notification rates.

According to a national cross-sectional study conducted in 2018, the prevalence of RR-TB among newly bacteriologically confirmed pulmonary TB patients was found to be 1.6% (95% confidence interval [CI]: 0.8–2.3%), slightly lower than the WHO estimated figure of 2.8% [6]. The prevalence of RR/MDR-TB among previously treated patients has been found to range between 14–16% [6]. In drug resistance studies conducted in the Littoral Region, the incidence of MDR-TB was found to be 4.5%, whereas in the Northwest and Southwest Regions, the prevalence of MDR-TB was 5.9% [7].

Treatment of DR-TB particularly its multidrug and rifampicin-resistant forms, involves extended second line therapy that is considerably more toxic and cloister than routine first-line treatment. Treatment success rates for MDR/RR-TB remain significantly lower than for drug-susceptible TB globally, and mortality associated with drug-resistant disease is considerable, especially in regions where the burden of HIV is high [8].

Since the mid-2000s, Cameroon has had specialized units for treating DR-TB predominantly MDR/RR-TB cases with Jamot Hospital in Yaoundé being the main referral hospital in the country. Beginning in 2008, patients eligible for the programme were treated using a standard 12-month shorter treatment regimen involving gatifloxacin, clofazimine, prothionamide, ethambutol, and pyrazinamide for the entire period, with kanamycin and isoniazid in addition to the above drugs used for a minimum four-month intensive period [9]. In a prospective observational study carried out at two national MDR-TB treatment units, 134 out of 150 (89%) patients achieved successful treatment a feat that garnered worldwide attention [9].

The more recent information from Yaoundé shows increased adverse outcomes within routine program settings. A retrospective cohort study conducted at Jamot Hospital for 2013–2019 showed that male gender, HIV coinfection, and moderate-to-severe anemia were independent predictors of unfavorable treatment outcomes in rifampicin-resistant pulmonary tuberculosis patients [10]. The database study from the Littoral Region during 2013–2022 revealed that the overall treatment success rate among DR-TB patients was 87.2%, and the risk factors for adverse outcomes included male gender, HIV status, and previous DR-TB treatment [11].

Despite these significant primary research efforts, no systematic review has been conducted to summarize the body of evidence on the outcomes of drug-resistant TB (DR-TB) treatment in Cameroon. There is currently a lack of uniformity in the way that the information is collected, due to the differences in design, duration, geography, and patient populations between studies.

The synthesis of treatment outcomes, which includes successful treatment, mortality, treatment failure, loss to follow-up, and adverse effects during treatment, is thus required to obtain estimates that are representative of the country, understand heterogeneity in treatment outcomes, and identify factors that contribute to poor treatment outcomes. This evidence is important for the National TB Program in providing services for drug resistant TB patients and tracking their progress toward the End TB Strategy objectives. The following protocol outlines the process of conducting such a review and meta-analysis.

Despite substantial primary research efforts, no systematic review has yet synthesized the evidence on DR-TB treatment outcomes in Cameroon. Available data remain heterogeneous due to differences in study design, duration, geographic setting, and patient populations. A comprehensive synthesis of treatment outcomes including treatment success, mortality, loss to follow-up, treatment failure, and adverse events is therefore needed to produce nationally representative estimates, understand outcome heterogeneity, and identify predictors of poor outcomes. Such evidence is essential for the National TB Program to optimize service delivery for DR-TB patients and to monitor progress toward End TB Strategy targets.

## 2. Methods

### 2.1. Study design and registration

This systematic review and meta-analysis was conducted following the Preferred Reporting Items for Systematic Reviews and Meta-Analyses (PRISMA) 2020 guidelines [12]. The protocol was registered in the International Prospective Register of Systematic Reviews (PROSPERO; CRD420261404490).

### 2.2. Eligibility criteria

#### Inclusion criteria

Studies were considered eligible if they: (1) were conducted in Cameroon, (2) included patients of any age with bacteriologically confirmed or clinically diagnosed drug-resistant tuberculosis (DR-TB), (3) employed an observational or interventional design (4) reported at least one WHO-defined treatment outcome (treatment success [cure and treatment completion], treatment failure, death, or loss to follow-up) or adverse drug reaction data; (5) were published in English or French and (6) involved human subjects.

#### Exclusion criteria

Case reports, narrative reviews, systematic reviews, meta-analyses, editorials, opinion pieces, conference abstracts, animal studies, in vitro or laboratory-only investigations, drug resistance surveillance studies not reporting patient-level treatment outcomes, and studies reporting outcomes only for drug-susceptible tuberculosis without disaggregation for DR-TB were excluded. Duplicate publications of the same dataset were excluded, with the most complete and recent publication retained for analysis.

### 2.3. Outcomes measurement and definition

The primary outcomes included the pooled rates of treatment success (cured and treatment completion), loss to follow-up, treatment failure, mortality, and adverse drug reactions. Treatment outcomes were defined according to WHO criteria [13]. Treatment outcomes were defined according to WHO guidelines. Treatment failure was defined as a patient whose treatment regimen needed to be terminated or permanently changed to a new regimen or treatment strategy. Cured was defined as a patient with pulmonary TB with bacteriologically confirmed TB at the beginning of treatment who completed treatment as recommended by the national policy, with evidence of bacteriologic response and no evidence of failure. Treatment completed was defined as a patient who completed treatment as recommended by the national policy but whose outcome did not meet the definition for cure or treatment failure. Died was defined as a patient who died before starting treatment or during the course of treatment. Lost to follow-up (LTFU) was defined as a patient who did not start treatment or whose treatment was interrupted for two consecutive months or more. Treatment success was defined as the sum of all patients cured and treatment completed. The prevalence of each outcome was calculated by dividing the number of patients experiencing the specific outcome by the total number of treated patients and presented in percentage. The secondary outcomes included factors associated with unfavorable outcomes. Unfavorable outcome included patients who were declared dead, LFTU, and treatment failure [14–16].

### 2.4. Search strategy

A comprehensive search was conducted in the following electronic databases: PubMed, Embase, Scopus, Web of Science, African Journals Online (AJOL), and the Cochrane Library. Grey literature was searched on Google Scholar, and reference lists of all selected studies were hand-searched. The search strategy employed controlled vocabulary terms (MeSH and Emtree) and free-text keywords using Boolean operators. Grey literature will be searched on Google Scholar, and reference lists from all selected studies will be hand-searched. For PubMed search strategy is presented below: (“MDR-TB”[tiab] OR “multidrug-resistant tuberculosis”[tiab] OR “multi-drug resistant tuberculosis”[tiab] OR “drug-resistant tuberculosis”[tiab]) AND (“Treatment Outcome”[MeSH] OR “treatment outcome”[tiab] OR “treatment success”[tiab] OR “cure rate”[tiab] OR “treatment completion”[tiab] OR “treatment failure”[tiab] OR “lost to follow-up”[tiab] OR “mortality”[tiab] OR “death”[tiab]) AND (“Cameroon”[MeSH] OR “Cameroon”[tiab] OR “Cameroun”[tiab]). The full search strategy by database is presented in Supplementary Materials (**Table S1**). The last search was performed on April 15, 2026.

All retrieved articles underwent deduplication and initial screening. Duplicates were excluded, and the remaining articles were screened against the inclusion criteria. Full texts of all articles were independently screened by two reviewers (FZLC and RT). Disagreements were resolved through discussion or consulting a third reviewer (MNT).

### 2.5. Data extraction

Data extraction was performed independently by two reviewers (FZLC and ADT) using a standardized data extraction form. Extracted information included study characteristics (first author, year of publication, year of study completion, study design, setting, region, sample size, tuberculosis infection localization, type of drug-resistant TB regimen, and outcome-related data. For each study, the total number of treated patients and the number with each treatment outcome (treatment success, death, loss to follow-up, and treatment failure) were extracted. Data on factors associated with unfavorable outcomes were also extracted. Disagreements between investigators were resolved through discussion or consultation with a third reviewer (MNT).

### 2.6. Risk of bias assessment

The quality and risk of bias of the included studies were assessed independently by two reviewers (FZLC and CA) using the Joanna Briggs Institute (JBI) critical appraisal tools, selected according to the design of each study. Disagreements were resolved by discussion or, when necessary, by involving a third reviewer (MNT) [17]. For cohort studies, the checklist contains eleven items assessing: the two groups were similar and recruited from the same population; exposures were measured similarly to assign people to exposed and unexposed groups; the exposure was measured in a valid and reliable way; confounding factors were identified and strategies to deal with them stated; whether the groups/participants were free of the outcome at the start of the study (or at the moment of exposure); outcomes were measured in a valid and reliable way; whether the follow-up time was sufficient for outcomes to occur; follow-up was complete, with reasons for loss to follow-up described and explored; strategies to address incomplete follow-up were utilized; and appropriate statistical analysis was used. Each criterion was scored as 1 (yes) or 0 (no or unclear). In this study, we defined and applied the following categorization of the overall risk of bias: low = ≥70%, moderate = 50% – 69%, and high risk of bias = <50% of “Yes” scores.

### 2.7. Statistical analysis

Pooled prevalence estimates and associated factors were generated using a random-effects model (DerSimonian-Laird method). Heterogeneity was assessed using the *I*² statistic, with values categorized as low (<25%), moderate (25–75%), or high (>75%). The generalized linear mixed models (GLMM) with probit-logit transformation (PLOGIT) were employed to effectively handle meta-analyses of binary data and accommodate studies with 0% or 100% event rates without requiring continuity corrections [18]. Where sufficient data were available, subgroup analyses were conducted for each outcome. Statistical significance was defined as *p-*value < 0.05. All analyses were conducted using the ‘meta’ package in R Statistics version 4.5.2 [19]. Factors significantly associated with unfavorable outcomes, reported in only one study, were synthesized narratively.

### 2.8. Publication bias and sensitivity analysis

Publication bias was assessed graphically using funnel plots and statistically using Egger’s and Begg’s tests [20, 21]. A *p-*value < 0.05 was considered evidence of significant publication bias. Furthermore, a leave-one-out sensitivity analysis was performed to determine the influence of individual studies on the overall findings.

## 3. Results

### 3.1. Study selection and inclusion

A total of 291 records were identified through database searches (PubMed, n = 13; Scopus, n = 28; Web of Science, n = 107; Embase, n = 34; Cochrane Library, n = 13; African Journals Online, n = 96). After removing 23 duplicate records, 268 records were screened based on titles and abstracts. Of these, 230 records were excluded as they did not meet the eligibility criteria. The remaining 38 full-text reports were assessed for eligibility. Of these, 24 were excluded for various reasons. Additionally, 1,833 records were identified through other methods, comprising 176 from Google Scholar and 1,657 via reference list searching. After screening, 19 reports were sought for retrieval and assessed for eligibility, and 18 were excluded because they did not report the outcome of interest. In total, 15 studies were included in the final review (**Fig. 1**).

**Fig. 1.**
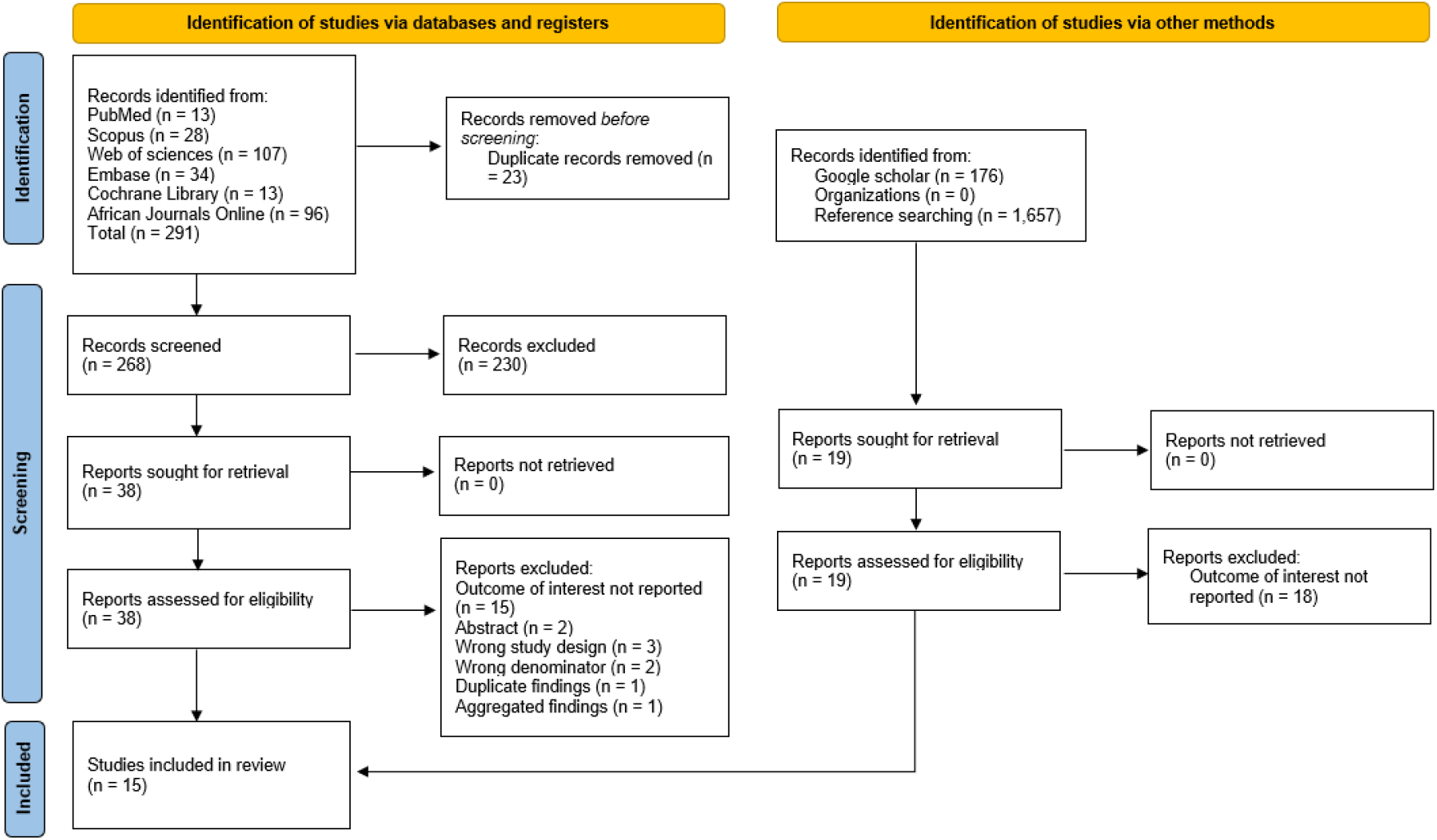
PRISMA diagram flow from study identification to inclusion in the systematic review and meta-analysis [12]

### 3.2. Characteristics of included studies

A total of 15 studies, published between 2002 and 2024, were included in this review, yielding 18 eligible reports based on distinct participant subgroups or treatment regimens. These studies were conducted across various regions of Cameroon, including the West, Littoral, Centre, North-West, South-West, and multi-regional settings. The majority were cohort studies (n = 14), with three cross-sectional studies and one case-control study. Most studies focused on patients with pulmonary tuberculosis (PTB; n = 14), with a few including both PTB and extrapulmonary TB (EPTB; n = 3).

The sample sizes ranged from 6 to 693 participants. Most studies (n = 13) reported on patients receiving standard treatment regimens, while three studies evaluated modified regimens, including fluoroquinolone-resistant (FQR-Mtr) and second-line injectable-resistant (SLIr-Mtr) regimens, primarily among RR-TB or multidrug-resistant (MDR-TB) patients and patients from the general population.

Treatment outcomes were variably reported across studies. Among studies that reported treatment success, rates ranged from 0.0% to 100%. Death occurred in 0.0% to 14.9% of patients across studies reporting this outcome. Lost to follow-up ranged from 0.0% to 12.8%, and treatment failure from 0.0% to 71.4% (**Table 1**).

**Table 1.**
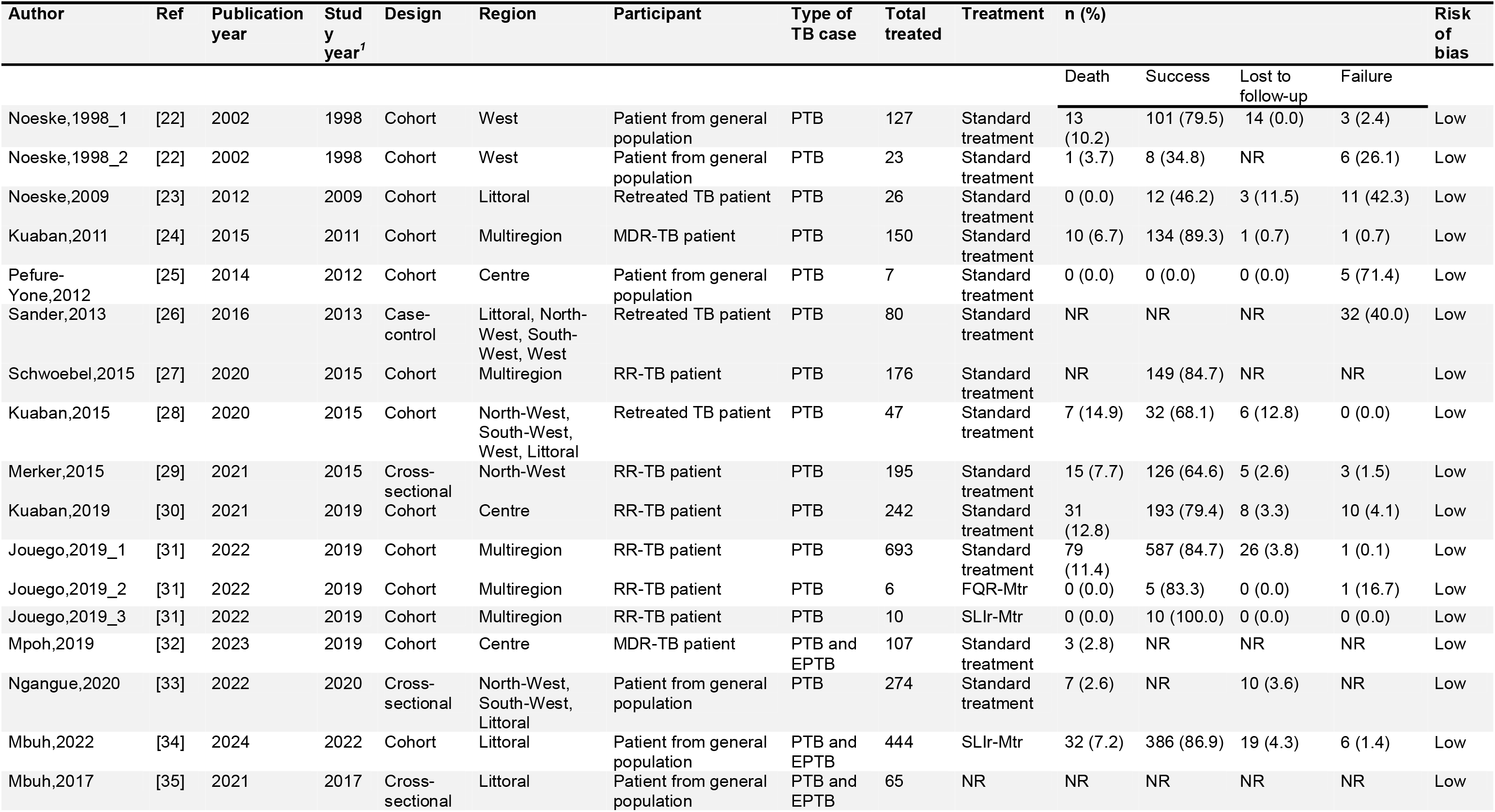
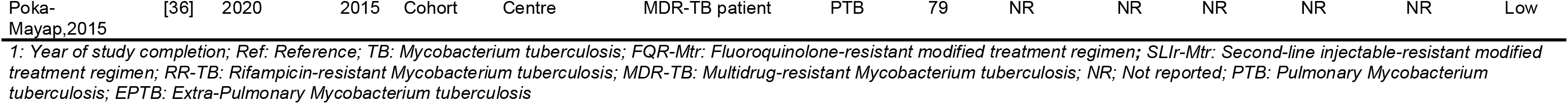
Synthesis of included studies characteristics.

### 3.3. Drug-resistant tuberculosis mortality

The pooled mortality rate estimated with a random-effects model was 6.8% (95% CI: 4.7%-9.7%) among 2,351 participants from 14 study reports. The largest study (n = 693) reported a mortality rate of 11.4% (95% CI: 9.1%-14.0%). Heterogeneity across studies was moderate (*I*² = 58.1%; *p* < 0.003) (**Fig. 2**). The finding was robust in the sensitivity analysis, and there was no significant sign of publication bias (Egger’s [*p* = 0.062] and Begg’s test [*p* = 0.784]) (**Figs S1-2**).

**Fig. 2.**
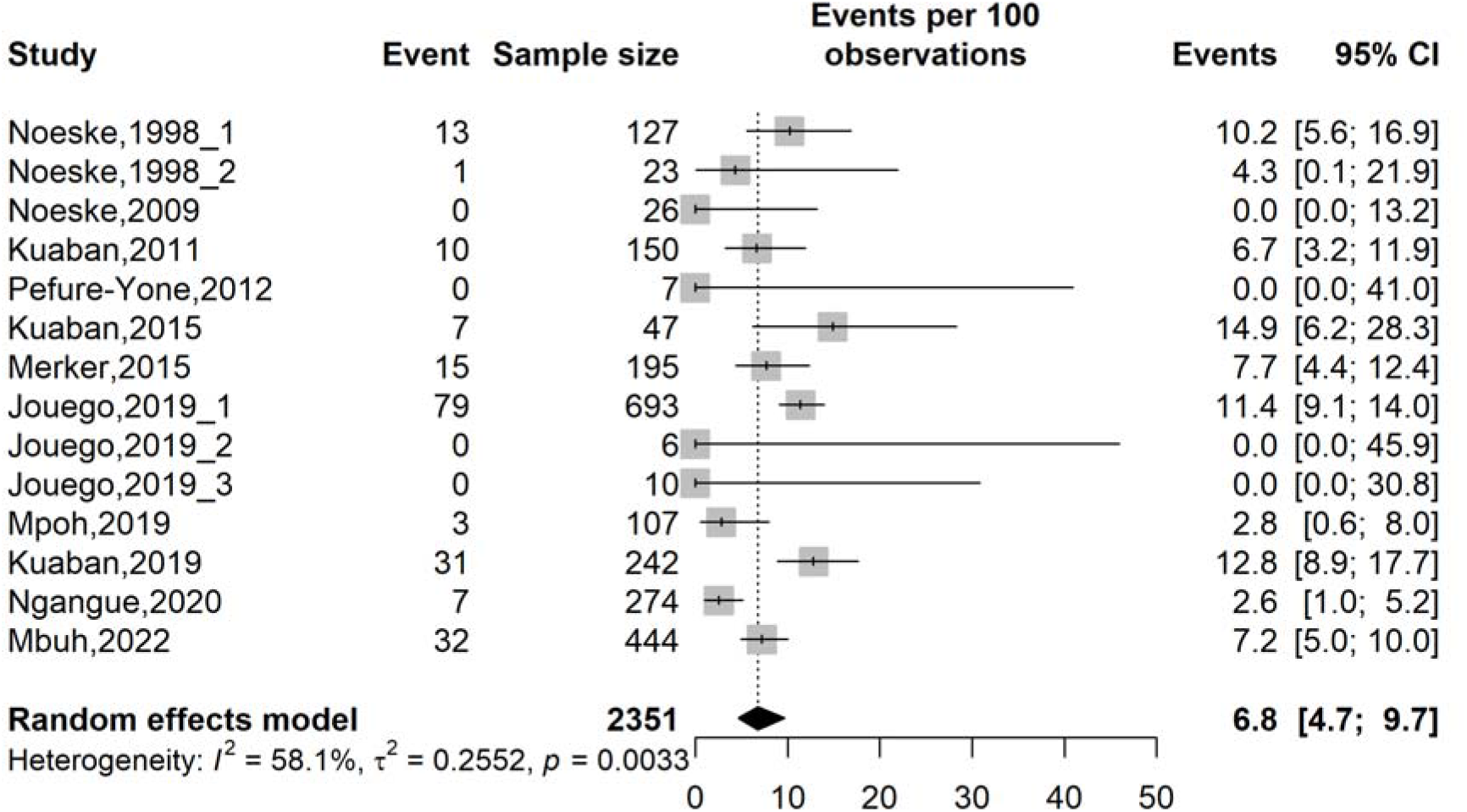
Forest plot of the pooled mortality rate among anti-tuberculosis drug-resistant patients in Cameroon, 1998-2022

The type of participants was identified as a significant source (*p* = 0.007) of heterogeneity among various subgroups explored for the pooled mortality rate among drug-resistant tuberculosis patients. Patients with RR-TB had the highest pooled mortality at 10.9% (95% CI: 9.2%-12.8%; 5 studies; n = 1,146) with no heterogeneity (*I*² = 0.0%). In contrast, patients with MDR-TB had a mortality of 5.0% (95% CI: 3.0%-8.3%; 3 studies; n = 280), while retreated patients and those from the general population had intermediate estimates of 4.8% and 5.6%, respectively. Subgroup analyses revealed no statistically significant differences in pooled mortality estimates by study period, study design, region, tuberculosis localization, or treatment regimen (**Table 2 and Figs. S3-8**).

**Table 2.** Subgroup meta-analysis of mortality rate among drug-resistant tuberculosis patients in Cameroon, 1998–2022.

| Subgroup | Sample size | Prevalence<sup>1</sup> (%) | 95% CI limits<sup>1</sup> |  | Number of reports | Heterogeneity statistic<sup>1</sup> |  | Subgroup difference *p*-value |
| --- | --- | --- | --- | --- | --- | --- | --- | --- |
|  |  |  | Lower | Upper |  | *I*<sup>2</sup>(%) | *p*-value |  |
| **Study period (year)<sup>2</sup>** |  |  |  |  |  |  |  | 0.637 |
| └ 2005 | 150 | 9.3 | 5.6 | 15.1 | 2 | 0.0 | 0.387 |  |
| 2005-2009 | 26 | 0.0 | 0.0 | 13.2 | 1 | - | - |  |
| 2010-2014 | 157 | 6.4 | 3.5 | 11.4 | 2 | 0.0 | 0.999 |  |
| 2015-2019 | 1,300 | 9.0 | 5.9 | 13.3 | 7 | 39.8 | 0.126 |  |
| 2020+ | 718 | 4.6 | 2.2 | 13.1 | 2 | 84.7 | 0.011 |  |
| **Study design** |  |  |  |  |  |  |  | 0.329 |
| Cross-sectional | 469 | 4.5 | 2.1 | 9.6 | 2 | 83.6 | 0.013 |  |
| Cohort | 1,882 | 8.0 | 5.6 | 11.2 | 12 | 33.5 | 0.122 |  |
| **Region** |  |  |  |  |  |  |  | 0.850 |
| Centre | 356 | 5.9 | 1.8 | 17.5 | 2 | 0.0 | 0.999 |  |
| Littoral | 470 | 6.8 | 4.9 | 9.5 | 2 | 0.0 | 0.999 |  |
| North-West | 195 | 7.7 | 4.4 | 12.4 | 1 | - | - |  |
| West | 150 | 9.3 | 5.6 | 15.1 | 2 | 0.0 | 0.387 |  |
| Multi-region | 1,180 | 6.5 | 3.4 | 12.2 | 6 | 73.4 | 0.002 |  |
| **Participant** |  |  |  |  |  |  |  | 0.007 |
| Patient from general population | 852 | 5.6 | 3.0 | 10.3 | 4 | 68.7 | 0.022 |  |
| Retreated tuberculosis patient | 73 | 4.8 | 0.3 | 48.3 | 2 | 0.0 | 0.999 |  |
| Rifampicin-resistant tuberculosis | 1,146 | 10.9 | 9.2 | 12.8 | 5 | 0.0 | 0.552 |  |
| Multidrug-resistant tuberculosis | 280 | 5.0 | 3.0 | 8.3 | 3 | 0.0 | 0.391 |  |
| **Tuberculosis infection localization** |  |  |  |  |  |  |  | 0.637 |
| Pulmonary | 1,800 | 7.2 | 4.8 | 10.7 | 12 | 51.7 | 0.019 |  |
| Mixed | 551 | 6.4 | 4.6 | 8.7 | 2 | 61.6 | 0.107 |  |
| **Treatment regimen<sup>3</sup>** |  |  |  |  |  |  |  | 0.955 |
| Standard treatment | 1,891 | 6.9 | 4.5 | 10.3 | 11 | 64.4 | 0.002 |  |
| Other regimen | 460 | 7.0 | 5.0 | 9.7 | 3 | 0.0 | 1.000 |  |

1: Random effects model; 2: Time frame aligns with the development of successive National Strategic Plans against tuberculosis in Cameroon; CI: Confidence interval; Mixed included pulmonary and extrapulmonary tuberculosis cases; 3: Other regimen included fluoroquinolone resistant modified treatment regimen [FQR-Mtr] and second-line injectable-resistant modified treatment regimen [SLIr-Mtr]

### 3.4. Drug-resistant tuberculosis treatment success

The overall pooled success rate was 74.2% (95% CI: 60.4%-84.4%; 13 reports) among 2,146 patients. However, high heterogeneity was observed across studies (*I*² = 88.1%; *p* < 0.001). Individual study success rates from 0.0% (95% CI: 0.0%-41.0%) to 100.0% (95% CI: 69.2%-100.0%) (**Fig. 3**). The sensitivity analysis revealed that the pooled estimate was robust, with no study significantly influencing the overall pooled estimate. There was no significant sign of publication bias (Egger’s test *p* = 0.191 and Begg’s test *p* = 0.222) (**Figs. S9-10**).

**Fig. 3.**
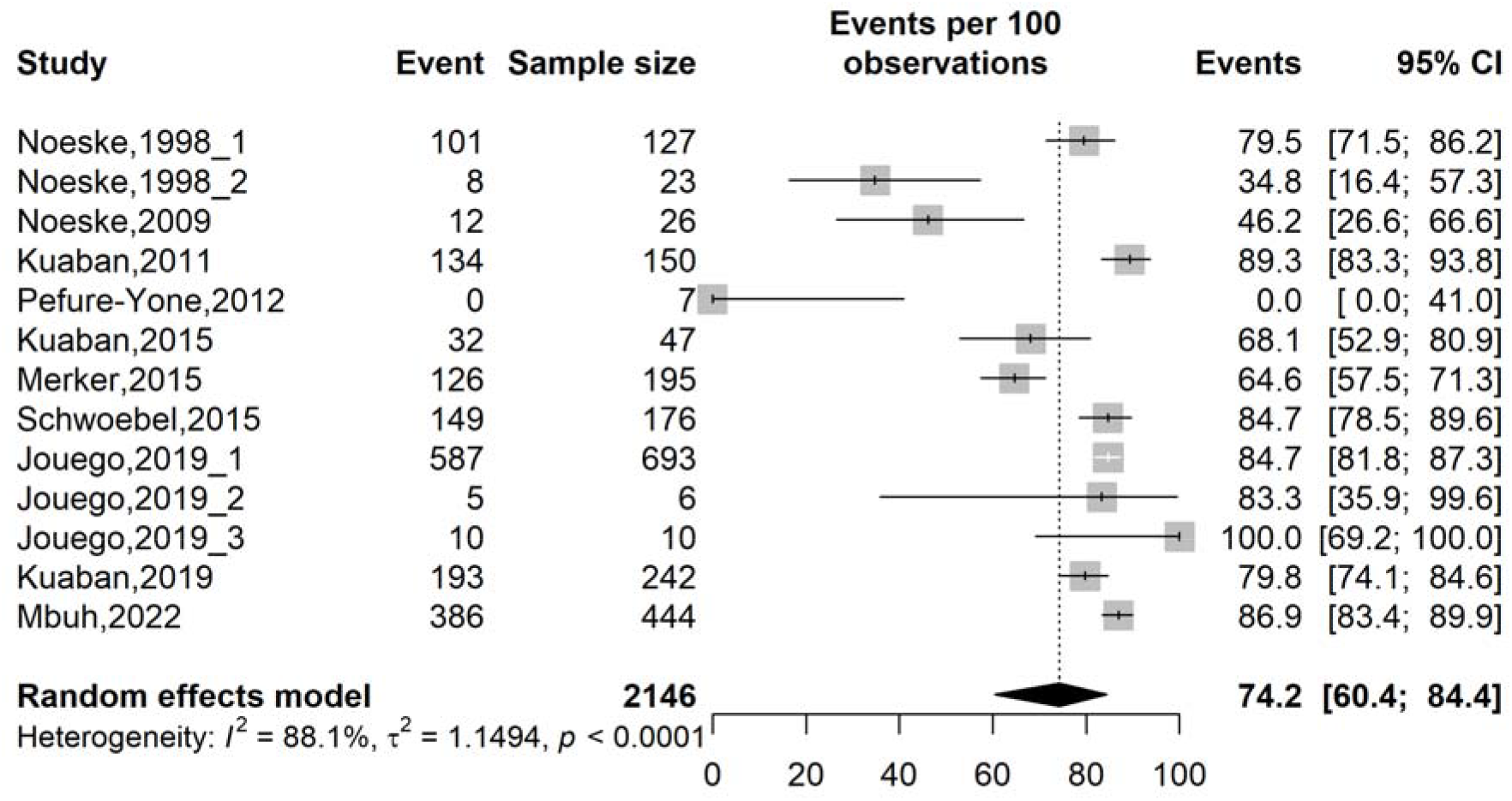
Forest plot of the pooled treatment success rate among anti-tuberculosis drug-resistant patients in Cameroon, 1998-2022

Subgroup analyses revealed significant effect modification for study period (*p* < 0.001), region (*p* < 0.001), infection localization (*p* = 0.014), and treatment regimen (*p* = 0.003). The success rates increased progressively over time; in the earliest period (<2005), the pooled estimate was 60.6% (95% CI: 27.5%-86.2%, 2 studies; n = 150 participants). This rose to 79.4% (95% CI: 71.8%-85.4%; 7 studies; n = 1,369) in the 2015-2019 period, and reached 86.9% (95% CI: 83.4%-89.9%; 1 study; n = 444) in the 2020+ period.

Patients with rifampicin-resistant tuberculosis achieved the highest success rate at 81.0% (95% CI: 72.9%-87.1%; n = 6 studies; n = 1,322). Patients from general population had the lowest success rate, at 51.7% (95% CI: 4.9%-95.7%; 3 studies; n = 578).

Geographically, the highest pooled estimate was observed in Multiregion studies (84.5%; 95% CI: 78.9%-88.8%; 6 studies; n = 1,082) and the lowest in the Centre region (18.0%; 95% CI: 0.1%-98.5%; 2 studies; n = 249).

Patients with mixed pulmonary and extrapulmonary tuberculosis achieved a success rate of 86.9% (95% CI: 83.4%-89.9%; 1 study; n = 444), compared to 72.6% (95% CI: 57.2%-83.9%; 12 studies; n = 1,702) among patients with pulmonary tuberculosis only. However, the mixed-group estimate should be interpreted with caution, as it is derived from a single study.

Patients receiving modified regimens comprising adapted treatment for fluoroquinolone resistance or second-line drug resistance achieved a pooled success rate of 87.2% (95% CI: 83.8% – 89.9%; 3 studies; n = 460). In contrast, patients receiving standard treatment regimens had a pooled success rate of 68.8% (95% CI: 52.2%-81.6%; 10 studies; n = 1,686) (**Table 3 and Figs. S11-16**).

**Table 3.** Subgroup meta-analysis of treatment success rate among drug-resistant tuberculosis patients in Cameroon, 1998–2022.

| Subgroup | Sample size | Prevalence<sup>1</sup> (%) | 95% CI limits<sup>1</sup> |  | Number of reports | Heterogeneity statistic<sup>1</sup> |  | Subgroup difference p-value |
| --- | --- | --- | --- | --- | --- | --- | --- | --- |
|  |  |  | Lower | Upper |  | I<sup>2</sup>(%) | p-value |  |
| **Study period (year)<sup>2</sup>** |  |  |  |  |  |  |  | 0.001 |
| └ 2005 | 150 | 60.6 | 27.5 | 86.2 | 2 | 93.9 | 0.001 |  |
| 2005-2009 | 26 | 46.2 | 26.6 | 66.6 | 1 | - | - |  |
| 2010-2014 | 157 | 19.8 | 0.0 | 99.5 | 2 | 0.0 | 0.999 |  |
| 2015-2019 | 1,369 | 79.4 | 71.8 | 85.4 | 7 | 86.1 | 0.001 |  |
| 2020+ | 444 | 86.9 | 83.4 | 89.9 | 1 | - | - |  |
| **Study design** |  |  |  |  |  |  |  | 0.660 |
| Cross-sectional | 195 | 64.6 | 57.5 | 71.3 | 1 | - | - |  |
| Cohort | 1,951 | 75.0 | 59.6 | 85.6 | 12 | 83.2 | 0.001 |  |
| **Region** |  |  |  |  |  |  |  | 0.001 |
| Centre | 249 | 18.0 | 0.1 | 98.5 | 2 | 0.0 | 0.999 |  |
| Littoral | 470 | 71.9 | 38.0 | 91.4 | 2 | 95.8 | 0.001 |  |
| North-West | 195 | 64.6 | 57.5 | 71.3 | 1 | - | - |  |
| West | 150 | 60.6 | 27.5 | 86.2 | 2 | 93.9 | 0.001 |  |
| Multi-region | 1,082 | 84.5 | 78.9 | 88.8 | 6 | 57.0 | 0.040 |  |
| **Participant** |  |  |  |  |  |  |  | 0.051 |
| Patient from general population | 578 | 51.7 | 4.9 | 95.7 | 3 | 52.9 | 0.119 |  |
| Retreated tuberculosis patient | 73 | 59.2 | 43.5 | 73.2 | 2 | 69.6 | 0.070 |  |
| Rifampicin-resistant tuberculosis | 1,322 | 81.0 | 72.9 | 87.1 | 6 | 87.3 | 0.001 |  |
| Multidrug-resistant tuberculosis | 173 | 68.9 | 24.1 | 93.9 | 2 | 96.6 | 0.001 |  |
| **Tuberculosis infection localization** |  |  |  |  |  |  |  | 0.014 |
| Pulmonary | 1,702 | 72.6 | 57.2 | 83.9 | 12 | 87.3 | 0.001 |  |
| Mixed | 444 | 86.9 | 83.4 | 89.9 | 1 | - | - |  |
| **Treatment regimen<sup>3</sup>** |  |  |  |  |  |  |  | 0.003 |
| Standard treatment | 1,686 | 68.8 | 52.2 | 81.6 | 10 | 89.6 | 0.001 |  |
| Other regimen | 460 | 87.2 | 83.8 | 89.9 | 3 | 0.0 | 0.967 |  |

1: Random effects model; 2: Time frame aligns with the development of successive National Strategic Plans against tuberculosis in Cameroon; CI: Confidence interval; Mixed included pulmonary and extrapulmonary tuberculosis cases; 3: Other regimen included fluoroquinolone resistant modified treatment regimen [FQR-Mtr] and second-line injectable-resistant modified treatment regimen [SLIR-Mtr]

### 3.5. Drug-resistant tuberculosis loss to follow-up

The overall pooled LTFU rate was 4.1% (95% CI: 2.8%-6.1%; 12 studies; n = 2,244 participants), with moderate heterogeneity (*I*² = 55.5%; *p* = 0.010) (**Fig. 4**). The overall pooled estimate was robust in the sensitivity analysis and no significant sign of publication bias was detected (**Figs. S17-18**).

**Fig. 4.**
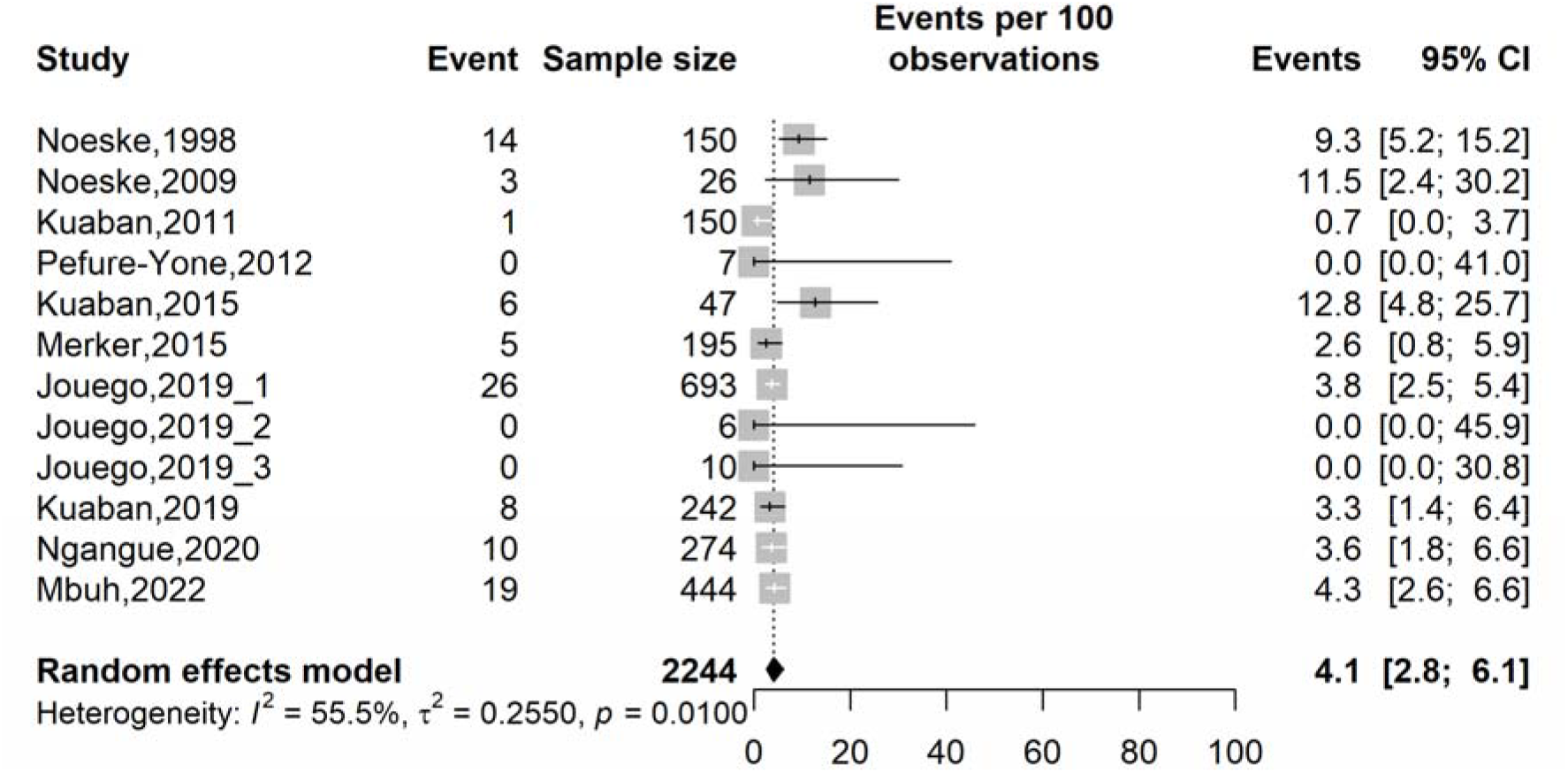
Forest plot of the pooled loss to follow-up rate among anti-tuberculosis drug-resistant patients in Cameroon, 1998-2022

Subgroup analyses revealed significant effect modification for participant subgroup (*p* = 0.00&), study period (*p* = 0.009) and region (*p* = 0.030). No significant effect modification was observed for tuberculosis infection localization (*p* = 0.879), study design (*p* = 0.354), and treatment regimen (*p* = 0.937).

Retreated tuberculosis patients had the highest LTFU rate at 12.3% (95% CI: 6.5%-22.0%, 2 studies; n = 73), which was significantly higher than patients from the general population at 5.0% (95% CI: 3.2%-7.8%, 4 studies; n = 875) and rifampicin-resistant TB patients at 3.4% (95% CI: 2.5%-4.6%, 5 studies; n = 1,146).

A significant temporal trend toward a decline in LTFU was observed. The earliest periods reported higher rates of 9.3% (95% CI: 5.2%-15.2%, 1 study; n = 150) for <2005 and 11.5% (95% CI: 2.4%-30.2%, 1 study; n = 26) for 2005-2009, while studies conducted after 2015 reported consistently lower rates of 3.8% (95% CI: 2.8%-5.0%, 6 studies; n = 1,193) for 2015-2019 and 4.0% (95% CI: 2.8%-5.8%, 2 studies; n = 718) for 2020+ (**Table 4** and Figs. S19-24).

**Table 4.** Subgroup meta-analysis of loss to follow-up rate among drug-resistant tuberculosis patients in Cameroon, 1998–2022.

| Subgroup | Sample size | Prevalence<sup>1</sup> (%) | 95% CI limits<sup>1</sup> |  | Number of studies | Heterogeneity statistic<sup>1</sup> |  | Subgroup difference p-value |
| --- | --- | --- | --- | --- | --- | --- | --- | --- |
|  |  |  | Lower | Upper |  | I<sup>2</sup>(%) | p-value |  |
| **Study period (year)<sup>2</sup>** |  |  |  |  |  |  |  | 0.009 |
| └ 2005 | 150 | 9.3 | 5.2 | 15.2 | 1 | - | - |  |
| 2005-2009 | 26 | 11.5 | 2.4 | 30.2 | 1 | - | - |  |
| 2010-2014 | 157 | 0.6 | 0.1 | 4.4 | 2 | 0.0 | 0.999 |  |
| 2015-2019 | 1,193 | 3.8 | 2.8 | 5.0 | 6 | 48.3 | 0.085 |  |
| 2020+ | 718 | 4.0 | 2.8 | 5.8 | 2 | 0.0 | 0.678 |  |
| **Study design** |  |  |  |  |  |  |  | 0.354 |
| Cross-sectional | 469 | 3.2 | 1.9 | 5.2 | 2 | 0.0 | 0.512 |  |
| Cohort | 1,775 | 4.5 | 2.7 | 7.3 | 10 | 58.6 | 0.010 |  |
| **Region** |  |  |  |  |  |  |  | 0.030 |
| Centre | 249 | 3.2 | 1.6 | 6.3 | 2 | 0.0 | 0.010 |  |
| Littoral | 470 | 4.7 | 3.1 | 7.0 | 2 | 62.3 | 0.103 |  |
| North-West | 195 | 2.6 | 0.8 | 5.9 | 1 | - | - |  |
| West | 150 | 9.3 | 5.2 | 15.2 | 1 | - | - |  |
| Multi-region | 1,180 | 3.4 | 1.5 | 7.3 | 6 | 57.1 | 0.040 |  |
| **Participant** |  |  |  |  |  |  |  | 0.001 |
| Patient from general population | 875 | 5.0 | 3.2 | 7.8 | 4 | 57.7 | 0.069 |  |
| Retreated tuberculosis patient | 73 | 12.3 | 6.5 | 22.0 | 2 | 0.0 | 0.879 |  |
| Rifampicin-resistant tuberculosis | 1,146 | 3.4 | 2.5 | 4.6 | 5 | 0.0 | 0.890 |  |
| Multidrug-resistant tuberculosis | 150 | 0.7 | 0.0 | 3.7 | 1 | - | - |  |
| **Tuberculosis infection localization** |  |  |  |  |  |  |  | 0.879 |
| Pulmonary | 1,800 | 4.1 | 2.5 | 6.5 | 11 | 59.2 | 0.006 |  |
| Mixed | 444 | 4.3 | 2.6 | 6.6 | 1 | - | - |  |
| **Treatment regimen<sup>3</sup>** |  |  |  |  |  |  |  | 0.937 |
| Standard treatment | 1,784 | 4.2 | 2.6 | 6.9 | 9 | 67.4 | 0.002 |  |
| Other regimen | 460 | 4.1 | 2.6 | 6.4 | 3 | 0.0 | 1.000 |  |

1: Random effects model; 2: Time frame aligns with the development of successive National Strategic Plans against tuberculosis in Cameroon; CI: Confidence interval; Mixed included pulmonary and extrapulmonary tuberculosis cases; 3: Other regimen included fluoroquinolone resistant modified treatment regimen [FQR-Mtr] and second-line injectable-resistant modified treatment regimen [SLIr-Mtr]

### 3.6. Drug-resistant tuberculosis treatment failure

In this meta-analysis of 12 studies evaluating treatment failure rates among patients with drug-resistant tuberculosis in Cameroon (1998–2022), the individual study estimates varied widely, ranging from 0.0% to 71.4%. The pooled failure rate was 5.0% (95% CI: 1.1–19.8), but this summary estimate should be interpreted with caution due to substantial heterogeneity (*I*² = 93.2%, *p* < 0.0001), indicating that the true effect is not consistent across settings (**Fig. 5**). The overall pooled failure rate was robust at sensitivity analysis, and no significant sign of publication bias was detected (**Figs. S25-26**).

**Fig. 5.**
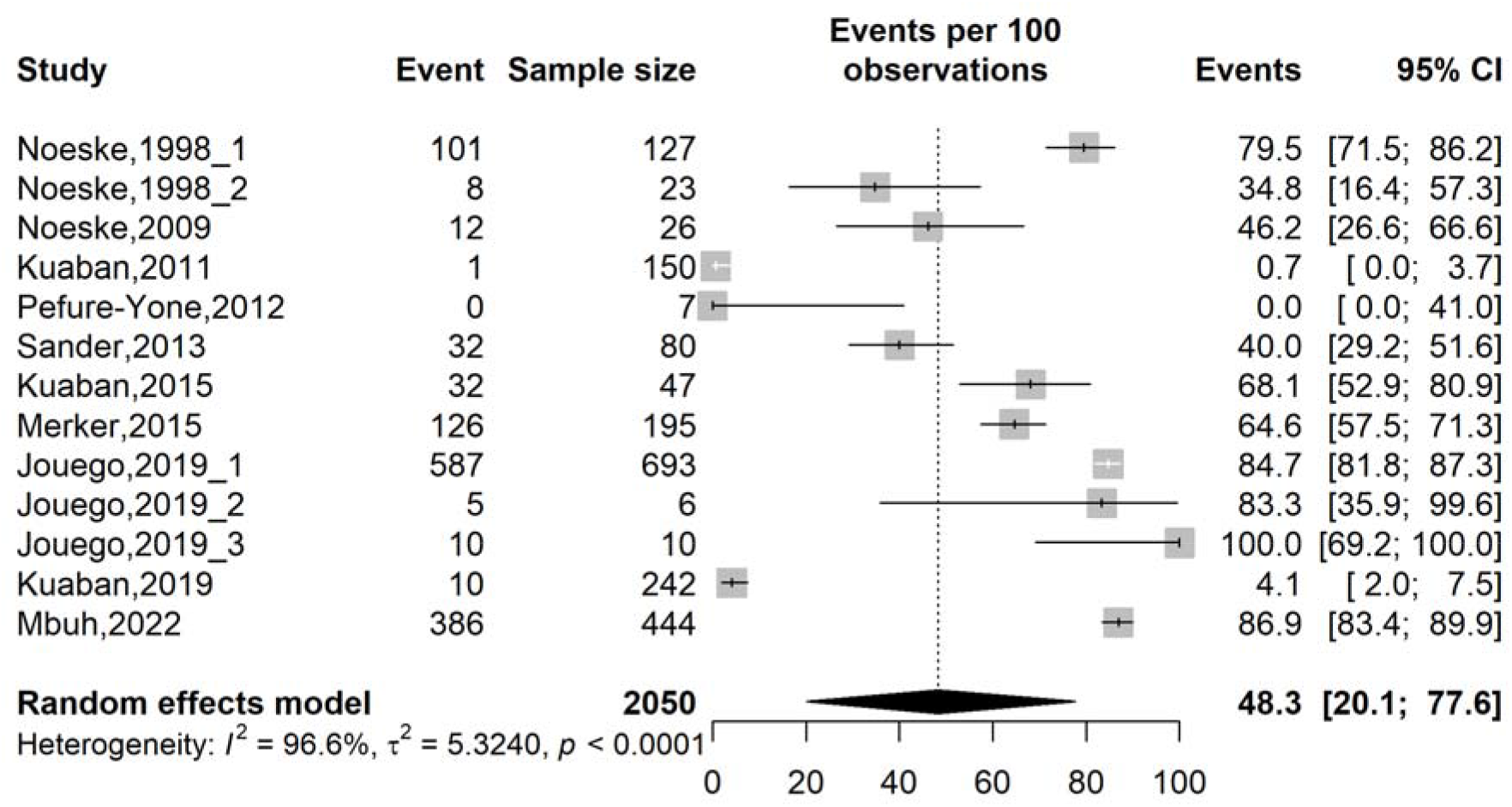
Forest plot of the pooled treatment failure rate among anti-tuberculosis drug-resistant patients in Cameroon, 1998-2022

The subgroup meta-analysis examining factors influencing treatment failure rates among patients with drug-resistant tuberculosis in Cameroon, revealed significant effect modification by study period, design, region, and tuberculosis infection localization, and treatment regimen (all p-values for subgroup differences ≤0.001). The temporal pattern was marked by a significant decline in failure rates from 60.6% (95% CI: 27.5–86.2; 2 studies; n = 150) in 2005 to 0.6% (95% CI: 0.1–4.4; 6 studies; n = 1,193) in 2015–2019 and 1.4% (95% CI: 0.5–2.9; 1 study, n = 444) from 2020 onwards, coinciding with the implementation of successive National Strategic Plans against tuberculosis. Treatment regimen was also associate with failure rates heterogeneity. Other regiment exhibiting lower failure rates (31.6%; 95% CI: 10.6-64.3; 10 studies, n = 1,590) compared to standard treatment for DR-TB (87.2%; 95% CI: 83.8-89.9; 3 studies, n = 460) (**Table 5 and Figs. S27-33**).

**Table 5.** Subgroup meta-analysis of failure rate among drug-resistant tuberculosis patients in Cameroon, 1998–2022.

| Subgroup | Sample size | Prevalence<sup>1</sup> (%) | 95% CI limits<sup>1</sup> | Number of | Heterogeneity statistic<sup>1</sup> | Subgroup difference |
| --- | --- | --- | --- | --- | --- | --- |

| | | | Lower | Upper | studies | $I^2$(%) | p-value | p-value |
| --- | --- | --- | --- | --- | --- | --- | --- | --- |
| **Study period (year)<sup>2</sup>** |  |  |  |  |  |  |  | 0.001 |
| ≤ 2005 | 150 | 60.6 | 27.5 | 86.2 | 2 | 93.9 | 0.001 |  |
| 2005-2009 | 26 | 46.2 | 26.6 | 66.6 | 1 | - | - |  |
| 2010-2014 | 237 | 3.3 | 0.1 | 48.4 | 3 | 90.0 | 0.001 |  |
| 2015-2019 | 1,193 | 0.6 | 0.1 | 4.4 | 6 | 97.7 | 0.001 |  |
| 2020+ | 444 | 1.4 | 0.5 | 2.9 | 1 | - | - |  |
| **Study design** |  |  |  |  |  |  |  | 0.001 |
| Cross-sectional | 195 | 64.6 | 57.5 | 71.3 | 1 | - | - |  |
| Case-control | 80 | 40.0 | 29.2 | 51.6 | 1 | - | - |  |
| Cohort | 1,775 | 47.4 | 15.1 | 82.0 | 11 | 96.6 | 0.001 |  |
| **Region** |  |  |  |  |  |  |  | 0.001 |
| Centre | 249 | 4.0 | 2.2 | 7.3 | 2 | 0.0 | 0.999 |  |
| Littoral | 470 | 71.9 | 38.0 | 91.4 | 2 | 95.8 | 0.001 |  |
| North-West | 275 | 53.2 | 36.0 | 69.6 | 2 | 92.7 | 0.001 |  |
| West | 150 | 60.6 | 27.7 | 86.2 | 2 | 92.1 | 0.001 |  |
| Multi-region | 906 | 68.4 | 9.3 | 97.9 | 5 | 92.2 | 0.001 |  |
| **Participant** |  |  |  |  |  |  |  | 0.306 |
| Patient from general population | 578 | 51.7 | 4.9 | 95.7 | 3 | 52.9 | 0.1194 |  |
| Retreated tuberculosis patient | 153 | 51.3 | 36.7 | 65.7 | 3 | 78.1 | 0.010 |  |
| Rifampicin-resistant tuberculosis | 1,146 | 71.7 | 20.6 | 96.1 | 5 | 98.2 | 0.001 |  |
| Multidrug-resistant tuberculosis | 173 | 5.5 | 0.2 | 59.9 | 2 | 93.7 | 0.001 |  |
| **Tuberculosis infection localization** |  |  |  |  |  |  |  | 0.003 |
| Pulmonary | 1,606 | 43.8 | 16.3 | 75.8 | 12 | 96.5 | 0.001 |  |
| Mixed | 444 | 86.9 | 83.4 | 89.9 | 1 | - | - |  |
| **Treatment regimen<sup>3</sup>** |  |  |  |  |  |  |  | 0.001 |
| Standard treatment | 1,590 | 31.6 | 10.6 | 64.3 | 10 | 97.1 | 0.001 |  |
| Other regimen | 460 | 87.2 | 83.8 | 89.9 | 3 | 0.0 | 0.967 |  |

1: Random effects model; 2: Time frame aligns with the development of successive National Strategic Plans against tuberculosis in Cameroon; CI: Confidence interval; Mixed included pulmonary and extrapulmonary tuberculosis cases; 3: Other regimen included fluoroquinolone-resistant modified treatment regimen [FQR-Mtr] and second-line injectable-resistant modified treatment regimen [SLIr-Mtr]

### 3.7. Prevalence of adverse drug events

The pooled prevalence of ADEs among patients treated for multidrug-resistant tuberculosis in Cameroon was 70.8% (95% CI: 40.2–89.7; *I*² = 96.1%; 3 studies; n = 251), indicating that the majority of patients experienced at least one ADE during treatment. Among specific ADE types, ototoxicity and gastro-intestinal disorders were the most frequently reported, with pooled prevalences of 41.9% (95% CI: 23.7–62.6; *I*² = 93.2%; 3 studies; n = 251) and 40.9% (95% CI: 25.5–58.2; *I*² = 89.5%; 2 studies; n = 172), respectively. The pooled prevalence of nephrotoxicity and neurological disorders showed moderate heterogeneity (*I*² ≤ 50%). Substantial heterogeneity was observed for most ADE types, particularly for the overall prevalence; ototoxicity, gastrointestinal disorders, and hepatotoxicity (*I*² ≥ 80%) (**Figs. 6 and S33**).

**Fig. 6.**
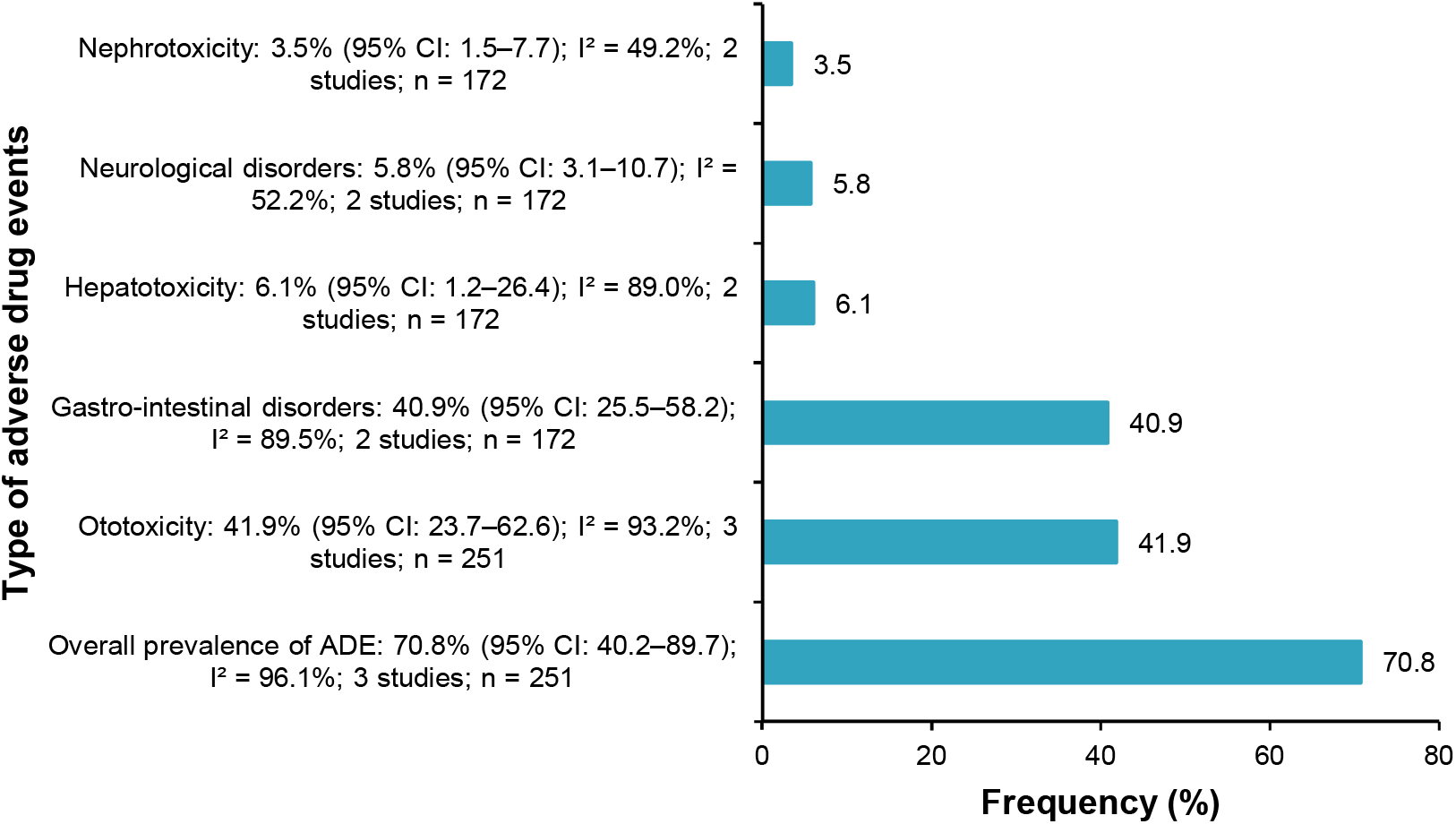
Trend of adverse drug events (ADE) among patients treated for multidrug-resistant tuberculosis in Cameroon, 2015–2019

### 3.8. Factors associated with unfavorable treatment outcomes

#### Meta-analysis

HIV-positive status was identified as a significant predictor of unfavorable treatment outcomes among drug-resistant tuberculosis patients in Cameroon, with a pooled odds ratio of 2.76 (95% CI: 1.95–3.93; 6 studies), indicating that HIV-coinfected patients had three times the odds of experiencing an unfavorable outcome compared to HIV-negative patients. The gender was also significantly associated with unfavorable outcome. Male gender was associated with 73% increased odds of unfavorable treatment outcomes following DR-TB management (OR: 1.73; 95% CI: 1.25-2.40; 5 studies) (**Table 6 and Figs.**

**Table 6.**
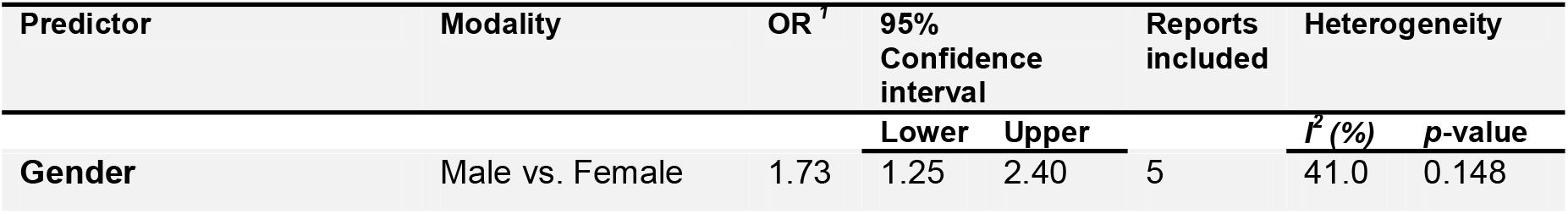
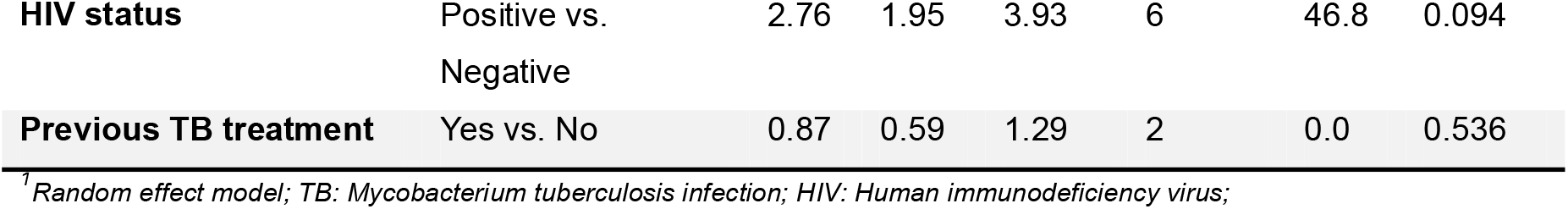
Predictors of unfavorable outcome among patients treated for drug-resistant tuberculosis in Cameroon, 1995–2022.

| Predictor | Modality | OR<sup>1</sup> | 95% Confidence interval |  | Reports included | Heterogeneity |  |
| --- | --- | --- | --- | --- | --- | --- | --- |
| | | | Lower | Upper | | $I^2$ (%) | p-value |
| Gender | Male vs. Female | 1.73 | 1.25 | 2.40 | 5 | 41.0 | 0.148 |
| **HIV status** | Positive vs. Negative | 2.76 | 1.95 | 3.93 | 6 | 46.8 | 0.094 |
| **Previous TB treatment** | Yes vs. No | 0.87 | 0.59 | 1.29 | 2 | 0.0 | 0.536 |

<sup>†</sup>Random effect model; TB: *Mycobacterium tuberculosis* infection; HIV: Human immunodeficiency virus;

#### Narrative review

Several additional predictors of unfavorable treatment outcomes were reported in individual studies but could not be included in the meta-analysis because they were not consistently evaluated across studies. These findings should therefore be interpreted as evidence from individual studies rather than pooled estimates. Among these, multidrug-resistant tuberculosis (MDR-TB) was associated with the highest odds of unfavorable treatment outcomes compared with non-MDR-TB (OR = 11.47, 95% CI: 3.12–42.12; *p* < 0.001)[22]. A history of previous DR-TB treatment was associated with a nearly 4-fold increased risk of poor outcomes (adjusted relative risk [aRR] = 3.9, 95% CI: 1.3–12.0, *p* = 0.02); however, this finding was based on a very small sample (n = 9) and should be interpreted with caution [34]. Missing baseline second-line drug susceptibility testing (DST) results were associated with a 65% higher mortality risk in RR-TB patients (adjusted sub-hazard ratio [aSHR] = 1.65, 95% CI: 1.04–2.60, *p* = 0.03). This finding was based on a very small sample (n = 9) and should be interpreted with caution [31]. Moderate to severe anemia (hemoglobin ≤ 10 g/dL) independently predicted unfavorable outcomes in RR-TB patients (OR: 2.87, 95% CI: 1.25–6.58, *p* = 0.013) [30]. Underweight (body mass index [BMI] < 18.5 kg/m²) significantly increased the risk of unfavorable outcome (adjusted hazard ratio [AHR]: 1.8, 95% CI: 1.4–2.5) [27]. Fluoroquinolone resistance was a strong predictor of bacteriological failure or relapse (AHR: 6.8, 95% CI: 3.5–13.2), while isoniazid resistance was also associated with an increased risk of failure/relapse (AHR: 9.4, 95% CI: 1.3–68.0) [27]. Extensive lung lesions (≥ 50% of lung zones affected) were independently associated with higher mortality risk (AHR: 1.6, 95% CI: 1.06–2.3) [27]. Additionally, age > 40 years was associated with a 62% higher risk of overall mortality among RR-TB patients (aSHR: 1.62, 95% CI: 1.01–2.60, *p* = 0.045) [31].

## 4. Discussion

This systematic review and meta-analysis provide the first comprehensive synthesis of treatment outcomes among patients with DR-TB in Cameroon. In our review, the combined treatment success rate was 74.2%. Mortality, loss to follow-up, and treatment failure rates were 6.8%, 4.1%, and 5.0%, respectively. ADEs were common, affecting about 70.8% of patients. The most frequent complications were ototoxicity and gastrointestinal problems. HIV-positive status and male sex were significantly associated with unfavorable treatment outcomes. Although treatment success has improved over time and negative outcomes have decreased, there were still substantial differences between studies. These differences likely reflect variations in patient groups, treatments, healthcare settings, and program periods.

Our review found that approximately 74.2% of patients with DR-TB in Cameroon achieved successful treatment outcomes, indicating that nearly three-quarters of patients completed treatment successfully. This treatment success proportion is higher than the recent global estimate of 68% among patients with MDR/RR-TB reported by the World Health Organization (WHO)[37], and is comparable to the 74.6% reported in a systematic review and meta-analysis from Central and West Africa [38].The similarity between our findings and those reported in Central and West Africa may reflect comparable epidemiological characteristics, treatment strategies, and programmatic approaches adopted across countries in the region. Notably, treatment success improved from approximately 61% in studies conducted before 2005 to nearly 87% in studies conducted from 2020 onwards, suggesting substantial improvements in DR-TB programme performance over successive implementation periods.

Over time, improvements such as better diagnostic tools, expanded rapid molecular and drug-susceptibility testing, more decentralized care, and stronger patient monitoring have likely helped increase treatment success. These changes align with global trends, in which treatment success has gradually improved since the introduction of WHO-recommended shorter regimens and patient-centered care models [37, 39]. Although patients receiving modified regimens achieved higher treatment success than those receiving standard regimens, this finding should be interpreted with caution, as it is based on a small number of observational studies. Likewise, the substantial between-study heterogeneity indicates that treatment outcomes varied across settings and patient populations.

In our review, we observed a pooled mortality proportion of 6.8%, which indicates that death remains an important unfavorable outcome among patients with DR-TB in Cameroon. This pooled mortality is lower than estimates reported in systematic reviews from Central and West Africa and among HIV co-infected MDR-TB patients in sub-Saharan Africa [38, 40, 41]. However, these comparisons should be interpreted with caution due to differences in study populations, treatment regimens, and healthcare settings. The relatively low mortality observed in Cameroon may reflect improvements in early diagnosis, wider access to drug-susceptibility testing, expansion of specialized DR-TB treatment centers, and strengthened programmatic management under the National Tuberculosis Program. In addition, the progressive integration of TB and HIV services and improved access to antiretroviral therapy may have contributed to better survival among co-infected patients. Nevertheless, mortality remains an important challenge, particularly among patients with HIV co-infection and severe anemia, both of which have been consistently associated with unfavorable treatment outcomes in Cameroon and other settings [28, 40, 41]. These findings underscore the importance of strengthening early case detection, integrating TB screening in HIV management, and ensuring prompt initiation of effective treatment to further reduce mortality among patients with DR-TB in Cameroon.

The pooled LTFU proportion of 4.1% in our review indicates relatively good retention among patients receiving DR-TB treatment in Cameroon. Moreover, LTFU declined over time, from 9.3% before 2005 to approximately 4% in studies conducted after 2015. This trend likely reflects improvements in patient follow-up and the progressive strengthening of DR-TB services under the National Tuberculosis Program. However, retreated tuberculosis patients had a substantially higher LTFU than other patient groups, suggesting that previous treatment history remains an important barrier to treatment completion. Similar observations have been reported in previous studies, where treatment fatigue, prolonged treatment duration, socioeconomic challenges, and previous negative treatment experiences contributed to poor adherence among previously treated patients [41, 42]. These findings highlight the importance of providing targeted adherence support for patients with a history of previous tuberculosis treatment.

Similarly, the pooled treatment failure proportion of 5.0% was relatively low and declined markedly over successive study periods. The reduction in treatment failure probably reflects improvements in diagnosis, treatment monitoring, and the implementation of more effective treatment regimens. However, treatment failure remained an important unfavorable outcome, particularly in earlier study periods, highlighting the need for continued efforts to optimize DR-TB management. These findings underscore the importance of expanding drug susceptibility testing and ensuring timely initiation of appropriate treatment to further reduce treatment failure among patients with DR-TB in Cameroon, in line with current WHO recommendations [43].

The pooled prevalence of ADEs of 70.8% in this review indicates that treatment-related toxicity remains a major challenge in the management of DR-TB in Cameroon. Ototoxicity and gastrointestinal disorders were the most frequently reported adverse events, consistent with previous reports from Cameroon and the known toxicity profile of second-line anti-tuberculosis drugs [35, 36]. These adverse events may compromise treatment adherence and contribute to unfavorable treatment outcomes. The high burden of ADEs observed in this review further supports the WHO recommendation to transition from injectable-containing regimens to safer all-oral regimens whenever appropriate [2].

HIV-positive patients had nearly threefold higher odds of unfavorable treatment outcomes, while male patients had 73% higher odds of unfavorable outcomes than females. These findings are consistent with previous systematic reviews and meta-analyses, which have shown that HIV co-infection and male sex are associated with poorer DR-TB treatment outcomes[40, 44]. HIV may adversely affect treatment outcomes through immunosuppression and increased susceptibility to opportunistic infections, while poorer outcomes among men have been attributed to delayed healthcare seeking, lower treatment adherence, and a higher prevalence of behavioral risk factors such as smoking and alcohol use [40, 44].These findings highlight the importance of strengthening integrated TB/HIV services and ensuring rigorous treatment follow-up for TB/HIV-coinfected patients to improve treatment outcomes among those with DR-TB.

The findings of this review have important implications for DR-TB management in Cameroon. Although treatment outcomes have improved over time, sustaining this progress will require ongoing investment in rapid drug susceptibility testing, integrated TB/HIV services, patient-centered adherence support, and pharmacovigilance. Strengthening these components of the National Tuberculosis Program and scaling up WHO-recommended treatment strategies could further improve outcomes and accelerate progress toward the End TB Strategy goals [45].

## 5. Limitations

Several limitations should be acknowledged in this review. First, the predominance of observational studies restricts the ability to draw causal inferences. Second, considerable heterogeneity was identified across several pooled outcomes indicating that the pooled estimates should be interpreted with caution and considered as average measures of the true effect. Third, certain analyses, particularly those concerning adverse drug events and predictors of unfavorable outcomes, relied on a limited number of studies. Additionally, the included studies did not represent all regions of Cameroon, as most of the evidence came from only a few regions. As a result, the pooled estimates may not comprehensively reflect the national epidemiology and treatment outcomes of DR-TB. Despite these limitations, this review presents the first comprehensive synthesis of DR-TB treatment outcomes in Cameroon and provides valuable evidence to guide policy, clinical practice, and future research.

## 6. Conclusions

This systematic review and meta-analysis is the first to synthesize data on treatment outcomes among patients with DR-TB in Cameroon. Treatment success has increased over time, and rates of death, loss to follow-up, and treatment failure have stayed relatively low. Still, adverse drug events were common, and people living with HIV had worse treatment outcomes. These results show progress in managing DR-TB in Cameroon but also highlight the need for ongoing efforts to improve outcomes. Improving rapid drug susceptibility testing, integrating TB and HIV services, focusing on patient-centered care, and monitoring drug safety will be important for better DR-TB management and for reaching the WHO End TB Strategy goals.

## Abbreviations

*ADEs*: Adverse Drug Events
*AHR*: Adjusted Hazard Ratio
*AJOL*: African Journals Online
*aRR*: Adjusted Relative Risk
*aSHR*: Adjusted Sub-Hazard Ratio
*BMI*: Body Mass Index
*CE*: Centre Region (Cameroon)
*CI*: Confidence Interval
*DR-TB*: Drug-Resistant Tuberculosis
*DST*: Drug Susceptibility Testing
*DTC*: Diagnostic and Treatment Center
*EPTB*: Extra-Pulmonary Tuberculosis
*FQR-Mtr*: Fluoroquinolone-Resistant Modified Treatment Regimen
*GLMM*: Generalized Linear Mixed Models
*HIV*: Human Immunodeficiency Virus
*JBI*: Joanna Briggs Institute
*LT*: Littoral Region (Cameroon)
*LTFU*: Lost to Follow-Up
*MDR-TB*: Multidrug-Resistant Tuberculosis
*MeSH*: Medical Subject Headings
*N/A*: Not Applicable
*NR*: Not Reported
*NW*: North-West Region (Cameroon)
*OR*: Odds Ratio
*PLOGIT*: Probit-Logit Transformation
*PRISMA*: Preferred Reporting Items for Systematic Reviews and Meta-Analyses
*PROSPERO*: International Prospective Register of Systematic Reviews
*PTB*: Pulmonary Tuberculosis
*Ref*: Reference
*RR-TB*: Rifampicin-Resistant Tuberculosis
*SLIr-Mtr*: Second-Line Injectable-Resistant Modified Treatment Regimen
*SW*: South-West Region (Cameroon)
*TB*: Tuberculosis
*WE*: West Region (Cameroon)
*WHO*: World Health Organization

## Supplementary Information

Supplementary Material

## Data Availability

All data generated in this study are available in the main manuscript and its supplementary material.

## Acknowledgments

Not applicable.

## Authors’ contributions

FZLC conceived the original idea of the study; FZLC and RT conducted the literature search; FZLC, RT, ADT, CA, and MNT selected the studies, extracted the relevant information, critically assessed included reports and synthesized the data; FZLC performed the analyses; FZLC, ADT, RT, and CA wrote the first draft of the manuscript. FZLC, ADT, CA, RT, RKNO, CJKM, and MNT critically reviewed and revised successive drafts of the manuscript. FZLC, ADT, CA, RT, RKNO, CJKM, and MNT read and approved the final version of the manuscript.

## Declarations

### Ethical approval and consent to participate

Not applicable.

### Consent for publication

Not applicable.

### Clinical trial number

Not applicable.

## References

1. WHO. Tuberculosis (TB). WHO | Regional Office for Africa, Brazzaville. 2026. https://www.afro.who.int/health-topics/tuberculosis-tb. Accessed 7 July 2026.

2. WHO. WHO consolidated guidelines on tuberculosis. Module 4: treatment - drug-resistant tuberculosis treatment, 2022 update. WHO, Geneva. 2022. https://www.who.int/publications/i/item/9789240063129. Accessed 7 July 2026.

3. Global, regional, and national age-specific progress towards the 2020 milestones of the WHO End TB Strategy: a systematic analysis for the Global Burden of Disease Study 2021. Lancet Infect Dis. 2024;24:698–725. 10.1016/S1473-3099(24)00007-0.

4. WHO. Global Tuberculosis Report 2023. WHO, Geneva. 2026. https://www.who.int/teams/global-programme-on-tuberculosis-and-lung-health/tb-reports/global-tuberculosis-report-2023. Accessed 7 July 2026.

5. Nyasulu PS, Doumbia CO, Ngah V, Togo ACG, Diarra B, Chongwe G. Multidrug-resistant tuberculosis: latest opinions on epidemiology, rapid diagnosis and management. Curr Opin Pulm Med. 2024;30:217–28. 10.1097/MCP.0000000000001070.

6. Noeske J, Yakam AN, Foe JLA, Nguafack D, Kuaban C. Rifampicin resistance in new bacteriologically confirmed pulmonary tuberculosis patients in Cameroon: a cross-sectional survey. BMC Res Notes. 2018;11:580. 10.1186/s13104-018-3675-0.

7. Drug Resistance Profiles of Mycobacterium tuberculosis Complex and Factors Associated with Drug Resistance in the Northwest and Southwest Regions of Cameroon | PLOS One. https://journals.plos.org/plosone/article?id=10.1371/journal.pone.0077410. Accessed 20 May 2026.

8. Falzon D, Gandhi N, Migliori GB, Sotgiu G, Cox H, Holtz TH, et al. Resistance to fluoroquinolones and second-line injectable drugs: impact on MDR-TB outcomes. Eur Respir J. 2013;42:156–68. 10.1183/09031936.00134712.

9. Kuaban C, Noeske J, Rieder HL, Aït-Khaled N, Abena Foe JL, Trébucq A. High effectiveness of a 12-month regimen for MDR-TB patients in Cameroon. Int J Tuberc Lung Dis. 2015;19:517–24. 10.5588/ijtld.14.0535.

10. Kuaban A, Balkissou AD, Ekongolo MCE, Nsounfon AW, Pefura-Yone EW, Kuaban C. Incidence and factors associated with unfavourable treatment outcome among patients with rifampicin-resistant pulmonary tuberculosis in Yaoundé, Cameroon. Pan Afr Med J. 2021;38:229. 10.11604/pamj.2021.38.229.28317.

11. Mbuh TP, Wandji A, Keugni L, Mboh S, Ane-Anyangwe I, Mbacham WF, et al. Predictors of Drug-Resistant Tuberculosis among High-Risk Population Diagnosed under National Program Conditions in the Littoral Region, Cameroon. Biomed Res Int. 2021;2021:8817442. 10.1155/2021/8817442.

12. Page MJ, McKenzie JE, Bossuyt PM, Boutron I, Hoffmann TC, Mulrow CD, et al. The PRISMA 2020 statement: an updated guideline for reporting systematic reviews. BMJ. 2021;372:n71. 10.1136/bmj.n71.

13. WHO. Outcome definitions. WHO TB Knowledge Sharing Platform. 2026. https://tbksp.who.int/en/node/3027. Accessed 28 June 2026.

14. Kaapu KG, Rukasha I. Trends and treatment outcomes of drug resistant tuberculosis in Limpopo Province, South Africa (2011–2019): A Retrospective Study. PLoS One. 2025;20:e0335600. 10.1371/journal.pone.0335600.

15. Poka-Mayap V, Dombu-Guiafaing RC, Balkissou AD, Mangamba L-ME, Kuaban A, Nsounfon AW, et al. Trends and determinants of unfavourable outcomes in paediatric tuberculosis: insights from a 20-year cohort in Cameroon. BMJ Open Resp Res. 2025;12. 10.1136/bmjresp-2025-003292.

16. Vadakunnel MJ, Nehru VJ, Brammacharry U, Ramachandra V, Palavesam S, Muthukumar A, et al. Factors associated with unfavourable treatment outcomes among patients with Multidrug-resistant Tuberculosis receiving outpatients care. Sci Rep. 2025;15:28335. 10.1038/s41598-025-13227-5.

17. JBI. JBI Critical Appraisal Tools | JBI. JBI. 2026. https://jbi.global/critical-appraisal-tools. Accessed 29 Mar 2026.

18. Stijnen T, Hamza TH, Ozdemir P. Random effects meta-analysis of event outcome in the framework of the generalized linear mixed model with applications in sparse data. Stat Med. 2010;29:3046–67. 10.1002/sim.4040.

19. R Core Team. R: A Language and Environment for Statistical Computing. R Foundation for Statistical Computing,Vienna, Austria. 2024. https://www.R-project.org/. Accessed 30 May 2024.

20. Egger M, Davey Smith G, Schneider M, Minder C. Bias in meta-analysis detected by a simple, graphical test. BMJ (Clinical research ed). 1997;315:629–34. 10.1136/bmj.315.7109.629.

21. Begg CB, Mazumdar M. Operating characteristics of a rank correlation test for publication bias. Biometrics. 1994;50:1088.

22. Noeske J, Nguenko PN. Impact of resistance to anti-tuberculosis drugs on treatment outcome using World Health Organization standard regimens. Transactions of the Royal Society of Tropical Medicine and Hygiene. 2002;96:429–33. 10.1016/S0035-9203(02)90383-4.

23. Noeske J, Voelz N, Fon E, Abena Foe J-L. Early results of systematic drug susceptibility testing in pulmonary tuberculosis retreatment cases in Cameroon. BMC Res Notes. 2012;5:160. 10.1186/1756-0500-5-160.

24. Kuaban C, Noeske J, Rieder HL, Aït-Khaled N, Abena Foe JL, Trébucq A. High effectiveness of a 12-month regimen for MDR-TB patients in Cameroon. int j tuberc lung dis. 2015;19:517–24. 10.5588/ijtld.14.0535.

25. Pefura-Yone EW, Kengne AP, Kuaban C. Non-conversion of sputum culture among patients with smear positive pulmonary tuberculosis in Cameroon: a prospective cohort study. BMC Infect Dis. 2014;14:138. 10.1186/1471-2334-14-138.

26. Sander MS, Vuchas CY, Numfor HN, Nsimen AN, Abena J-LF, Noeske J, et al. Sputum bacterial load predicts multidrug-resistant tuberculosis in retreatment patients: a case-control study. int j tuberc lung dis. 2016;20:793–9. 10.5588/ijtld.15.0259.

27. Schwœbel V, Trébucq A, Kashongwe Z, Bakayoko AS, Kuaban C, Noeske J, et al. Outcomes of a nine-month regimen for rifampicin-resistant tuberculosis up to 24 months after treatment completion in nine African countries. eClinicalMedicine. 2020;20. 10.1016/j.eclinm.2020.100268.

28. Kuaban C, Toukam LDI, Sander M. Treatment outcomes and factors associated with unfavourable outcome among previously treated tuberculosis patients with isoniazid resistance in four regions of Cameroon. Pan Afr Med J. 2020;37:1–12. 10.11604/pamj.2020.37.45.25684.

29. Merker M, Egbe NF, Ngangue YR, Vuchas C, Kohl TA, Dreyer V, et al. Transmission patterns of rifampicin resistant *Mycobacterium tuberculosis* complex strains in Cameroon: a genomic epidemiological study. BMC Infect Dis. 2021;21:891. 10.1186/s12879-021-06593-8.

30. Kuaban A, Balkissou AD, Essadi Ekongolo MC, Nsounfon AW, Pefura-Yone EW, Kuaban C. Incidence and factors associated with unfavourable treatment outcome among patients with rifampicin-resistant pulmonary tuberculosis in Yaounde, Cameroon. Pan Afr Med J. 2021;38. 10.11604/pamj.2021.38.229.28317.

31. Jouego CG, Gils T, Piubello A, Mbassa V, Kuate A, Ngono A, et al. Programmatic management of rifampicin-resistant tuberculosis with standard regimen in Cameroon: a retrospective cohort study. International Journal of Infectious Diseases. 2022;124:81–8. 10.1016/j.ijid.2022.09.012.

32. Mpoh M, Deli V, Daniel T, Salvo F. Safety of antituberculosis agents used for multidrug-resistant tuberculosis among patients attending the Jamot Hospital of Yaounde, Cameroon. Int J Mycobacteriol. 2023;12:168. 10.4103/ijmy.ijmy_88_23.

33. Ngangue YR, Mbuli C, Neh A, Nshom E, Koudjou A, Palmer D, et al. Diagnostic Accuracy of the Truenat MTB Plus Assay and Comparison with the Xpert MTB/RIF Assay to Detect Tuberculosis among Hospital Outpatients in Cameroon. J Clin Microbiol. 2022;60:e00155–22. 10.1128/jcm.00155-22.

34. Mbuh TP, Mendjime P, Goupeyou-Wandji I-A, Donkeng-Donfack VF, Kahou J, Endale Mangamba L-M, et al. Trends of drug-resistant tuberculosis and risk factors to poor treatment-outcome: a database analysis in Littoral region-Cameroon, 2013–2022. BMC Public Health. 2024;24:3195. 10.1186/s12889-024-20585-8.

35. Mbuh TP, Meriki HD, Thumamo Pokam BD, Adeline W, Enoka F, Ghislain T, et al. Incidence of Adverse Drug Events among Patients on Second Line Anti-Tuberculosis Regimen in the Littoral Region of Cameroon. The International Journal of Mycobacteriology. 2021;10:463–8. 10.4103/ijmy.ijmy_160_21.

36. Poka-Mayap V, Balkissou Adamou D, Pefura-Yone EW, Kuaban C. Ototoxicité liée à la kanamycine au cours du traitement de la tuberculose multirésistante. Revue des Maladies Respiratoires. 2020;37:369–75. 10.1016/j.rmr.2019.12.005.

37. Chen Z, Wang T, Du J, Sun L, Wang G, Ni R, et al. Decoding the WHO Global Tuberculosis Report 2024: A Critical Analysis of Global and Chinese Key Data. Zoonoses. 2025;5:999. 10.15212/ZOONOSES-2024-0061.

38. Toft AL, Dahl VN, Sifna A, Ige OM, Schwoebel V, Souleymane MB, et al. Treatment outcomes for multidrug- and rifampicin-resistant tuberculosis in Central and West Africa: a systematic review and meta-analysis. Int J Infect Dis. 2022;124 Suppl 1:S107–16. 10.1016/j.ijid.2022.08.015.

39. Tiberi S, Utjesanovic N, Galvin J, Centis R, D’Ambrosio L, van den Boom M, et al. Drug resistant TB - latest developments in epidemiology, diagnostics and management. Int J Infect Dis. 2022;124 Suppl 1:S20–5. 10.1016/j.ijid.2022.03.026.

40. Chem ED, Van Hout MC, Hope V. Treatment outcomes and antiretroviral uptake in multidrug-resistant tuberculosis and HIV co-infected patients in Sub Saharan Africa: a systematic review and meta-analysis. BMC Infect Dis. 2019;19:723. 10.1186/s12879-019-4317-4.

41. Teferi MY, El-Khatib Z, Boltena MT, Andualem AT, Asamoah BO, Biru M, et al. Tuberculosis Treatment Outcome and Predictors in Africa: A Systematic Review and Meta-Analysis. Int J Environ Res Public Health. 2021;18:10678. 10.3390/ijerph182010678.

42. Walker EF, Flook M, Rodger AJ, Fielding KL, Stagg HR. Quantifying non-adherence to anti-tuberculosis treatment due to early discontinuation: a systematic literature review of timings to loss to follow-up. BMJ Open Resp Res. 2024;11. 10.1136/bmjresp-2023-001894.

43. Lee H, Kim J, Kim J, Park Y-J. [Review of the Global Burden of Tuberculosis in 2023: Insights from the WHO Global Tuberculosis Report 2024]. Jugan Geongang Gwa Jilbyeong. 2025;18 11 Suppl:55–69. 10.56786/PHWR.2025.18.11suppl.5.

44. Chidambaram V, Tun NL, Majella MG, Ruelas Castillo J, Ayeh SK, Kumar A, et al. Male Sex Is Associated With Worse Microbiological and Clinical Outcomes Following Tuberculosis Treatment: A Retrospective Cohort Study, a Systematic Review of the Literature, and Meta-analysis. Clin Infect Dis. 2021;73:1580–8. 10.1093/cid/ciab527.

45. WHO. Global Tuberculosis Report 2024. WHO, Geneva. 2026. https://www.who.int/teams/global-programme-on-tuberculosis-and-lung-health/tb-reports/global-tuberculosis-report-2024. Accessed 7 July 2026.

