## Supplementary Material for "Treatment outcomes of drug-resistant *Mycobacterium tuberculosis* infection in Cameroon: a systematic review and meta-analysis"

**Tables S1** Search strategy by databases

| **Database** | **Search string** | **Number of entries** |
| --- | --- | --- |
| **PubMed** | ("MDR-TB"[tiab] OR "multidrug-resistant tuberculosis"[tiab] OR "multi-drug resistant tuberculosis"[tiab] OR "drug-resistant tuberculosis"[tiab]) AND ("Treatment Outcome"[Mesh] OR "treatment outcome"[tiab] OR "treatment success"[tiab] OR "cure rate"[tiab] OR "treatment completion"[tiab] OR "treatment failure"[tiab] OR "lost to follow-up"[tiab] OR "mortality"[tiab] OR "death"[tiab]) AND ("Cameroon"[Mesh] OR "Cameroon"[tiab] OR "Cameroun"[tiab]) | 13 |
| **Scopus** | TITLE-ABS-KEY("MDR-TB" OR "multidrug-resistant tuberculosis" OR "multi-drug resistant tuberculosis" OR "drug-resistant tuberculosis") AND TITLE-ABS-KEY("treatment outcome" OR "treatment success" OR "cure rate" OR "treatment completion" OR "treatment failure" OR "lost to follow-up" OR "mortality") AND TITLE-ABS-KEY("Cameroon" OR "Cameroun") | 28 |
| **Web of sciences** | ((((((TS = "MDR-TB") OR (TS = "multidrug-resistant tuberculosis")) OR (TS = "multi-drug resistant tuberculosis")) OR (TS = "drug-resistant tuberculosis")) AND (((((((((TS = "Treatment Outcome") OR (TS = "treatment outcome")) OR (TS = "treatment success")) OR (TS = "cure rate")) OR (TS = "treatment completion")) OR (TS = "treatment failure")) OR (TS = "lost to follow-up")) OR (TS = "mortality")) OR (TS = "death"))) AND ((TS = "Cameroon") OR (TS = "Cameroun"))) | 107 |
| **Embase** | ('multidrug resistant tuberculosis'/exp OR'drug resistant tuberculosis'/exp OR'MDR-TB':ti,ab,kw OR  'multidrug-resistant tuberculosis':ti,ab,kw OR  'multi-drug resistant tuberculosis':ti,ab,kw OR  'drug-resistant tuberculosis':ti,ab,kw) AND  ('treatment outcome'/exp OR'treatment success'/exp OR  'treatment failure'/exp OR'cure'/exp OR'treatment completion':ti,ab,kw OR'mortality'/exp OR  'death'/exp OR'lost to follow up'/exp OR'treatment outcome*':ti,ab,kw OR'treatment success*':ti,ab,kw OR  'cure rate*':ti,ab,kw OR'cure*':ti,ab,kw OR'treatment completion':ti,ab,kw OR'treatment failure*':ti,ab,kw OR  'lost to follow-up':ti,ab,kw OR'lost to follow up':ti,ab,kw OR'default*':ti,ab,kw OR'mortality':ti,ab,kw OR'death*':ti,ab,kw  ) AND ('cameroon'/exp ORCameroon:ti,ab,kw ORCameroun:ti,ab,kw) | 34 |
| **Cochrane Library** | (MDR-TB OR multidrug-resistant tuberculosis OR drug-resistant tuberculosis) AND (treatment outcome AND (Cameroon OR Cameroun)) | 13 |
| **AJOL** | (MDR-TB OR multidrug-resistant tuberculosis OR drug-resistant tuberculosis) AND (treatment outcome AND (Cameroon OR Cameroun)) | 96 |

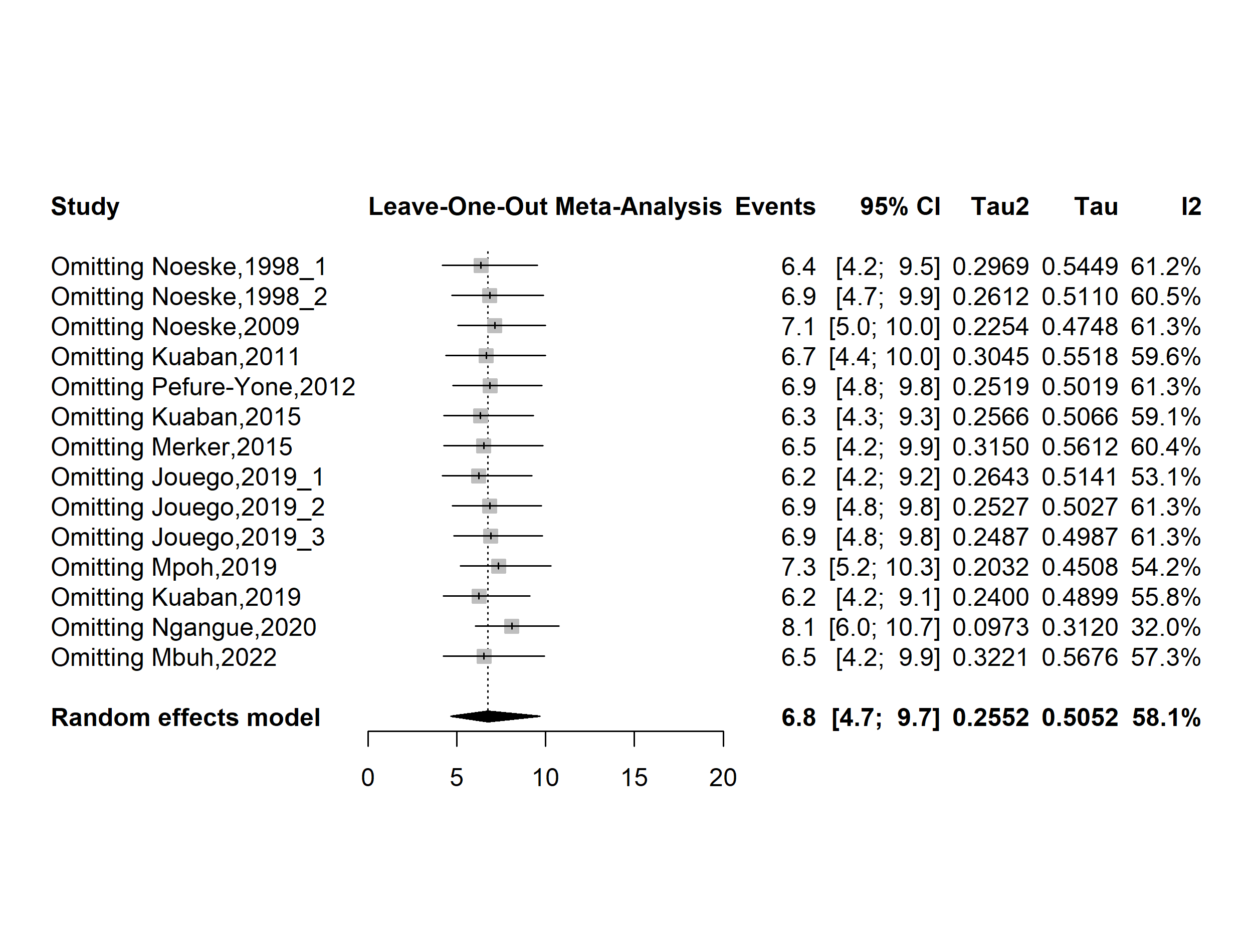

**Fig. S1** Forest plot displaying the sensitivity analysis of the pooled mortality rate among drug-resistant tuberculosis patients in Cameroon, 1998-2022

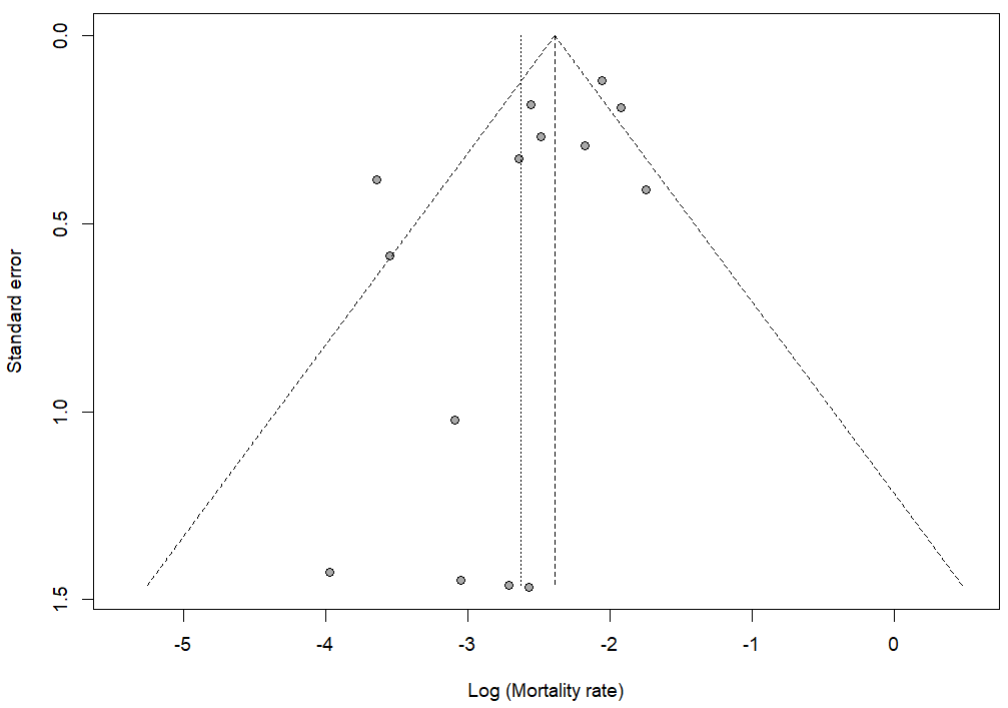

Egger’s test *p*-value = 0.062

Begg’s test *p*-value = 0.784

**Fig. S2** Funnel plot assessing publication bias among studies reporting mortality rates among drug-resistant tuberculosis patients in Cameroon, 1998–2022

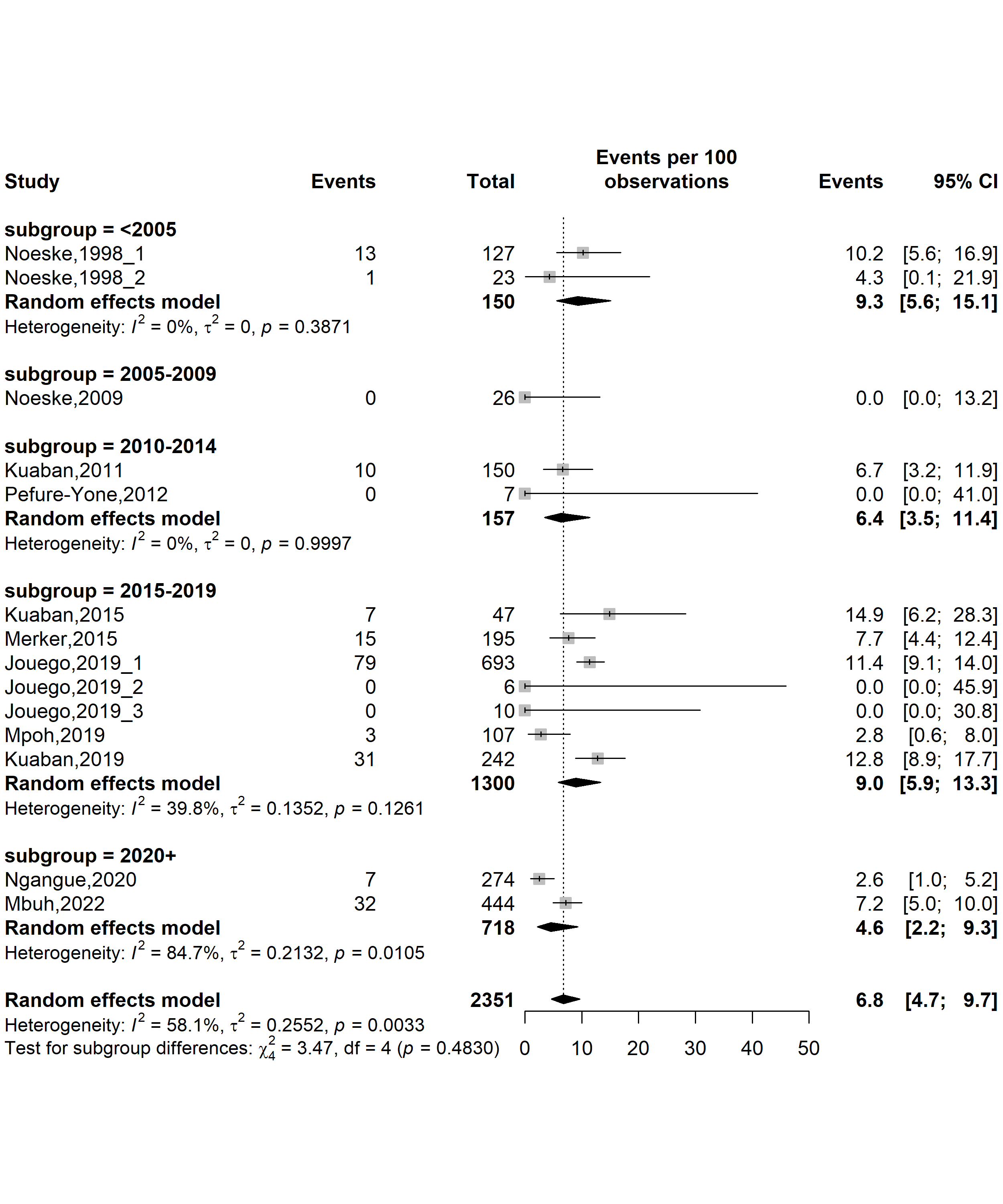

**Fig. S3** Forest plot displaying the subgroup meta-analysis of the pooled of the pooled mortality rate among drug-resistant tuberculosis patients in Cameroon by study timeframe, 1998-2022

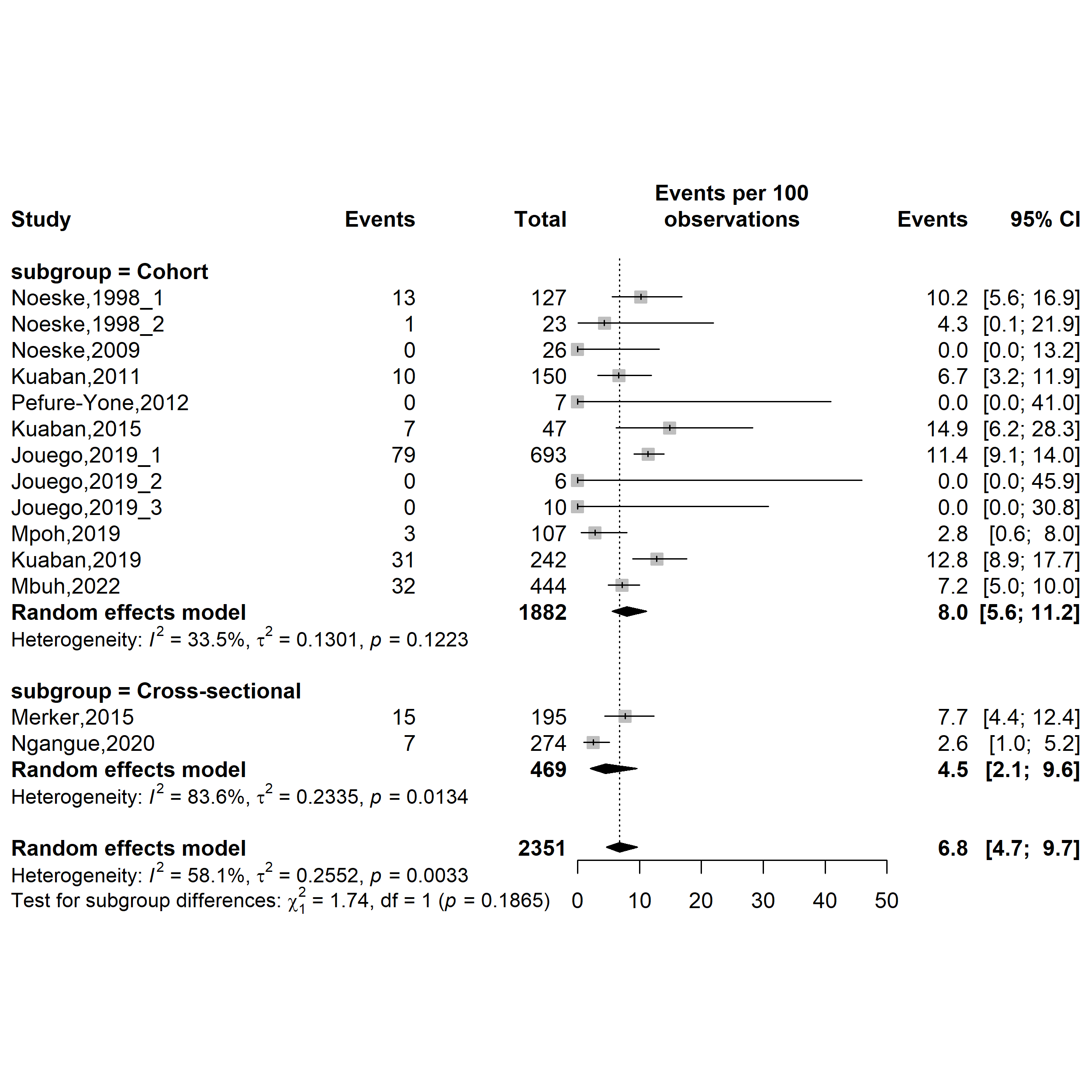

**Fig. S4** Forest plot displaying the subgroup meta-analysis of the pooled mortality rate among drug-resistant tuberculosis patients in Cameroon by study designs, 1998-2022

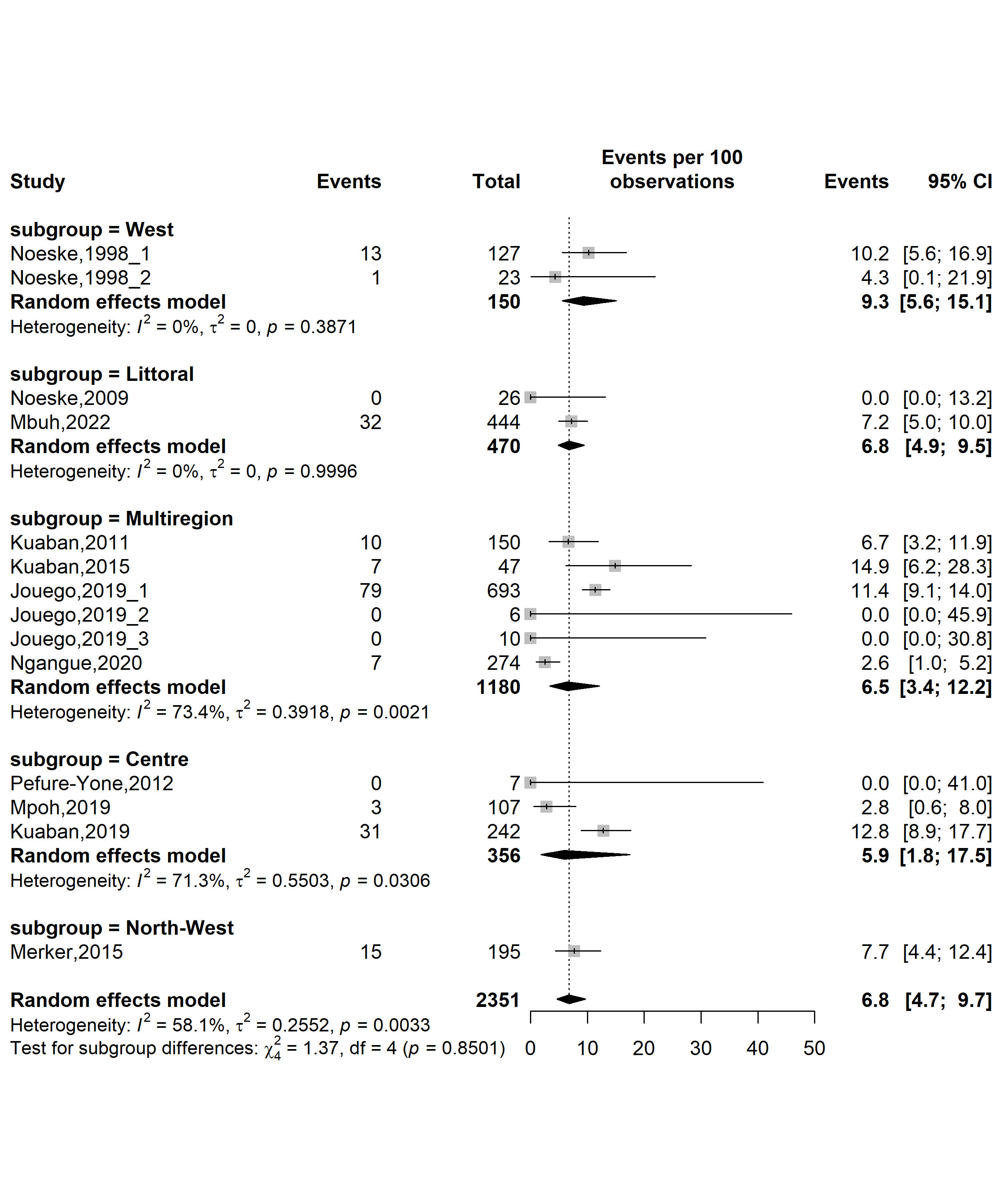

**Fig. S5** Forest plot displaying the subgroup meta-analysis of the pooled mortality rate among drug-resistant tuberculosis patients in Cameroon by study regions, 1998-2022

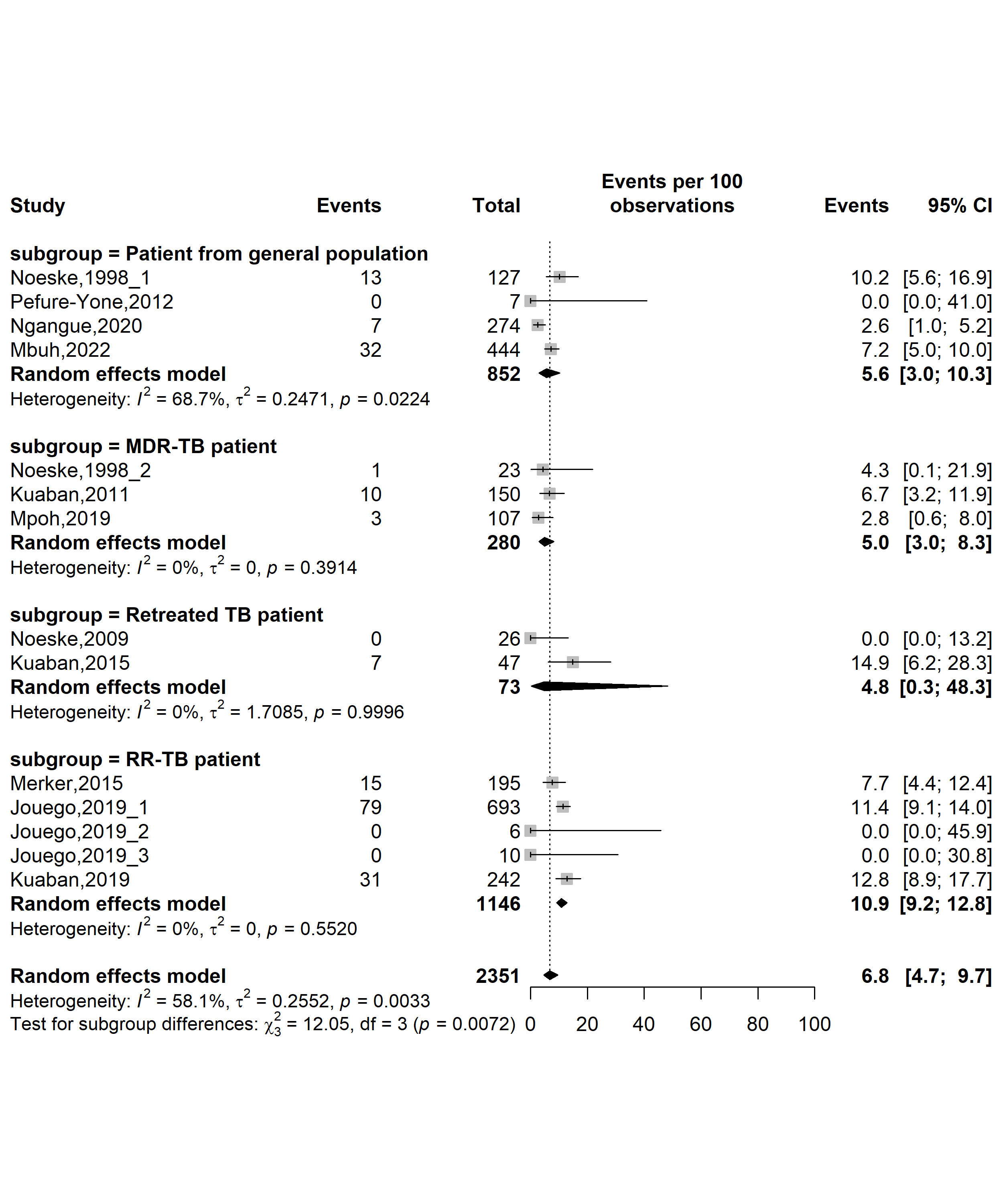

**Fig. S6** Forest plot displaying the subgroup meta-analysis of the pooled mortality rate among drug-resistant tuberculosis patients in Cameroon by types of patients, 1998-2022

*(TB: Mycobacterium tuberculosis infection; RR-TB: Rifampicin-resistant tuberculosis; MDR-TB: Multidrug-resistant tuberculosis)*

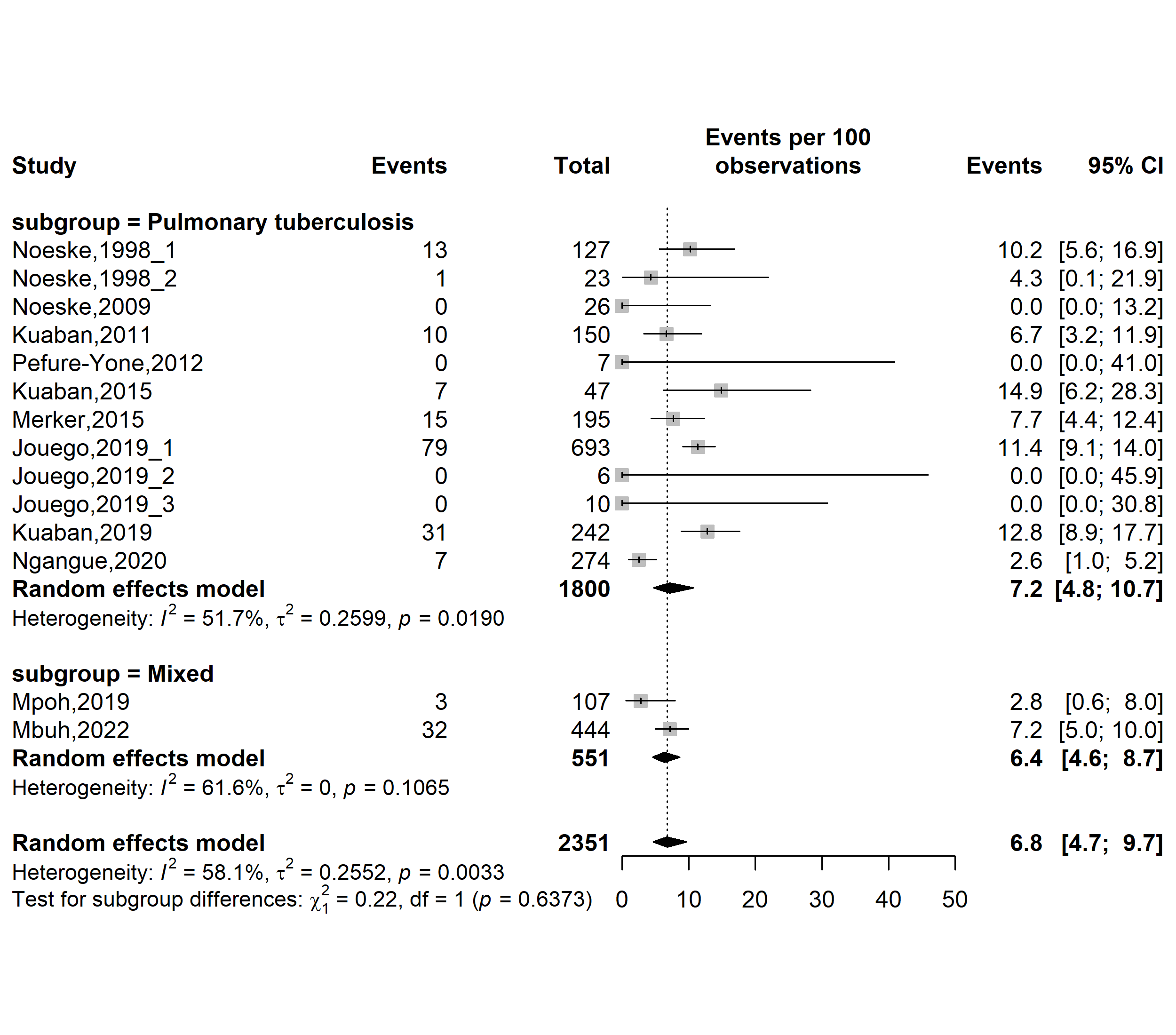

**Fig. S7** Forest plot displaying the subgroup meta-analysis of the pooled mortality rate among drug-resistant tuberculosis patients in Cameroon by tuberculosis infection localizations, 1998-2022

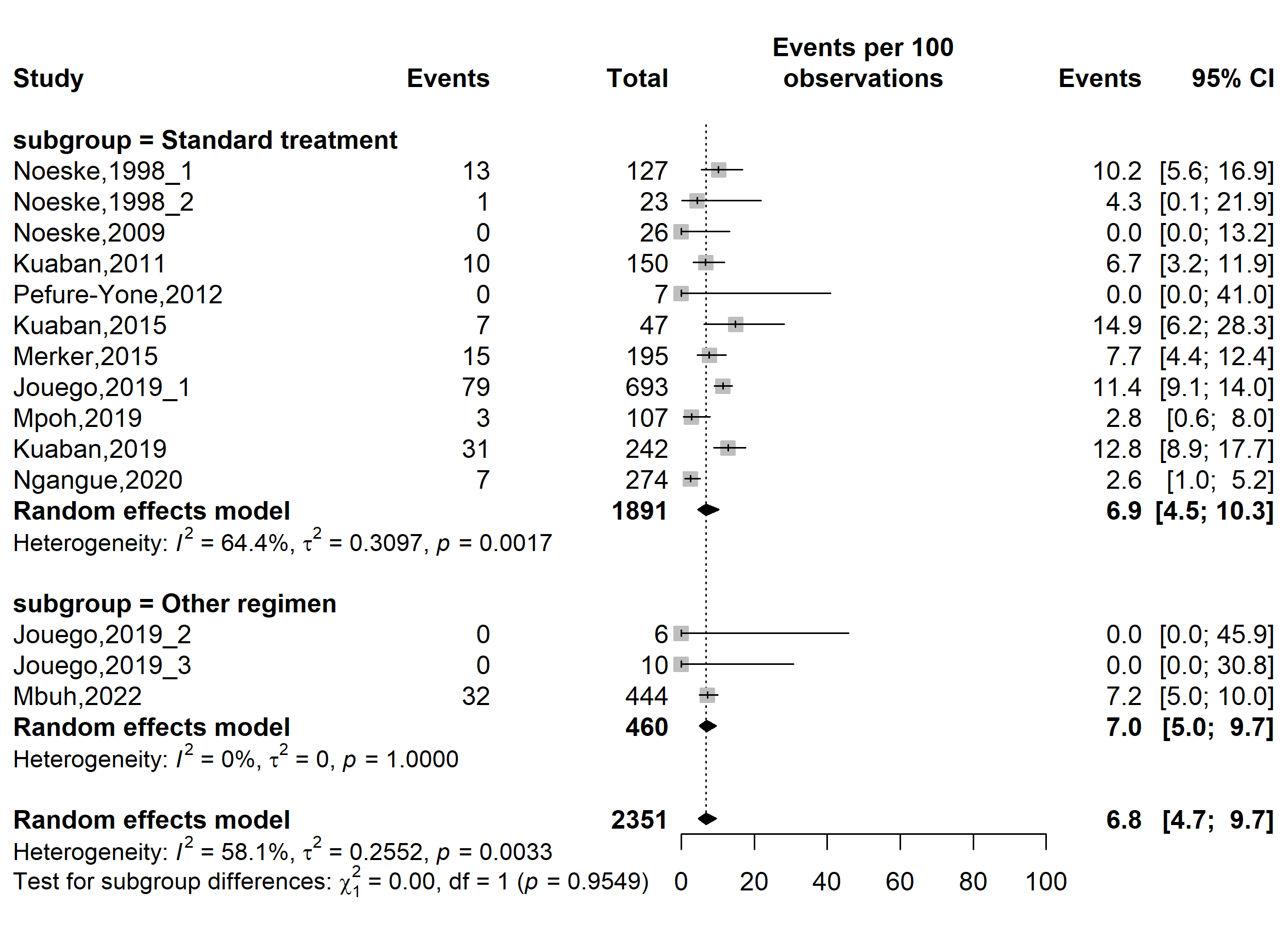

**Fig. S8** Forest plot displaying the subgroup meta-analysis of the pooled mortality rate among drug-resistant tuberculosis patients in Cameroon by types of resistant tuberculosis regimens, 1998-2022

(*Other regimen included fluroquinolone resistant modified treatment regimen [FQr-Mtr] and second-line injectable-resistant modified treatment regimen [SLIr-Mtr]*)

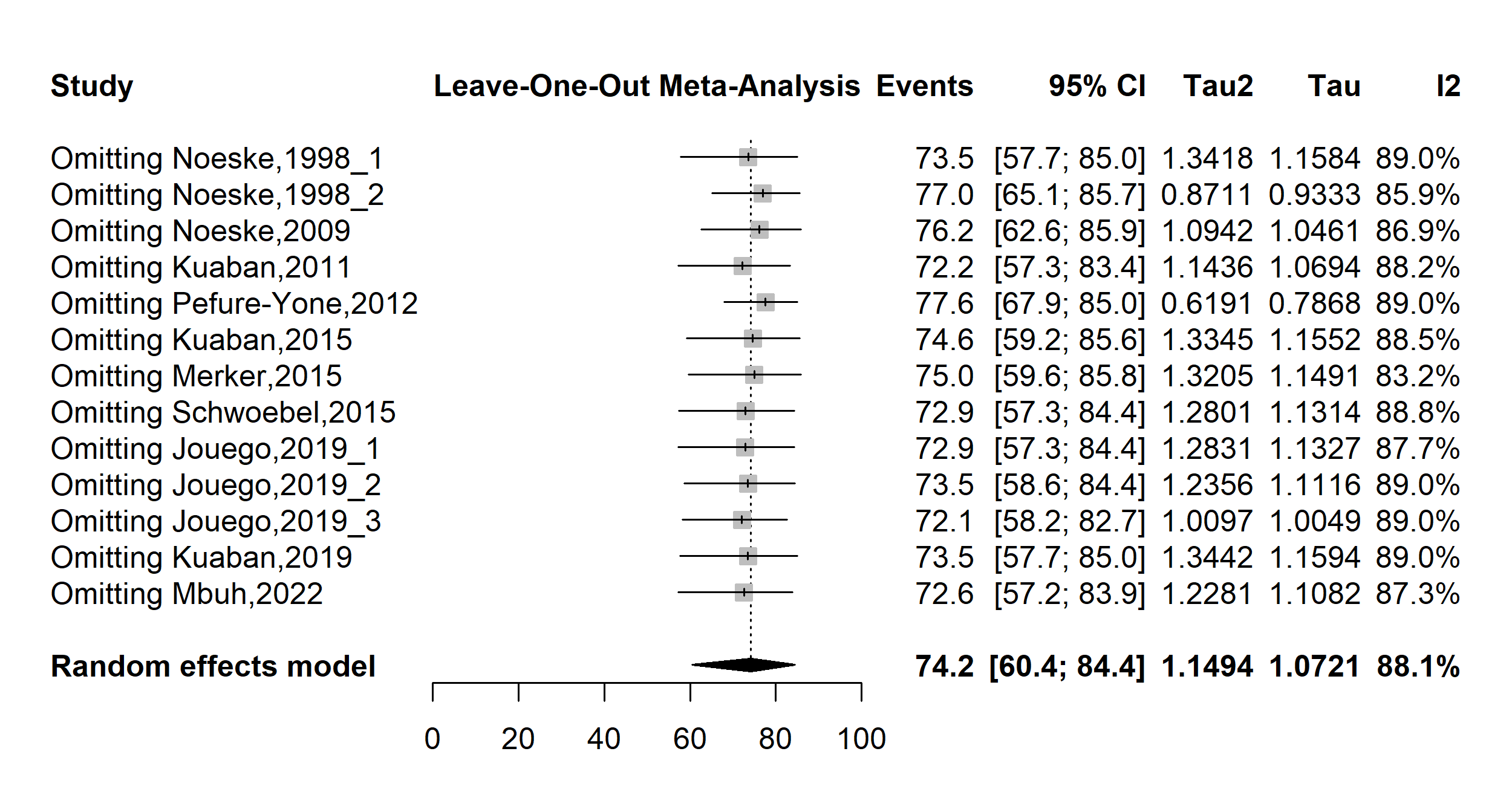

**Fig. S9** Forest plot displaying the sensitivity analysis of the pooled treatment success rate among drug-resistant tuberculosis patients in Cameroon, 1998-2022

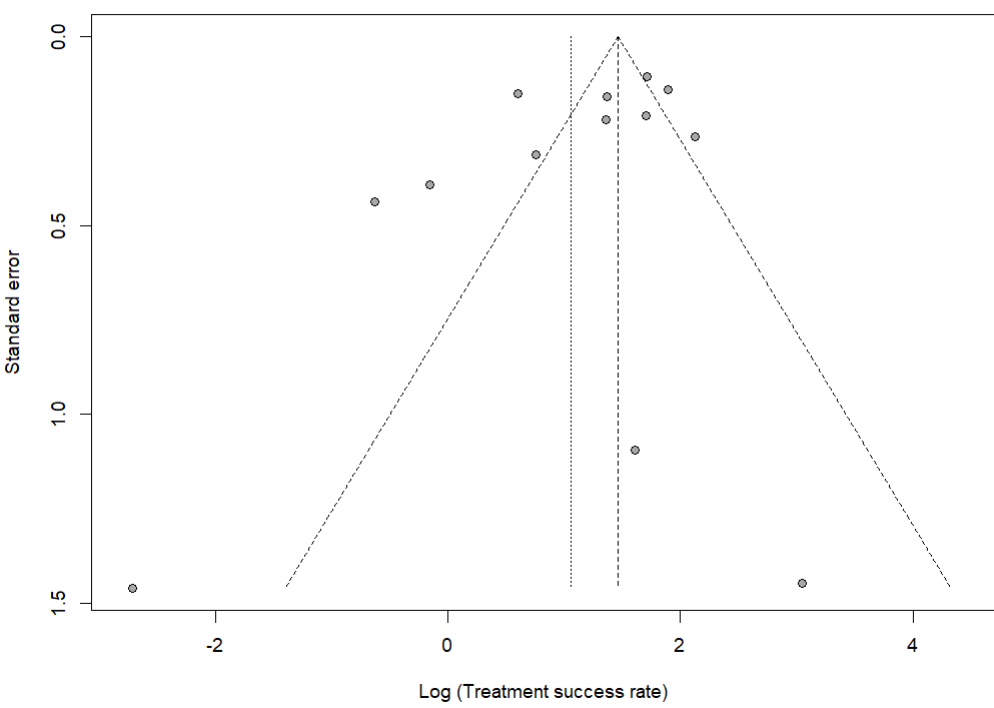

Egger’s test *p*-value = 0.191

Begg’s test *p*-value = 0.222

**Fig. S10** Funnel plot assessing publication bias among studies reporting treatment success rate among drug-resistant tuberculosis patients in Cameroon, 1998–2022

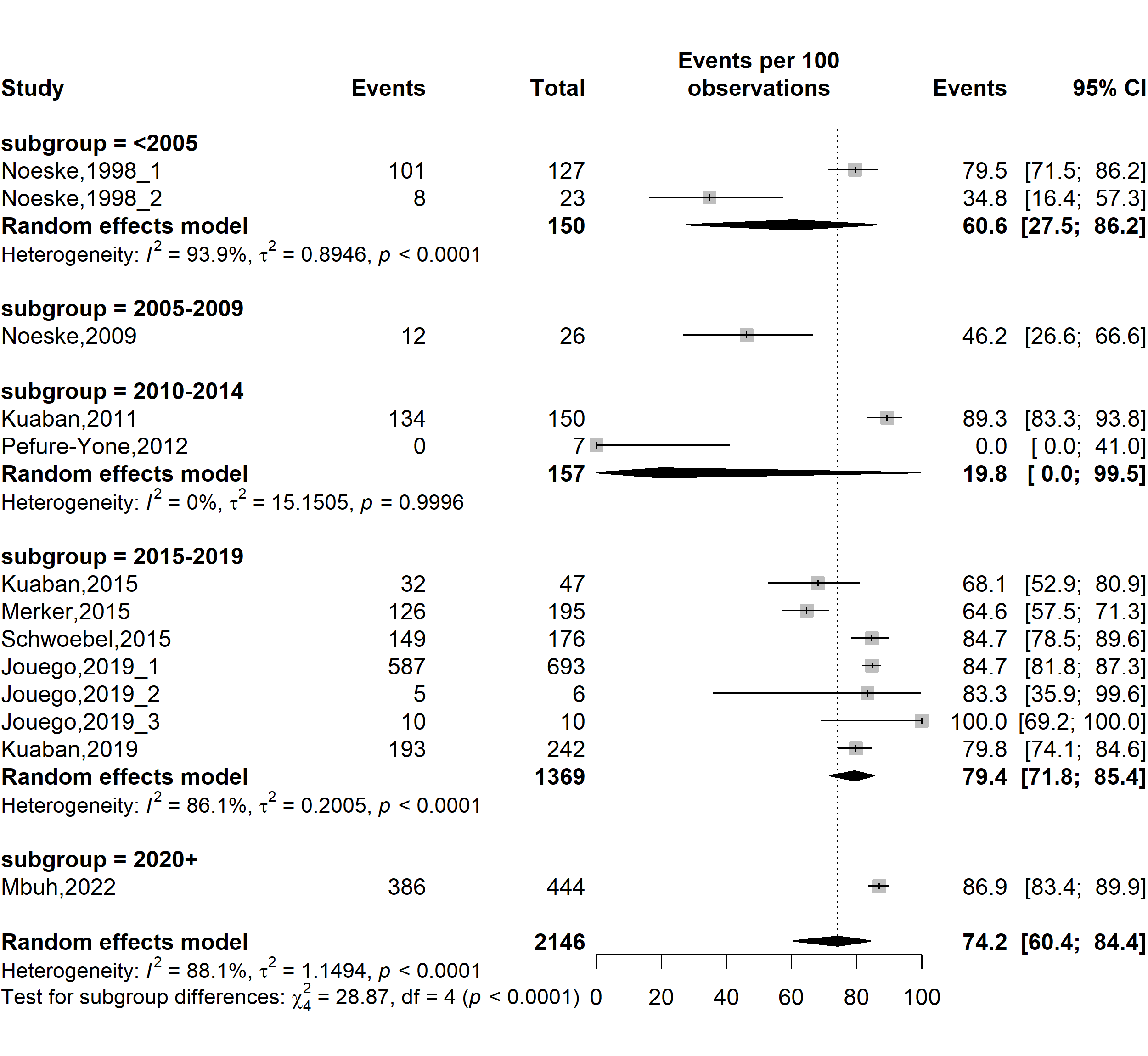

**Fig. S11** Forest plot displaying the subgroup meta-analysis of the pooled treatment success rate among drug-resistant tuberculosis patients in Cameroon by study timeframe, 1998-2022

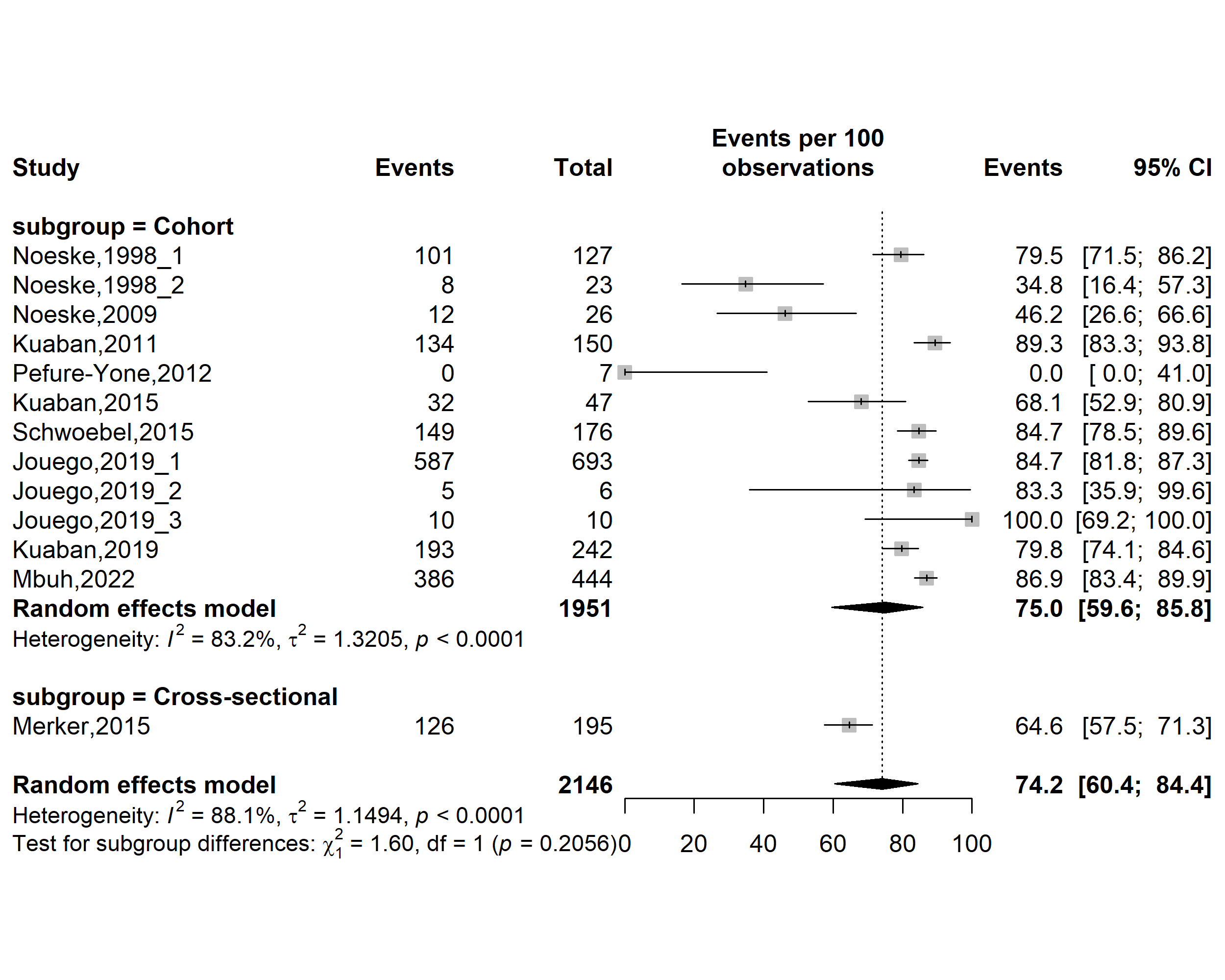

**Fig. S12** Forest plot displaying the subgroup meta-analysis of the pooled treatment success rate among drug-resistant tuberculosis patients in Cameroon by study designs, 1998-2022

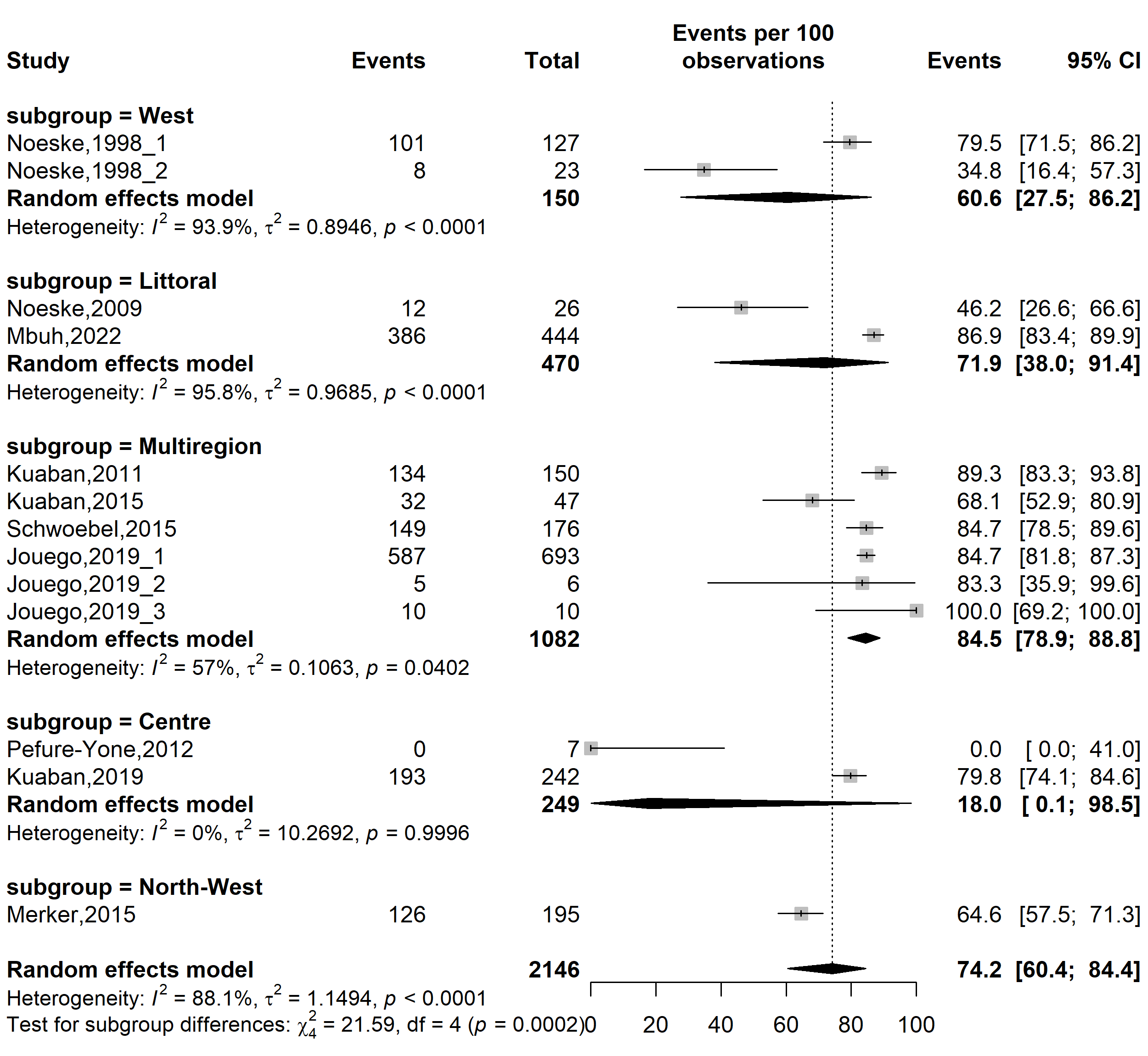

**Fig. S13** Forest plot displaying the subgroup meta-analysis of the pooled treatment success rate among drug-resistant tuberculosis patients in Cameroon by study regions, 1998-2022

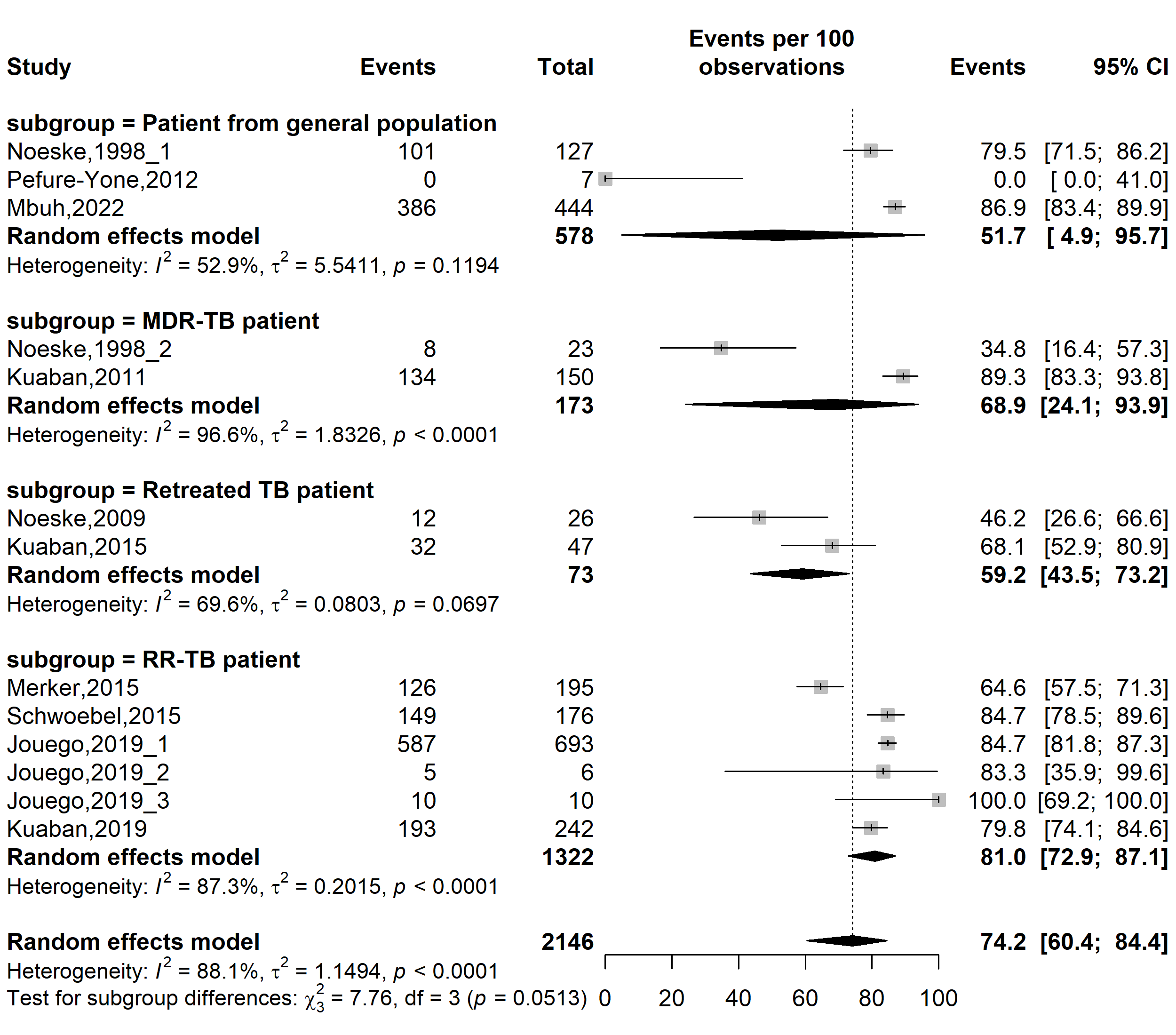

**Fig. S14** Forest plot displaying the subgroup meta-analysis of the pooled treatment success rate among drug-resistant tuberculosis patients in Cameroon by types of patients, 1998-2022

*(TB: Mycobacterium tuberculosis infection; RR-TB: Rifampicin-resistant tuberculosis; MDR-TB: Multidrug-resistant tuberculosis)*

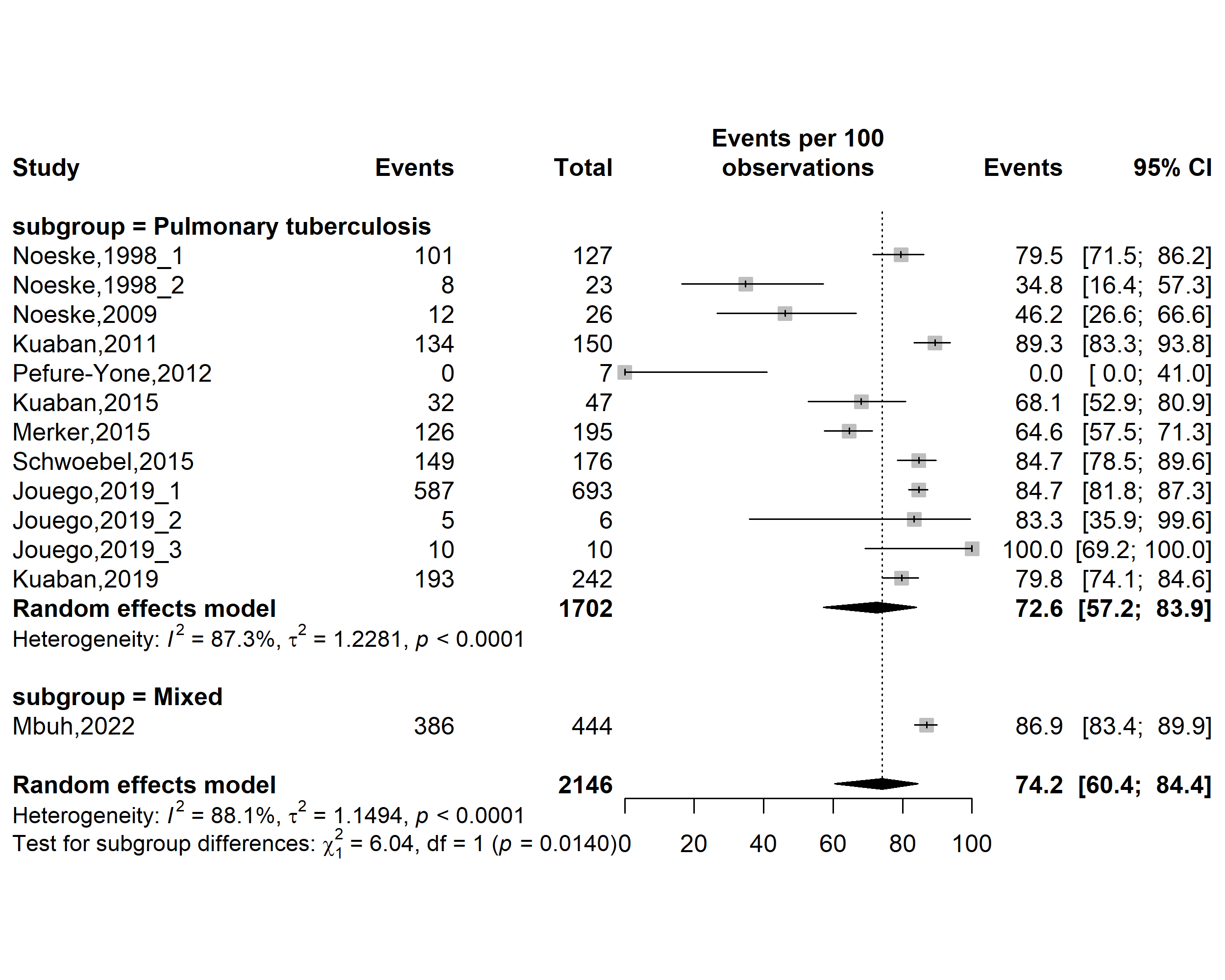

**Fig. S15** Forest plot displaying the subgroup meta-analysis of the pooled treatment success rate among drug-resistant tuberculosis patients in Cameroon by tuberculosis infection localizations, 1998-2022

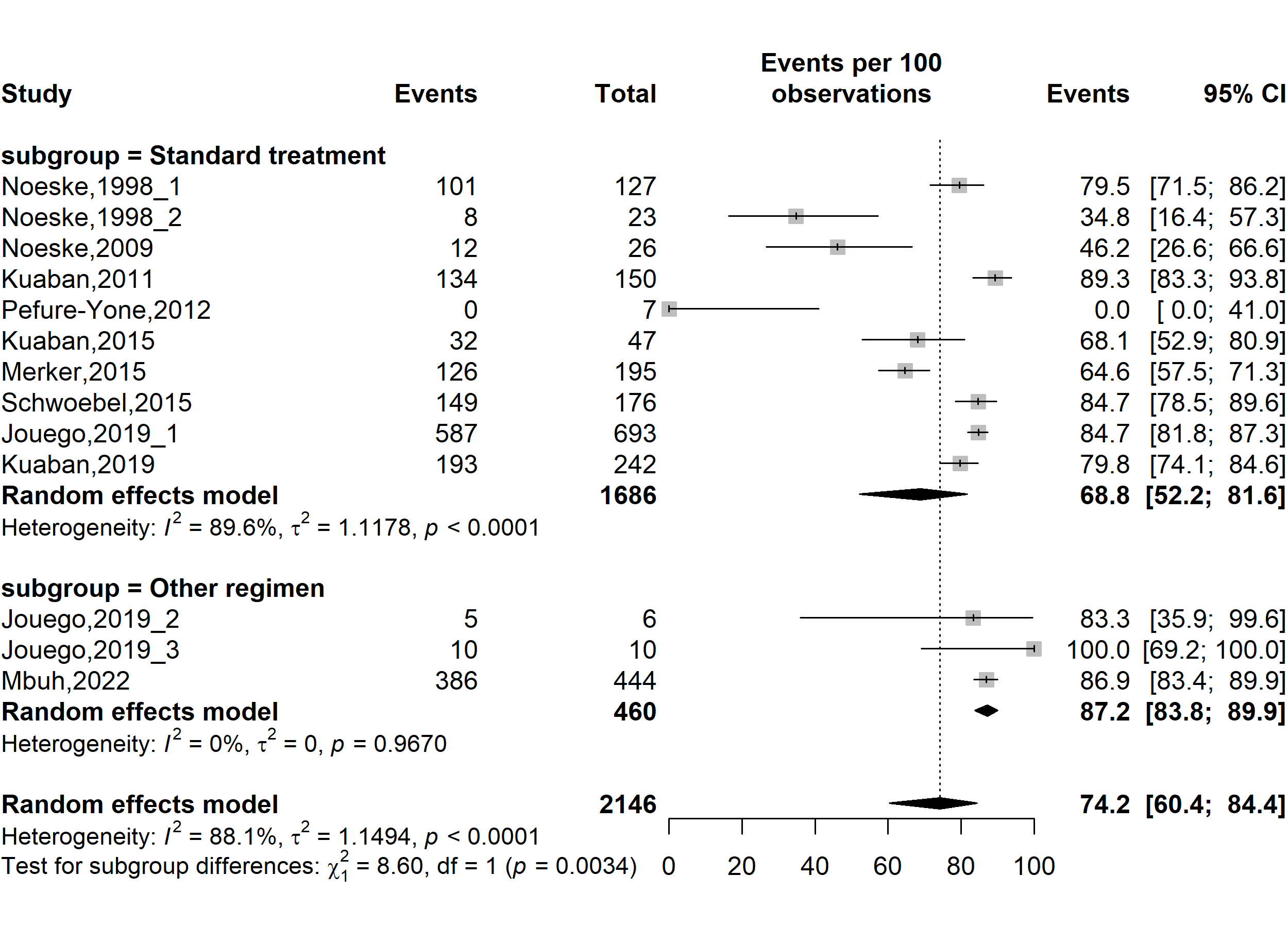

**Fig. S16** Forest plot displaying the subgroup meta-analysis of the pooled treatment success rate among drug-resistant tuberculosis patients in Cameroon by types of resistant tuberculosis regimens, 1998-2022

(*Other regimen included fluroquinolone resistant modified treatment regimen [FQr-Mtr] and second-line injectable-resistant modified treatment regimen [SLIr-Mtr]*)

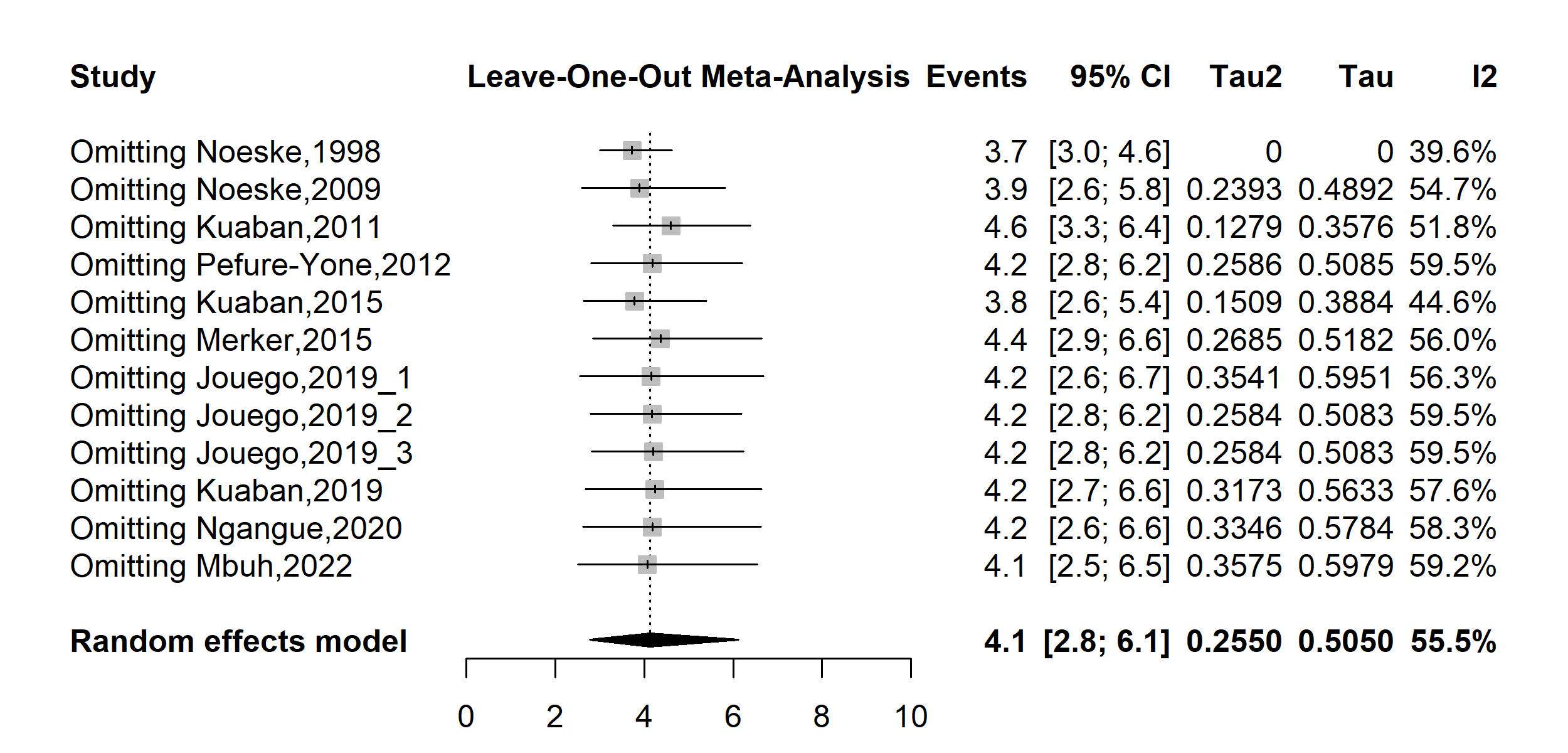

**Fig. S17** Forest plot displaying the sensitivity analysis of the pooled loss to follow-up rate among drug-resistant tuberculosis patients in Cameroon, 1998-2022

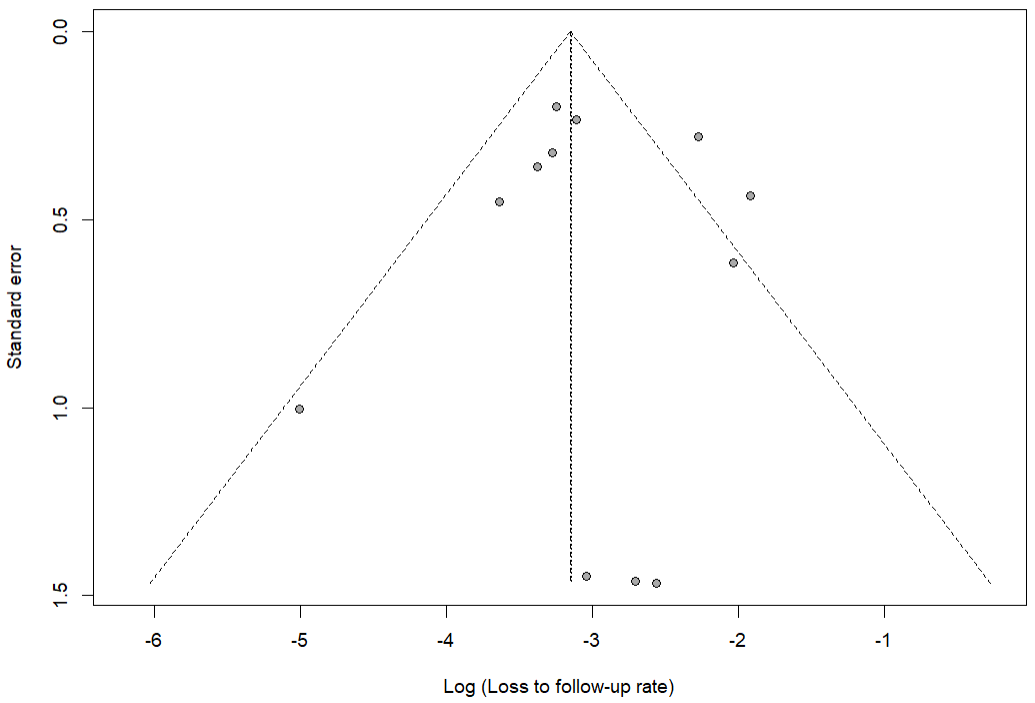

Egger’s test *p*-value = 0.885

Begg’s test *p*-value = 0.784

**Fig. S18** Funnel plot assessing publication bias among studies reporting loss to follow-up rate among drug-resistant tuberculosis patients in Cameroon, 1998–2022

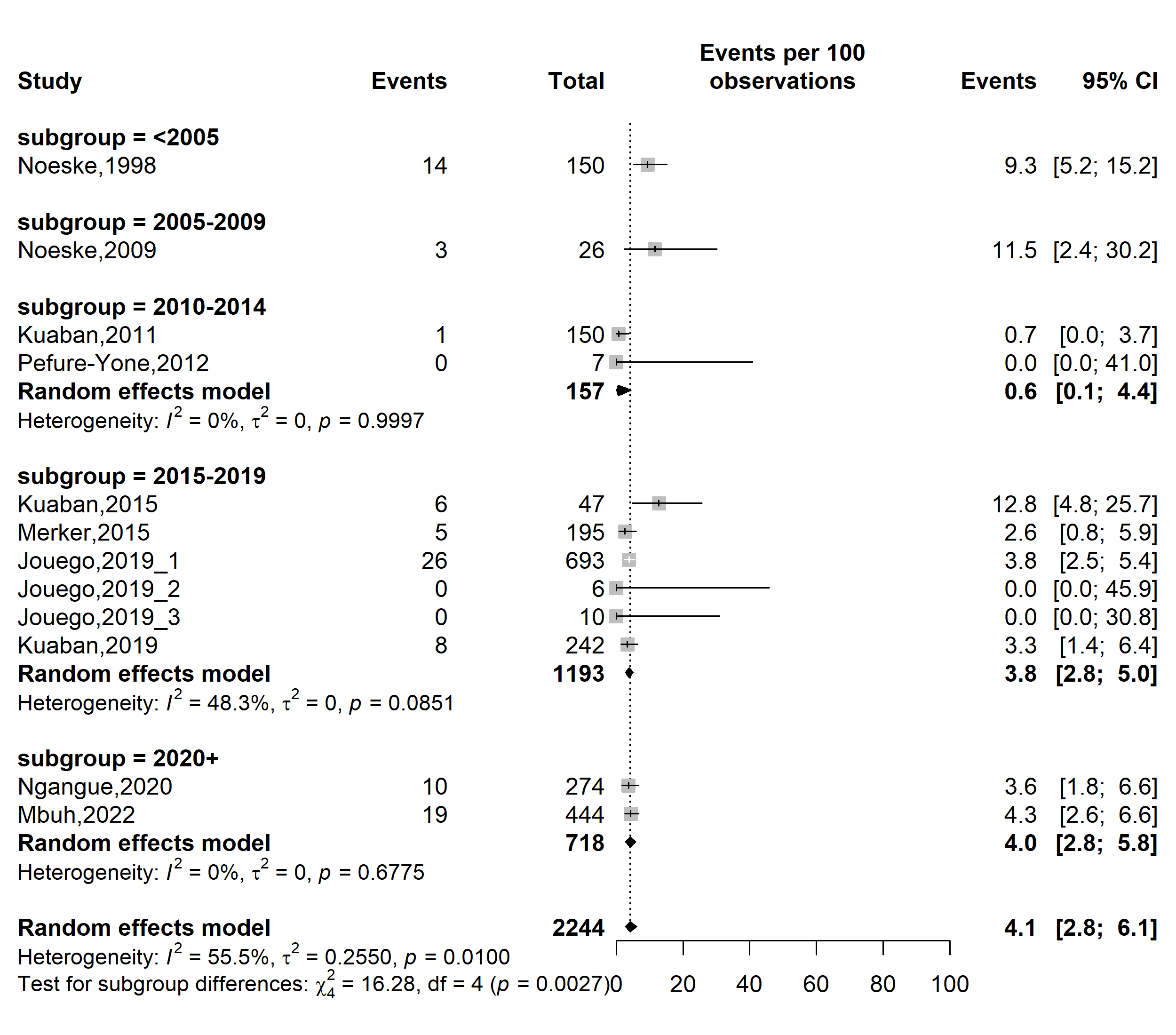

**Fig. S19** Forest plot displaying the subgroup meta-analysis of the pooled loss to follow-up rate among drug-resistant tuberculosis patients in Cameroon by study timeframe, 1998-2022

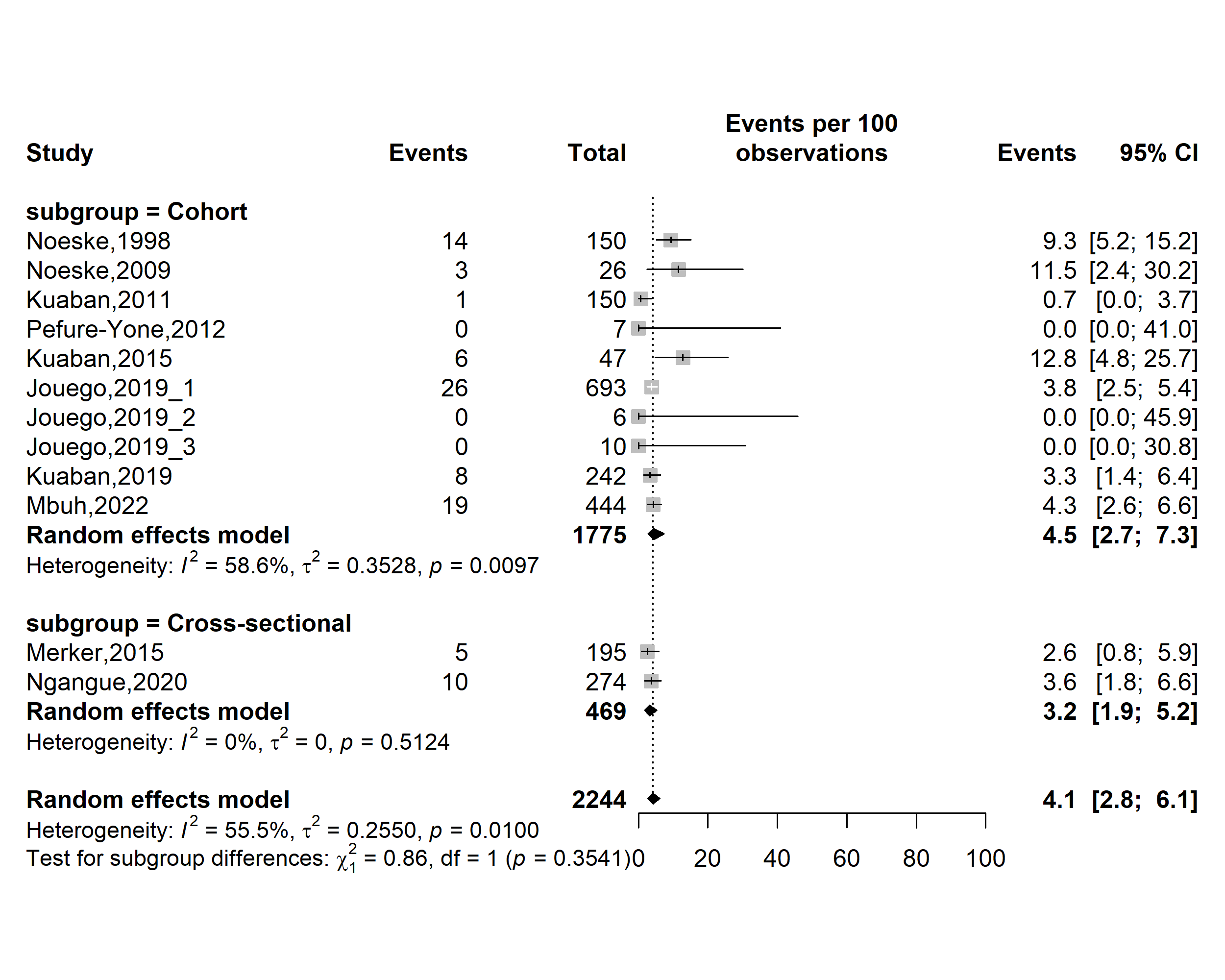

**Fig. S20** Forest plot displaying the subgroup meta-analysis of the pooled loss to follow-up rate among drug-resistant tuberculosis patients in Cameroon by study designs, 1998-2022

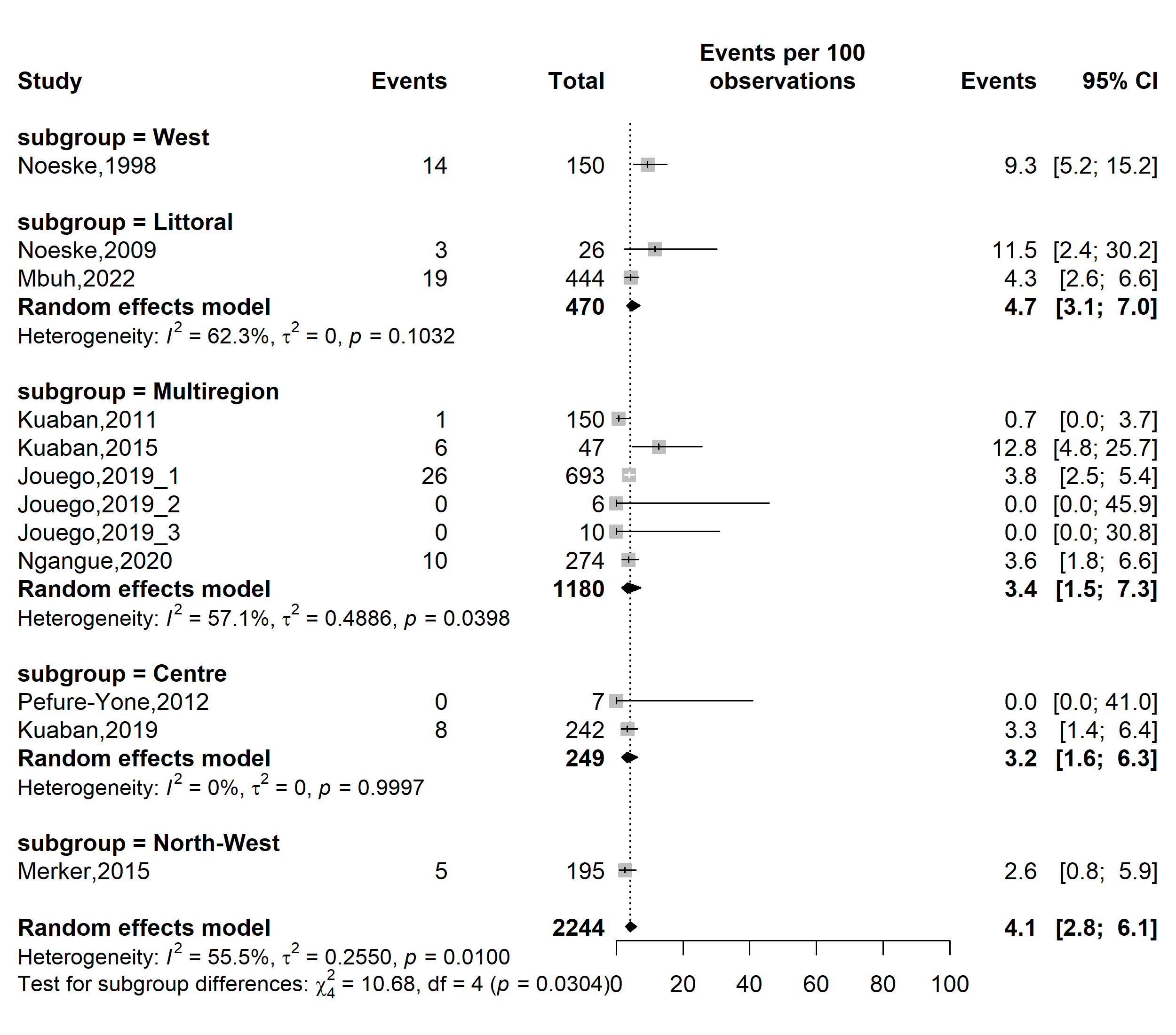

**Fig. S21** Forest plot displaying the subgroup meta-analysis of the pooled loss to follow-up rate among drug-resistant tuberculosis patients in Cameroon by study regions, 1998-2022

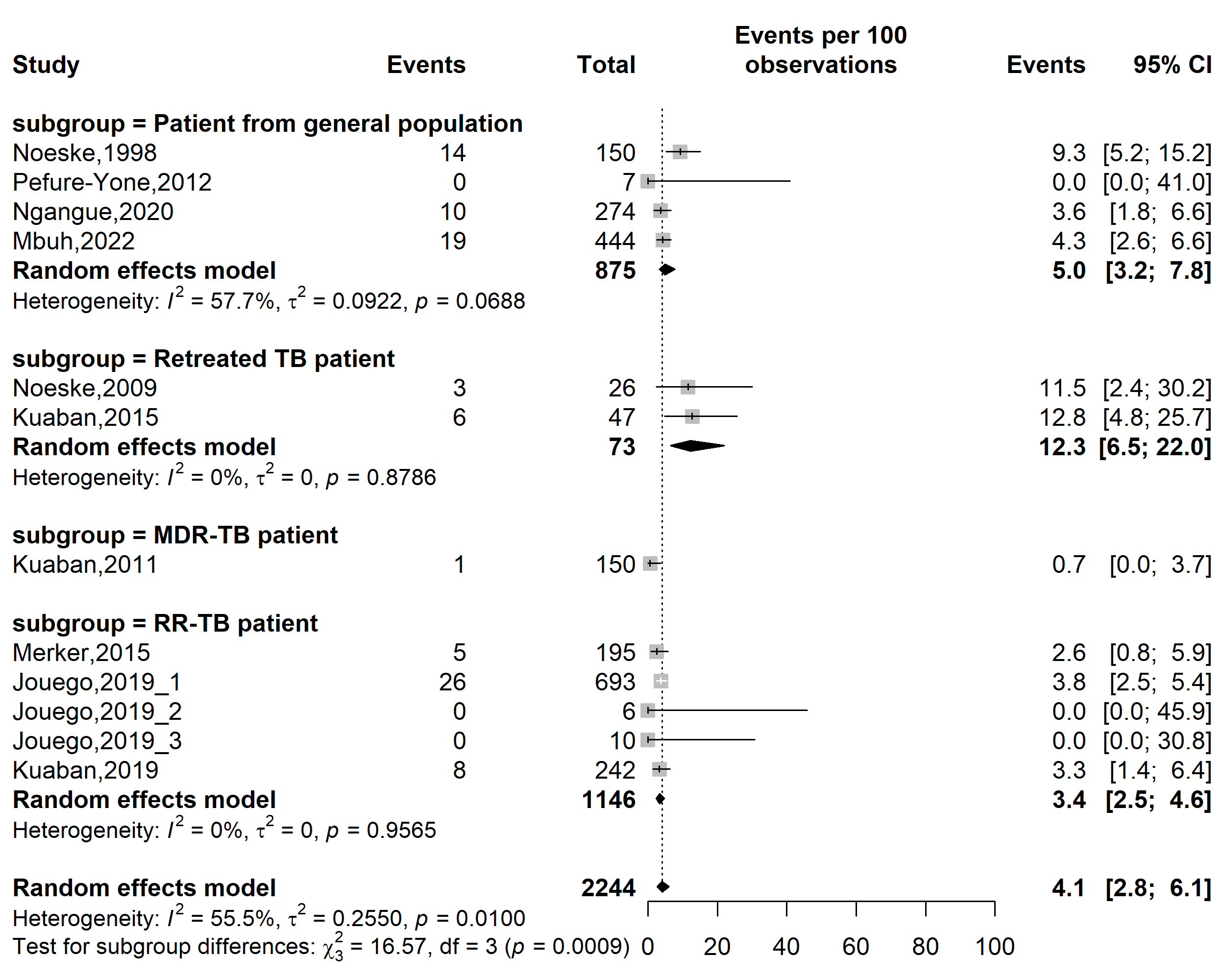

**Fig. S22** Forest plot displaying the subgroup meta-analysis of the pooled loss to follow-up rate among drug-resistant tuberculosis patients in Cameroon by types of patients, 1998-2022

*(TB: Mycobacterium tuberculosis infection; RR-TB: Rifampicin-resistant tuberculosis; MDR-TB: Multidrug-resistant tuberculosis)*

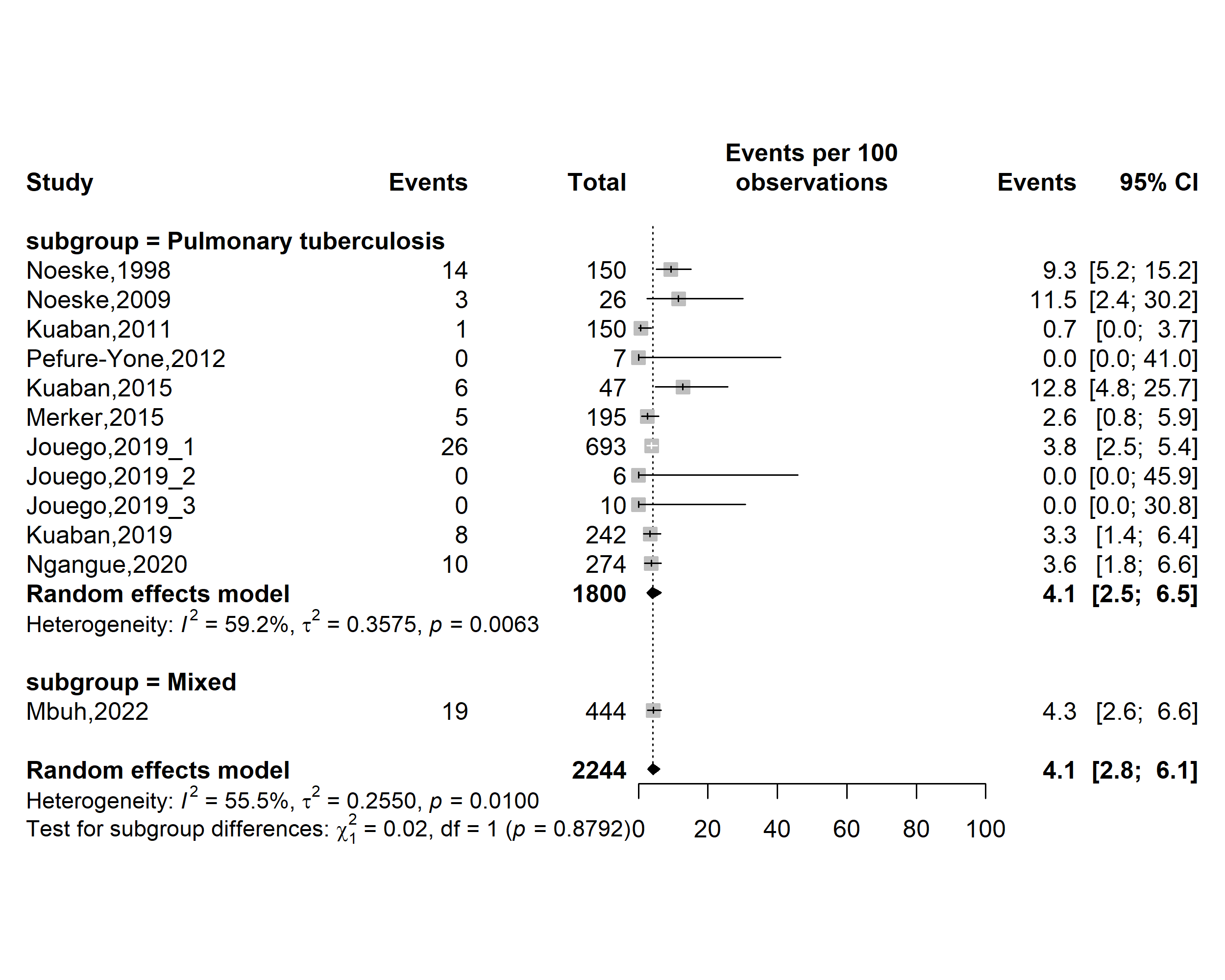

**Fig. S23** Forest plot displaying the subgroup meta-analysis of the pooled loss to follow-up rate among drug-resistant tuberculosis patients in Cameroon by tuberculosis infection localizations, 1998-2022

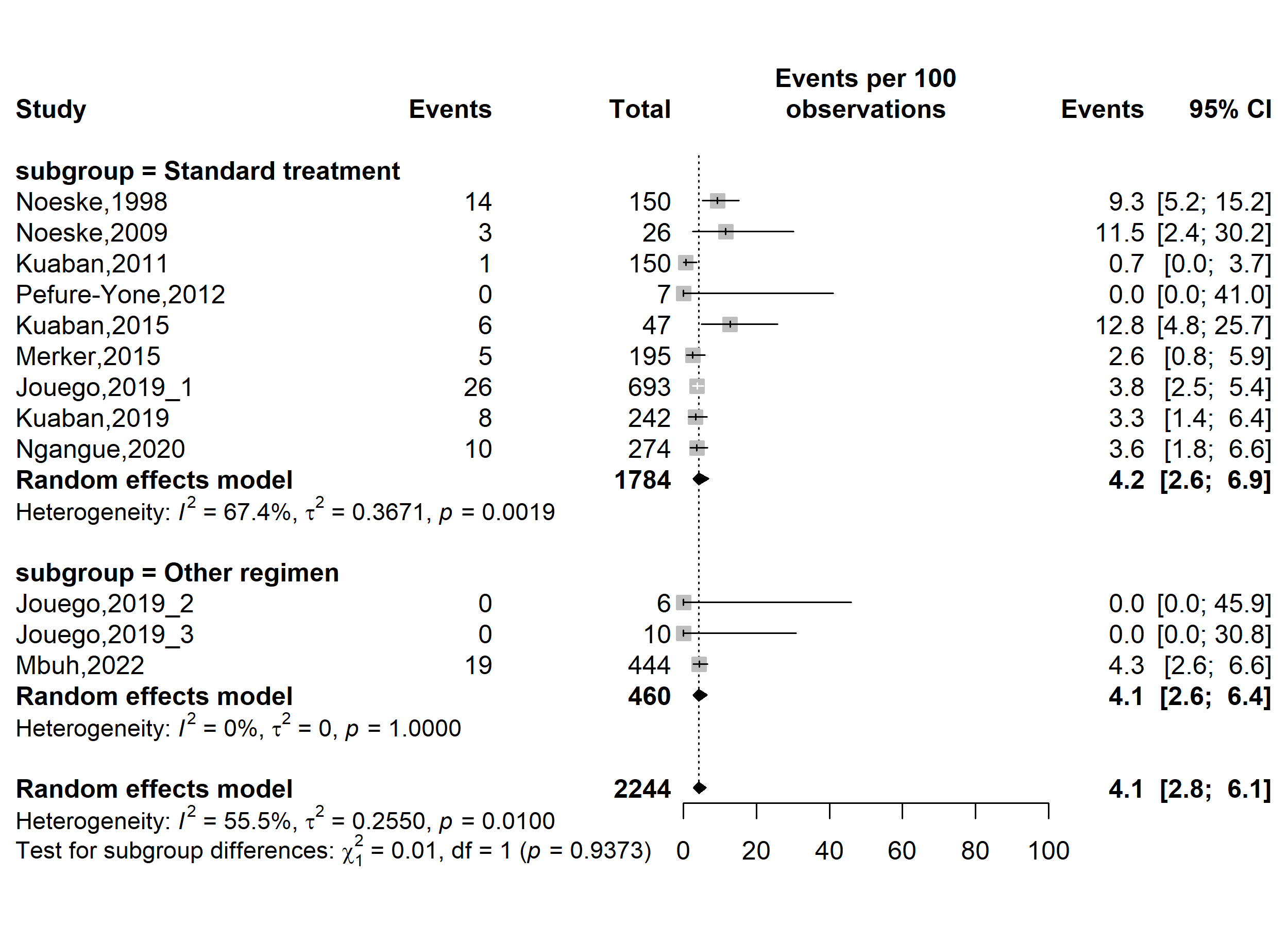

**Fig. S24** Forest plot displaying the subgroup meta-analysis of the pooled loss to follow-up rate among drug-resistant tuberculosis patients in Cameroon by types of resistant tuberculosis regimens, 1998-2022

(*Other regimen included fluroquinolone resistant modified treatment regimen [FQr-Mtr] and second-line injectable-resistant modified treatment regimen [SLIr-Mtr]*)

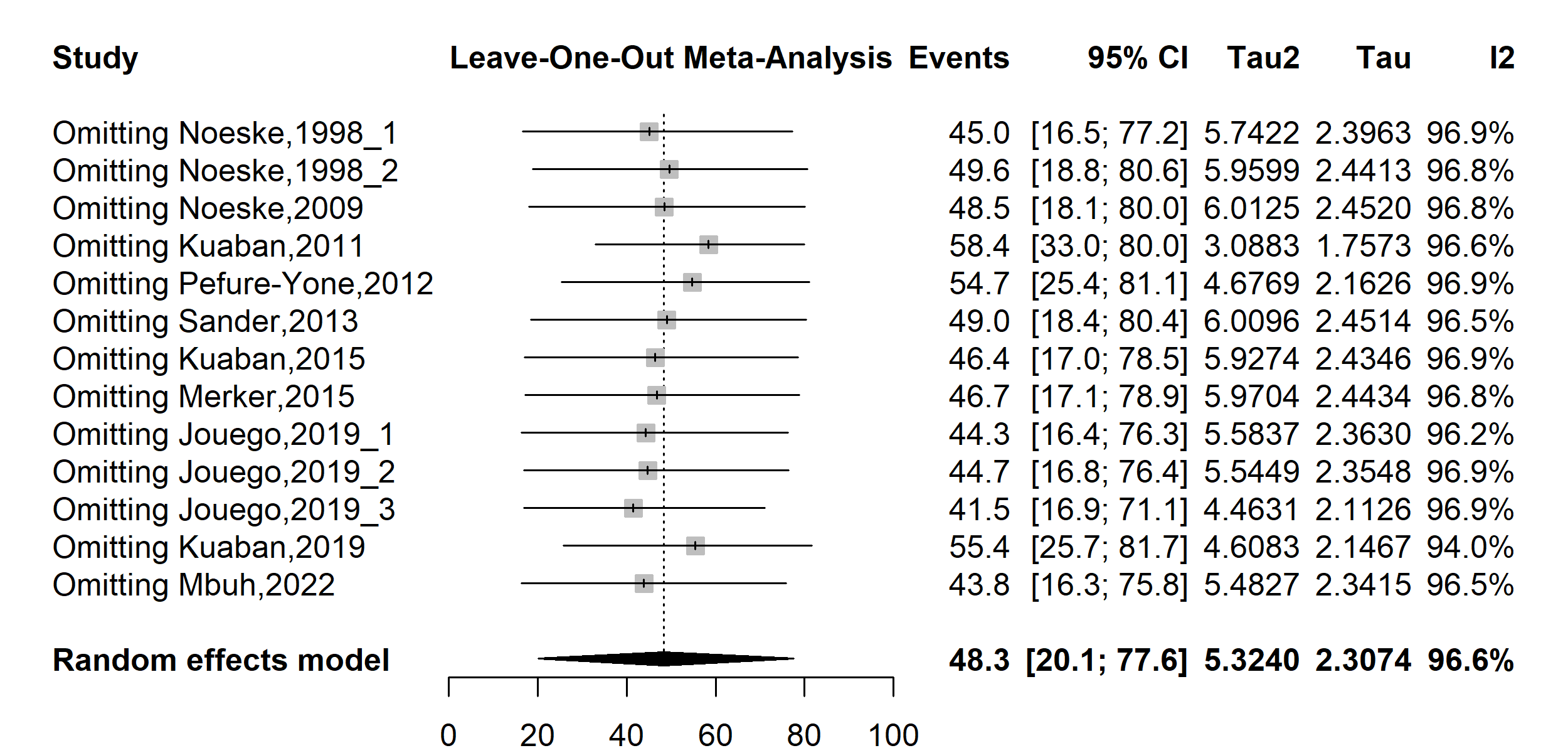

**Fig. S25** Forest plot displaying the sensitivity analysis of the pooled treatment failure rate among drug-resistant tuberculosis patients in Cameroon, 1998-2022

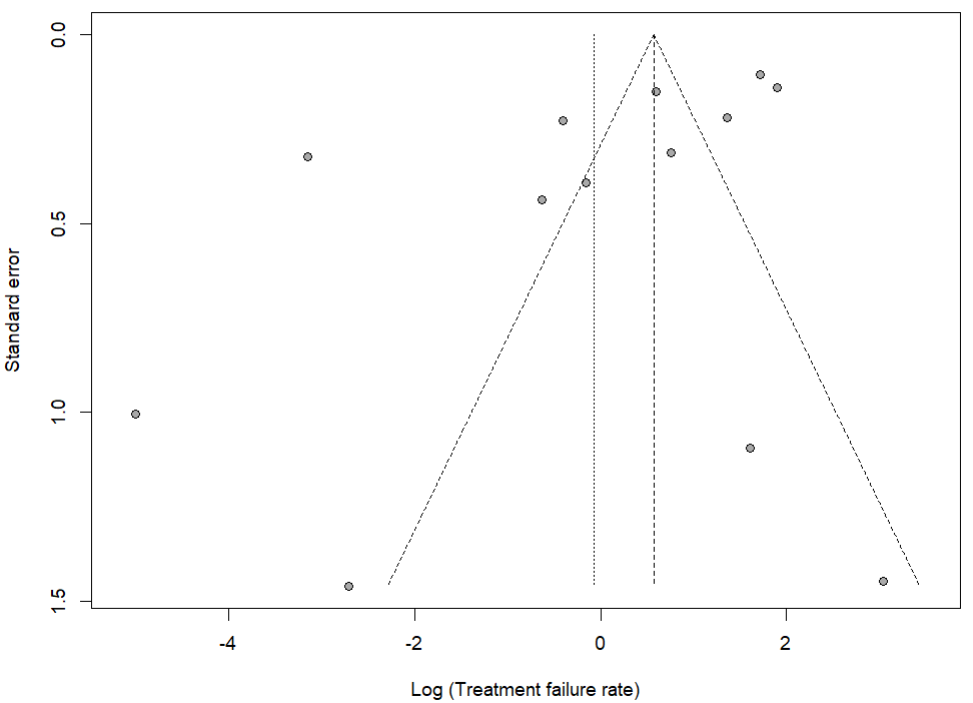

Egger’s test *p*-value = 0.056

Begg’s test *p*-value = 0.329

**Fig. S26** Funnel plot assessing publication bias among studies reporting treatment failure rate among drug-resistant tuberculosis patients in Cameroon, 1998–2022

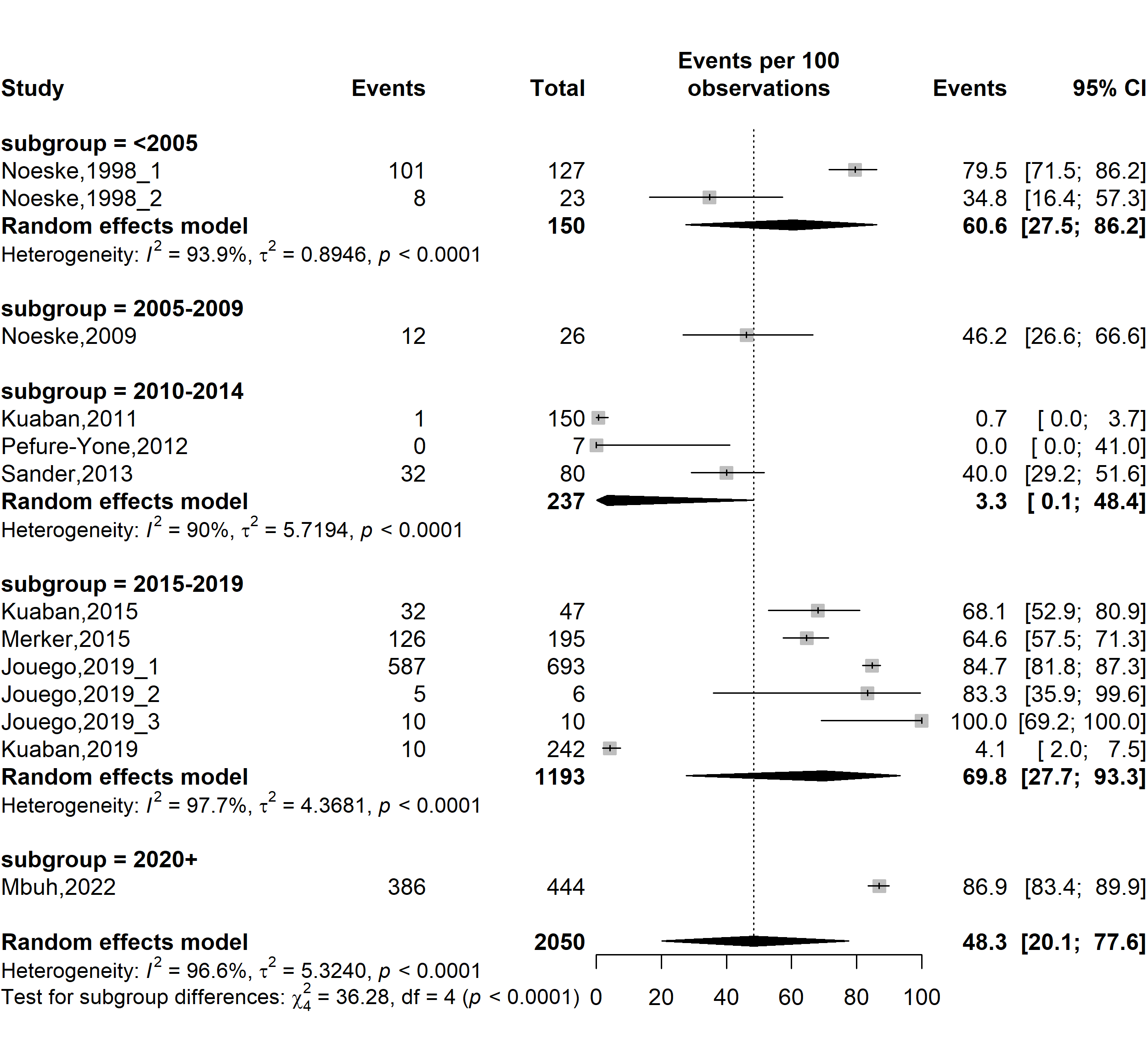

**Fig. S27** Forest plot displaying the subgroup meta-analysis of the pooled treatment failure rate among drug-resistant tuberculosis patients in Cameroon by study timeframe, 1998-2022

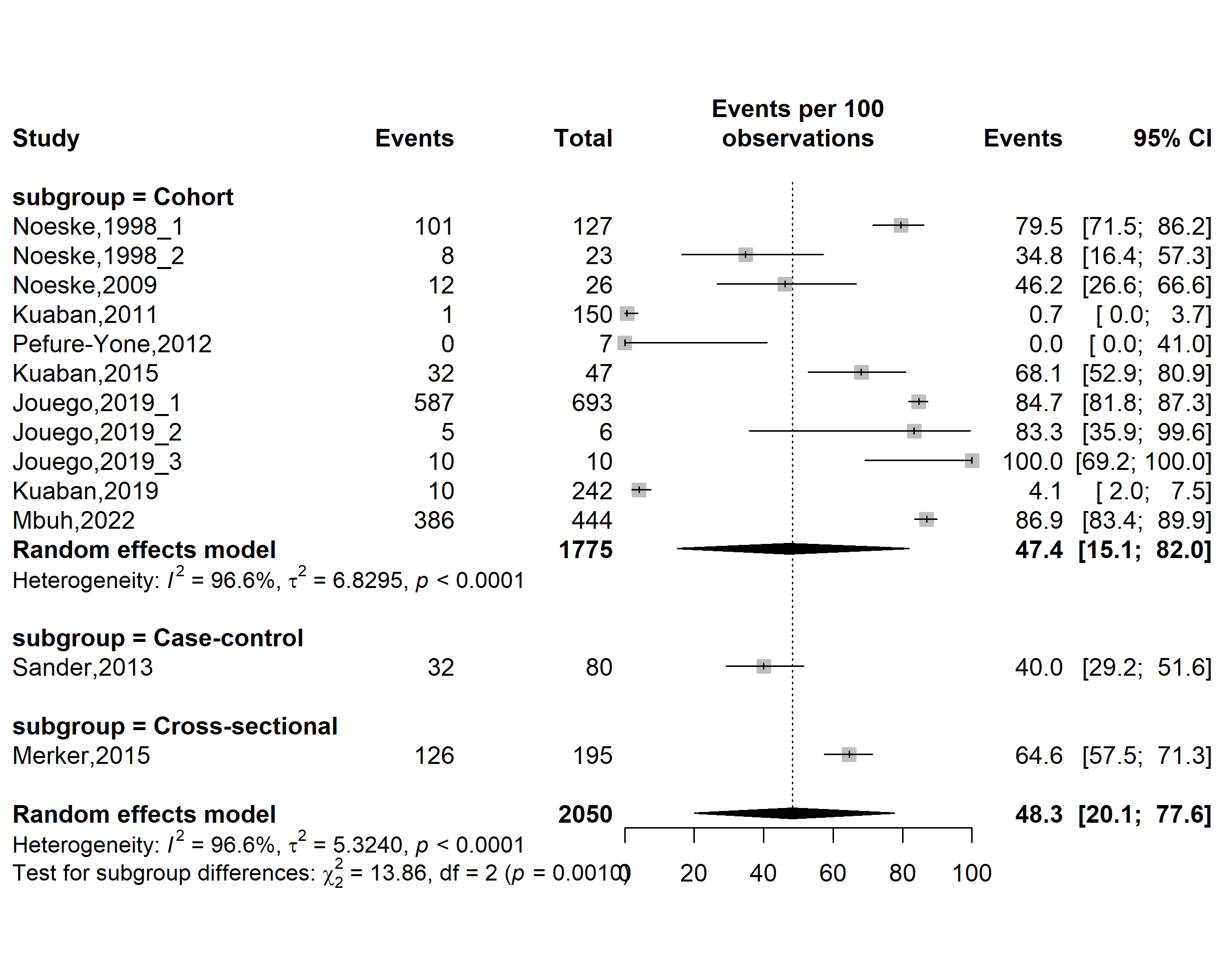

**Fig. S28** Forest plot displaying the subgroup meta-analysis of the pooled treatment failure rate among drug-resistant tuberculosis patients in Cameroon by study designs, 1998-2022

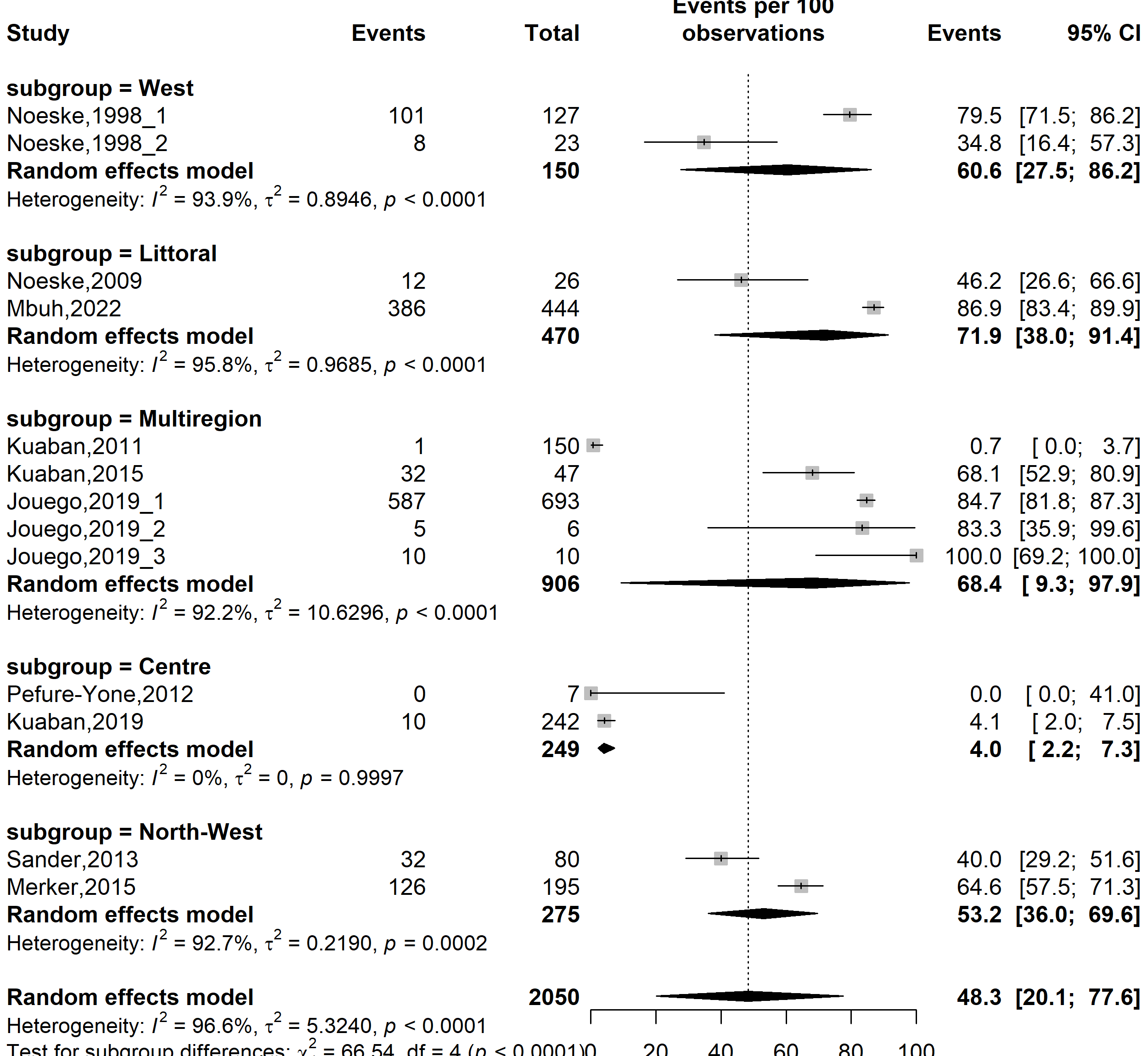

**Fig. S29** Forest plot displaying the subgroup meta-analysis of the pooled treatment failure rate among drug-resistant tuberculosis patients in Cameroon by study regions, 1998-2022

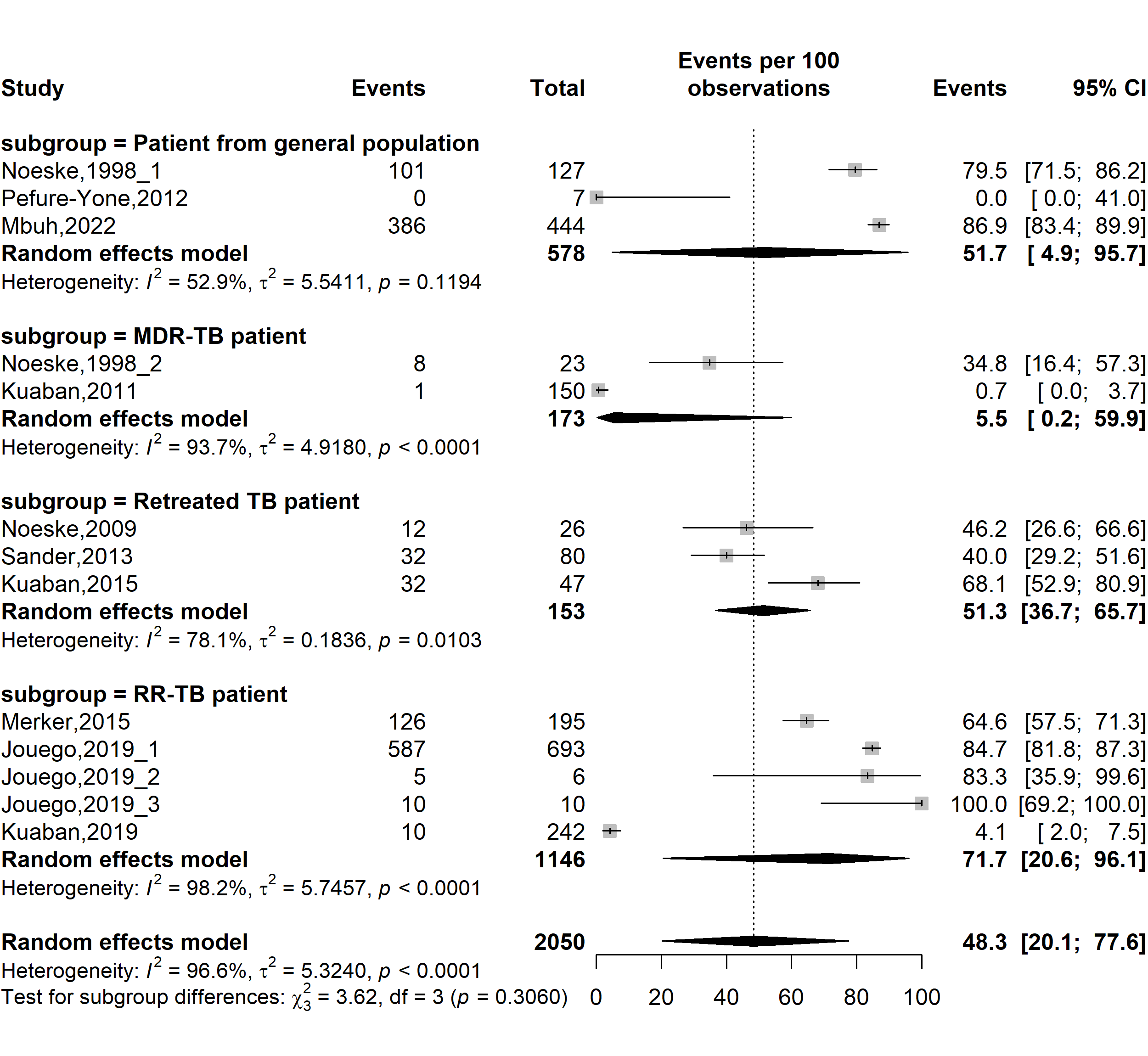

**Fig. S30** Forest plot displaying the subgroup meta-analysis of the pooled treatment failure rate among drug-resistant tuberculosis patients in Cameroon by types of patients, 1998-2022

*(TB: Mycobacterium tuberculosis infection; RR-TB: Rifampicin-resistant tuberculosis; MDR-TB: Multidrug-resistant tuberculosis)*

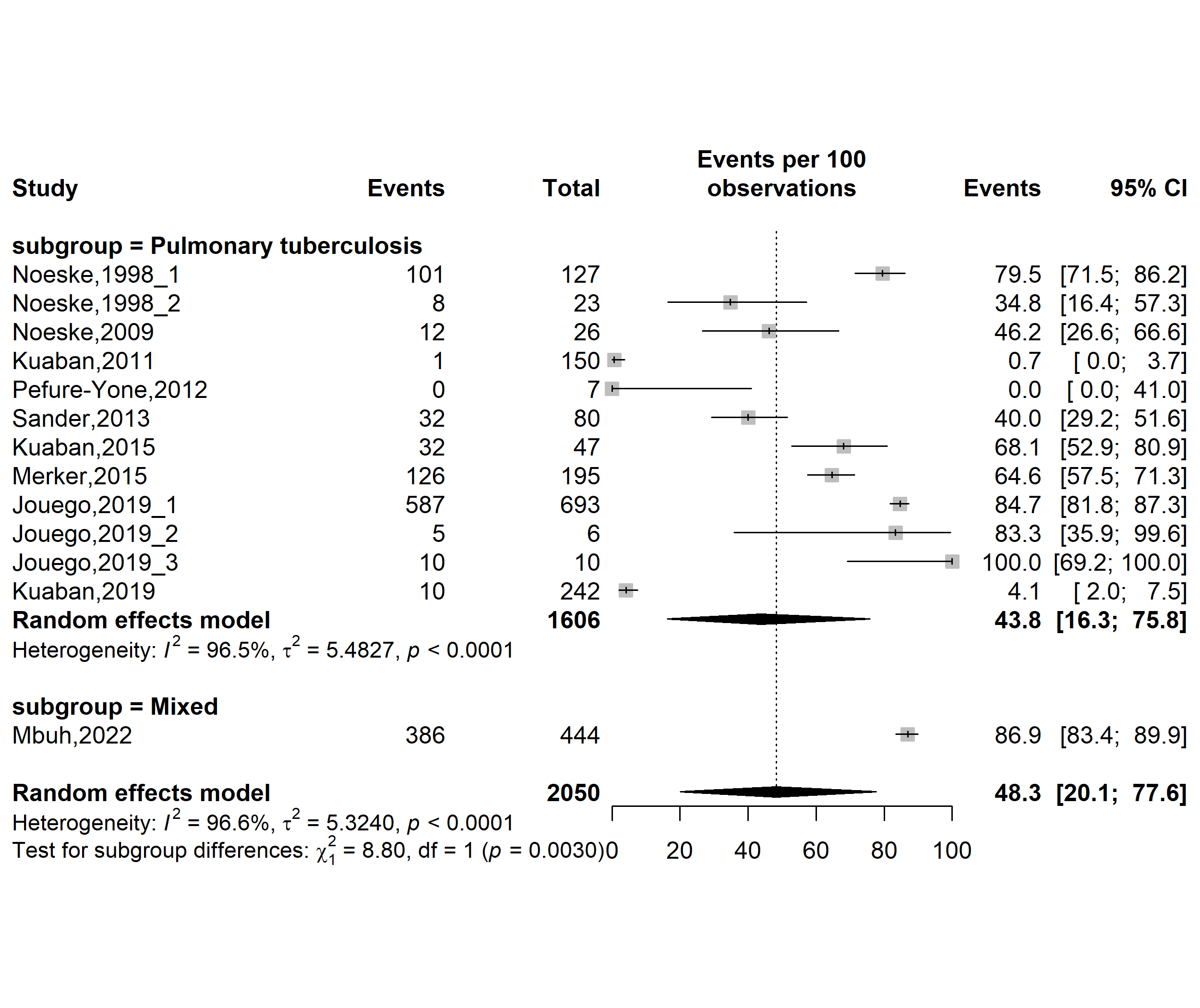

**Fig. S31** Forest plot displaying the subgroup meta-analysis of the pooled treatment failure rate among drug-resistant tuberculosis patients in Cameroon by tuberculosis infection localizations, 1998-2022

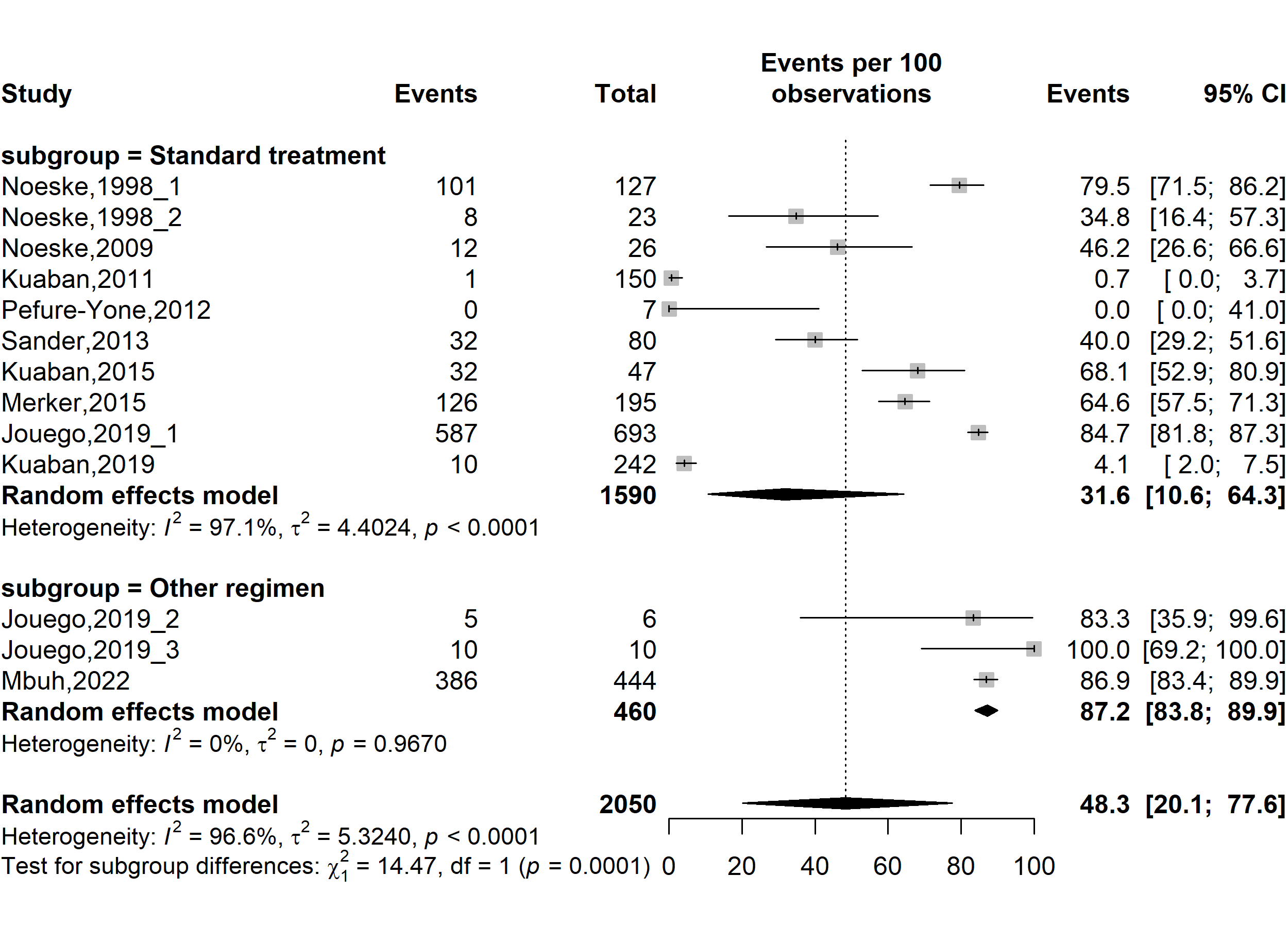

**Fig. S32** Forest plot displaying the subgroup meta-analysis of the pooled treatment failure rate among drug-resistant tuberculosis patients in Cameroon by types of resistant tuberculosis regimens, 1998-2022

(*Other regimen included fluroquinolone resistant modified treatment regimen [FQr-Mtr] and second-line injectable-resistant modified treatment regimen [SLIr-Mtr]*)

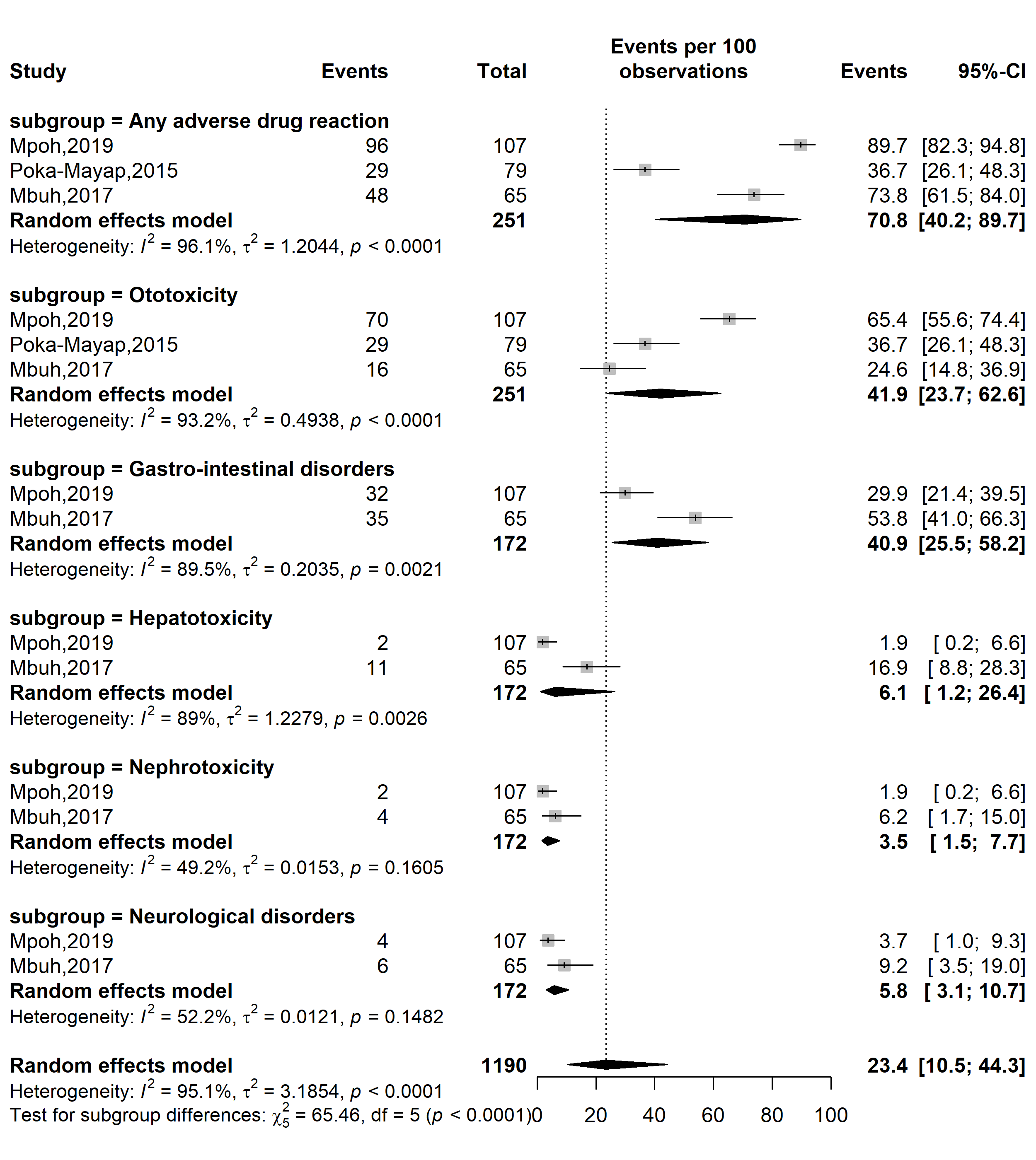

**Fig. S33** Forest plot displaying the prevalence of adverse drug events among patients treated for multidrug-resistant tuberculosis in Cameroon, 2015–2019

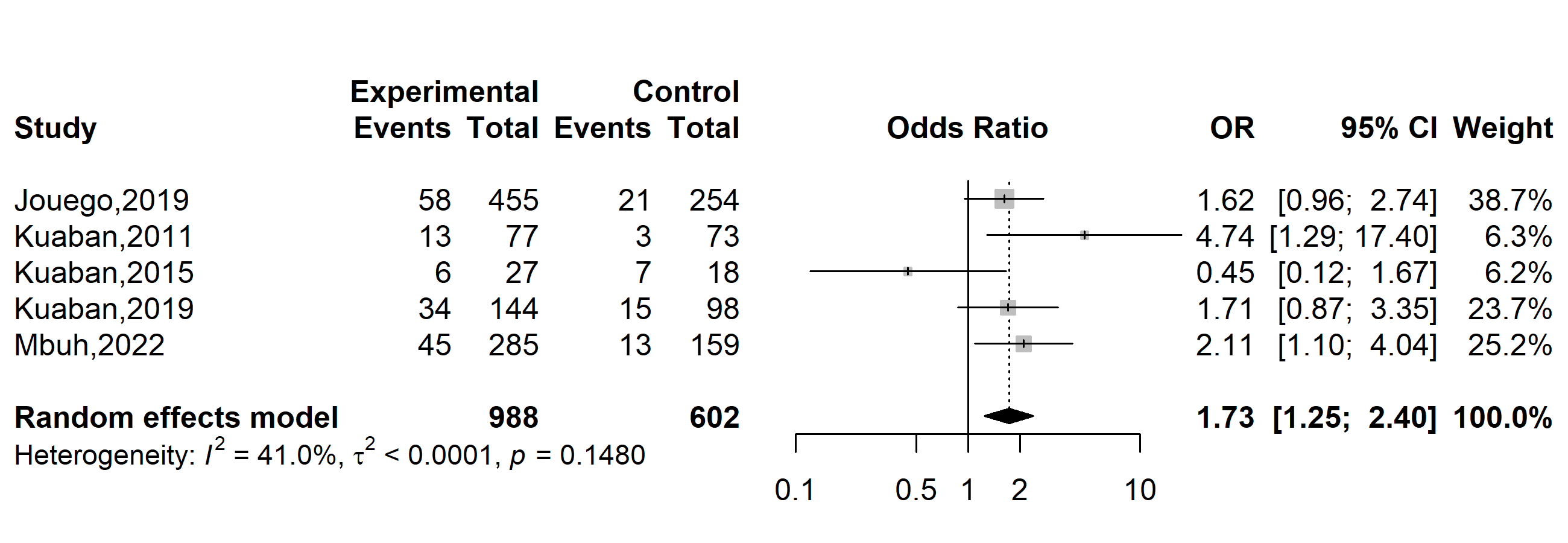

**Fig. S34** Pooled odds ratio of unfavorable outcome among patients treated for drug-resistant tuberculosis in Cameroon, 2015-2022 (Gender: Male vs. Female)

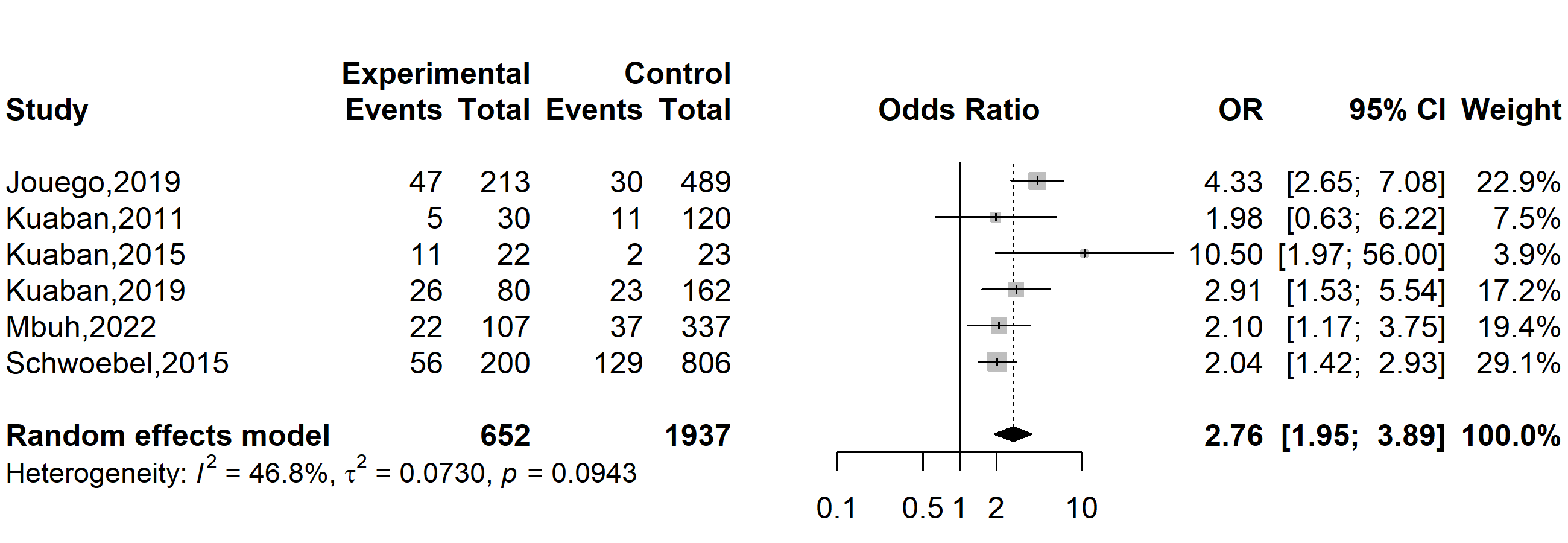

**Fig. S35** Pooled odds ratio of unfavorable outcome among patients treated for drug-resistant tuberculosis in Cameroon, 2015-2022 (Human immunodeficiency virus status: Positive vs. Negative)

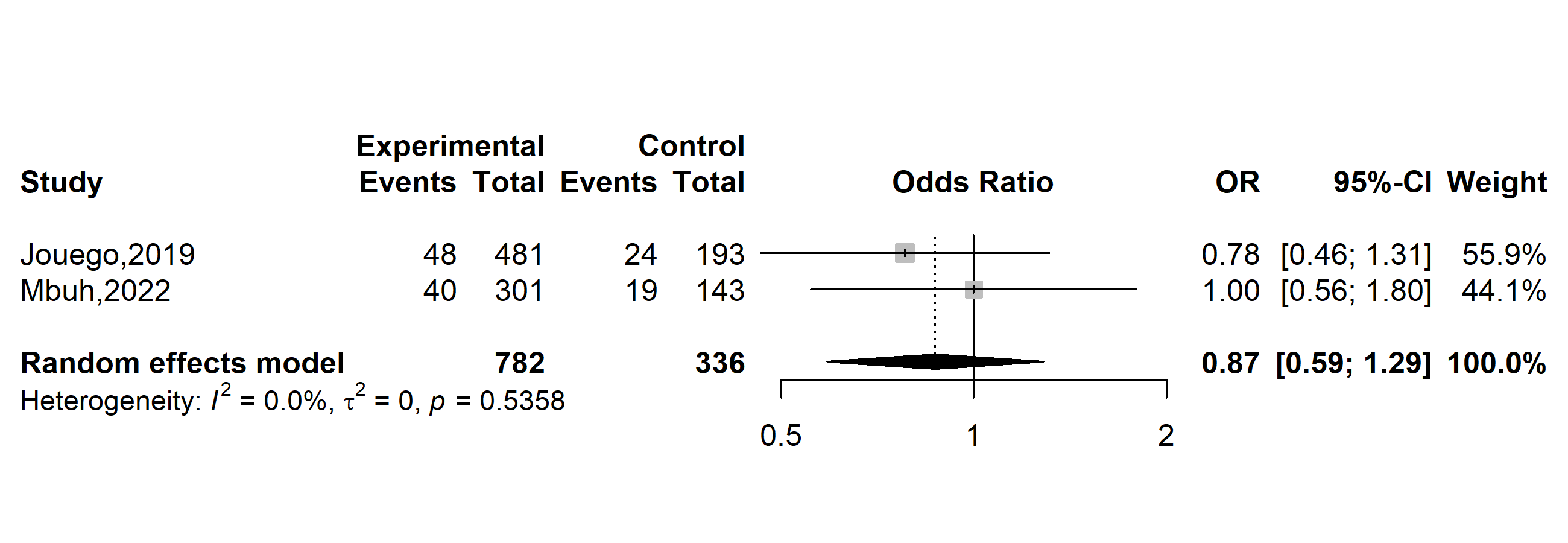

**Fig. S36** Pooled odds ratio of unfavorable outcome among patients treated for drug-resistant tuberculosis in Cameroon, 2015-2022 (Previous tuberculosis treatment: Yes vs. No)
